# Artificial intelligence in health professions education: *A state-of-the-art meta-review*

**DOI:** 10.1101/2025.10.20.25338371

**Authors:** M. Mahbub Hossain, Puspita Hossain, Tamal Joyti Roy, Jyoti Das, Samia Tasnim, Ping Ma, Winston Liaw

## Abstract

The growing adoption of artificial intelligence (AI) technologies in healthcare is transforming modern healthcare systems, necessitating current and future healthcare providers to be educated on the meaningful use of AI in their academic and professional activities. Despite an emerging body of literature emphasizing the use of AI in health professions education (HPE) and the availability of multiple reviews on this topic, there is a lack of meta-research evidence that can provide a broader overview of the evidence landscape reported across the existing systematically conducted literature reviews. This meta-review aimed to synthesize evidence on the applications of different AI technologies in HPE, multi-level factors influencing the applications of AI in HPE, and associated outcomes from existing systematically conducted literature reviews (SCLRs). A total of 48 eligible SCLRs were identified from six databases and additional sources, and the synthesized findings suggest emerging use cases of multiple AI technologies among HPE users and institutions, including AI-assisted instructional delivery, augmenting learning sessions, content optimization, and providing feedback. While most reviews reported positive HPE-related outcomes, there are critical challenges at the user and institutional levels, which should be considered for effective AI implementation in HPE. Building AI capacities among HPE users and facilitating AI resources development are critical for AI adoption. This meta-review may inform HPE and broader healthcare communities to advance knowledge and practice on evidence-based AI in HPE settings.

## Introduction

Artificial intelligence (AI) technologies are emerging as critical drivers of digital transformation across different frontiers of human lives globally. The evolving use cases of AI include making predictions and supporting decision-making, leveraging large bodies of data from diverse sources, and making their applications effective and scalable (1,2). Since the early stages of AI development, the healthcare community has been engaged in critical discourses on how human health and wellness can be promoted using AI (3–5). In recent years, notable progress has been made in AI applications in healthcare research with significant implications for policymaking and practice. Some of those advancements include analyzing or predicting patient-specific risks and outcomes, facilitating diagnoses, innovating advanced therapeutics (e.g., drug development, robotic surgery, etc.), developing personalized care plans, delivering health information and education, supporting patient-provider communication, and promoting shared decision-making (6–8).

Healthcare professionals and early-career trainers are increasingly recognizing the need for learning AI in the context of healthcare practice (9). Moreover, healthcare educators are emphasizing the need for integrating AI to advance health professions education (HPE) delivery and building capacities in institutions of higher education in healthcare. Previous research on AI applications in healthcare highlights a growing interest of the HPE community on the development of AI-related educational content and opportunities (10,11). Moreover, the use of AI-based tools and resources in HPE delivery in educational settings can potentially benefit HPE educators and students maximizing educational outcomes (12). Several reviews have been published summarizing these diverse use cases of AI applications in different areas of HPE (12–17); however, a broader synthesis of existing meta-research evidence appeared to be unavailable upon a structured search in the PubMed database.

A meta-review, also known as a systematic review of reviews, of existing meta-research studies on AI use in different frontiers of HPE can substantially benefit scholarship and practice in several ways. First, a systematic overview of existing systematically conducted literature reviews (SCLRs) can inform the current state-of-the-art evidence on AI applications in HPE, which can support the adoption of evidence-based interventions and resources for advancing HPE. Second, evaluating the current evidence base would inform the evidence gap; therefore, mapping the existing development of AI in HPE can possibly facilitate future research and development addressing those gaps. Third, the available evidence on AI use in HPE can suggest high-value areas of AI deployment for a wide range of individual and organizational outcomes, thus supporting HPE decision-makers to assess the incremental value of AI applications within the scope of their strategic initiatives. Lastly, examining the factors associated with AI use in HPE can enable the key stakeholders to address potential barriers to implementing AI and develop optimal strategies for effective use of AI in HPE. Therefore, the objective of this meta-review was to synthesize the existing meta-research evidence on AI applications in different areas of HPE. Specifically, this meta-review aims to answer the following questions: a) what are the applications of different AI technologies in health professions education (HPE)?, b) what are the outcomes of AI applications in HPE?, and c) what are the factors influencing the applications of AI in HPE?

## Methods

This meta-review was conducted using a protocol pre-registered with the Open Science Framework (available at: osf.io/dkbmn), and reported in accordance with the Preferred Reporting Items for Systematic reviews and Meta-Analyses (PRISMA) guidelines (18).

## Key concepts of this meta-review

### Types of participants

This meta-review considered SCLRs that focused on participants related to HPE, including but not limited to health professions students, educators, administrators, and other stakeholders. Given the significant heterogeneity in academic roles, the level of utilization of educational resources and technologies, and possible impacts, this review included SCLRs that recruited articles representing different participants from HPE. Moreover, the scope of HPE was considered broadly as it may consist of all possible forms of educational levels from diverse cadres of health professions, including medical education, nursing education, dental education, public health and allied health professionals, amongst others.

### Types of interventions/phenomena

This meta-review included SCLRs that primarily examined any form of applications of AI in the context of HPE. Such applications included the use of AI-based learning management systems, AI-enabled classrooms or learning spaces, AI-integrated tools and resources used within the scope of HPE, capacity building resources for using AI in any area of healthcare as a part of HPE (e.g., AI curricula in HPE), among other possible use cases. AI tools and resources are being used by HPE students, educators, and programs in many areas of HPE (e.g., AI-enabled learning management systems or digital tutoring services). In addition, development and implementation of AI tools and related resources (e.g., AI-related curricula and learning resources that influence AI use in HPE users) are critical epistemic and practice areas, which may inform capacity building for AI use in HPE. In such cases, the primary goal of eligible articles had to be aligned with the broader objectives of this review providing insights on AI and related resources utilization in the context of HPE. This meta-review recognized the evolving state of AI and its applications across different structural and functional levels of HPE, and aimed to include all possible applications to provide inclusive meta-evidence.

### Type of comparisons

This meta-review considered all comparative scenarios, including studies or reviews that did not report or specify any comparison groups.

### Types of outcomes

This meta-review included any outcomes associated with AI use in HPE as a part of addressing one of the core objectives of the review. Such outcomes included educational, health, and organizational outcomes associated with AI use in HPE. Moreover, this review reported the state of evidence on AI use in HPE and associated factors, which were considered as the knowledge outcomes of this review.

### Types of studies

This meta-review included all forms of SCLRs, such as traditional systematic reviews, meta-analyses, meta-syntheses, meta-aggregative reviews, scoping reviews, critical or integrative reviews, and other forms of literature reviews that report a systematic process of literature search and synthesis.

## Eligibility criteria

This meta-review included articles that fulfill all the following criteria:

1. Focused on any area of HPE in any geographic region or population,
2. Primarily focused on the use of AI-based technologies or related resources in HPE as described earlier,
3. Were published in any form of SCLRs,
4. Published as peer-reviewed journal articles, and
5. Full text was available in the English language.

## Search strategy

This meta-review deployed a structured search strategy (outlined in Table 1) in Medline, American Psychological Association (APA) PsycInfo, Cumulative Index to Nursing and Allied Health Literature (CINAHL), Cochrane Database of Systematic Reviews, Web of Science, and Scopus. In addition, Google Scholar and other resources, such as cited articles and references of the selected articles, were searched to identify additional articles that might be eligible for this meta-review. No language, geographic, time, or other possible restrictions were applied at this stage. The preliminary search was conducted on 01/17/2025 to assess the feasibility of this meta-review and the final search was conducted on 05/01/2025.

**Table 1:** Search strategy using specific keywords and appropriate Boolean operators.

| Search query | Concept(s) | Keywords (used in titles, abstracts, topics, author-provided keywords, medical subject headings, and other keywords) |
| --- | --- | --- |
| 1 | **Artificial intelligence** | "artificial intelligence" OR "machine learning" OR "deep learning" OR "ChatGPT" or "large language model*" OR "LLM*" OR "natural language processing*" or "neural network*" or chatbot* or "supervised learning*" OR "unsupervised learning*" OR "semisupervised learning*" OR "artificial general intelligence" |
| 2 | **Health professions education** | "health professions education*" OR "health professions student*" OR "medical education*" OR "medical school*" OR "medical curricul*" OR "medical student*" OR "nursing education*" OR "nursing school*" OR "nursing curricul*" OR "nursing student*" OR "dental education*" OR "dental school*" OR "dental curricul*" OR "dental student*" OR "clinical education*" OR "clinical school*" OR "clinical curricul*" OR "clinical student*" OR "allied health education*" OR "public health education*" OR "health services education*" OR "healthcare professional education" |
| 3 | **Systematically conducted literature reviews (SCLRs)** | "systematic review*" OR "systematic literature review*" OR "meta-analy*" OR "metaanaly*" OR "quantitative review*" OR "mixed methods review*" OR "qualitative review*" OR "meta-synthesis*" OR "pooled prevalence" OR "pooled effect*" OR "pooled estimate*" OR "scoping review*" OR "critical review*" OR "meta-aggregat*" OR "critical interpretive synthesis*" OR "integrative review*" OR "evidence-based review*" OR "synthesized evidence" |
| **Final search strategy** | **1 AND 2 AND 3**<br>**(with no additional search restrictions)** |  |

## Study selection

All retrieved citations were uploaded to a cloud-based review screening tool (Rayyan.ai) and have been evaluated independently by two reviewers using the pre-determined eligibility criteria. Any conflicts at this stage were reviewed by a third reviewer and addressed through iterative consultations between these three reviewers. In case of any further conflicts, other review team members with related expertise were consulted as needed. Following this round of review, all full texts of the initially screened citations were retrieved and reviewed by three reviewers, and decisions about their inclusion were made based on consensus. The references of the finally selected articles were further assessed to identify potential citations that might be relevant to this review. Furthermore, the full texts of all such citations were assessed by three reviewers, and articles meeting all review criteria were retained for data extraction.

## Data extraction and synthesis

A pre-designed tool was used for data extraction from each included article, which was pilot-tested first using at least five articles and modified further for inclusive and extensive data extraction. Two reviewers independently extracted data from each article, followed by an iterative review with the help of a third team member. Possible inconsistencies in data extraction were addressed through discussion and revisiting each article as a group whenever necessary. The final set of extracted data was narratively synthesized and presented, addressing the objectives of this meta-review.

## Results

### Overview of the included articles

A total of 1894 citations were identified from database searching, which included 645 duplicates and 1051 non-eligible citations following de-duplication. Further, 86 citations were retained for full-text assessment and 50 of those were considered ineligible. This process yielded 36 eligible articles. Moreover, an additional 2072 citations were retrieved from Google Scholar, which provided 12 additional articles found eligible after full-text review. The process of literature review is illustrated in Figure 1. The final corpus of eligible articles included 48 SCLRs that met all inclusion criteria, and were published between 2018 and 2025 (please see Table 2). Among the included articles, 23 articles were published in 2025, and 12 articles were published in 2024, indicating the increase in research on AI use in HPE in recent years. The included review articles included research from various countries; however, most of the articles were focused on high-income countries of North America, Europe, and Asia. On the other hand, only two articles included papers from the African continent (17,19).

**Figure 1:**
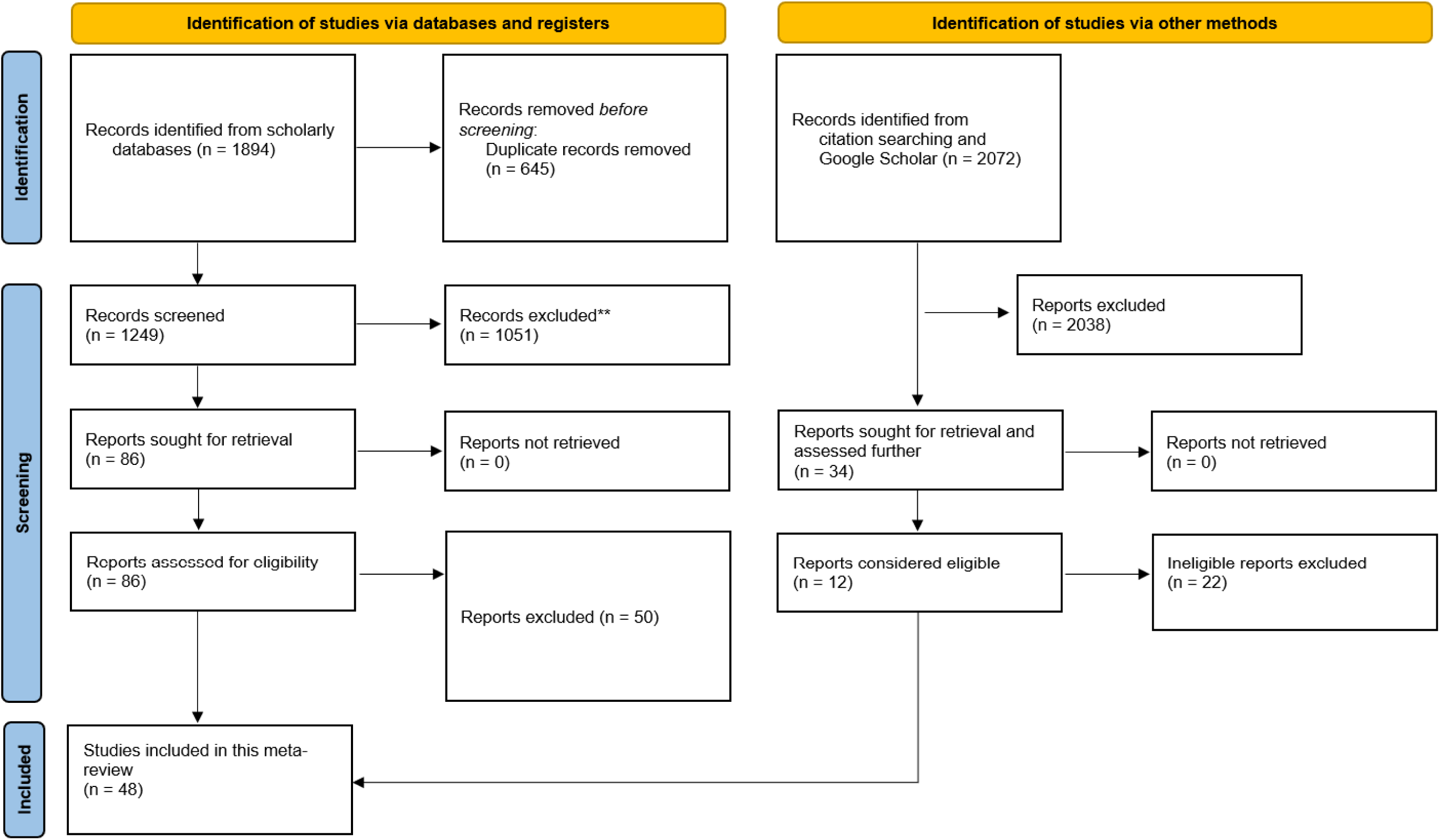
Flow diagram of the literature review process

**Table 2:**
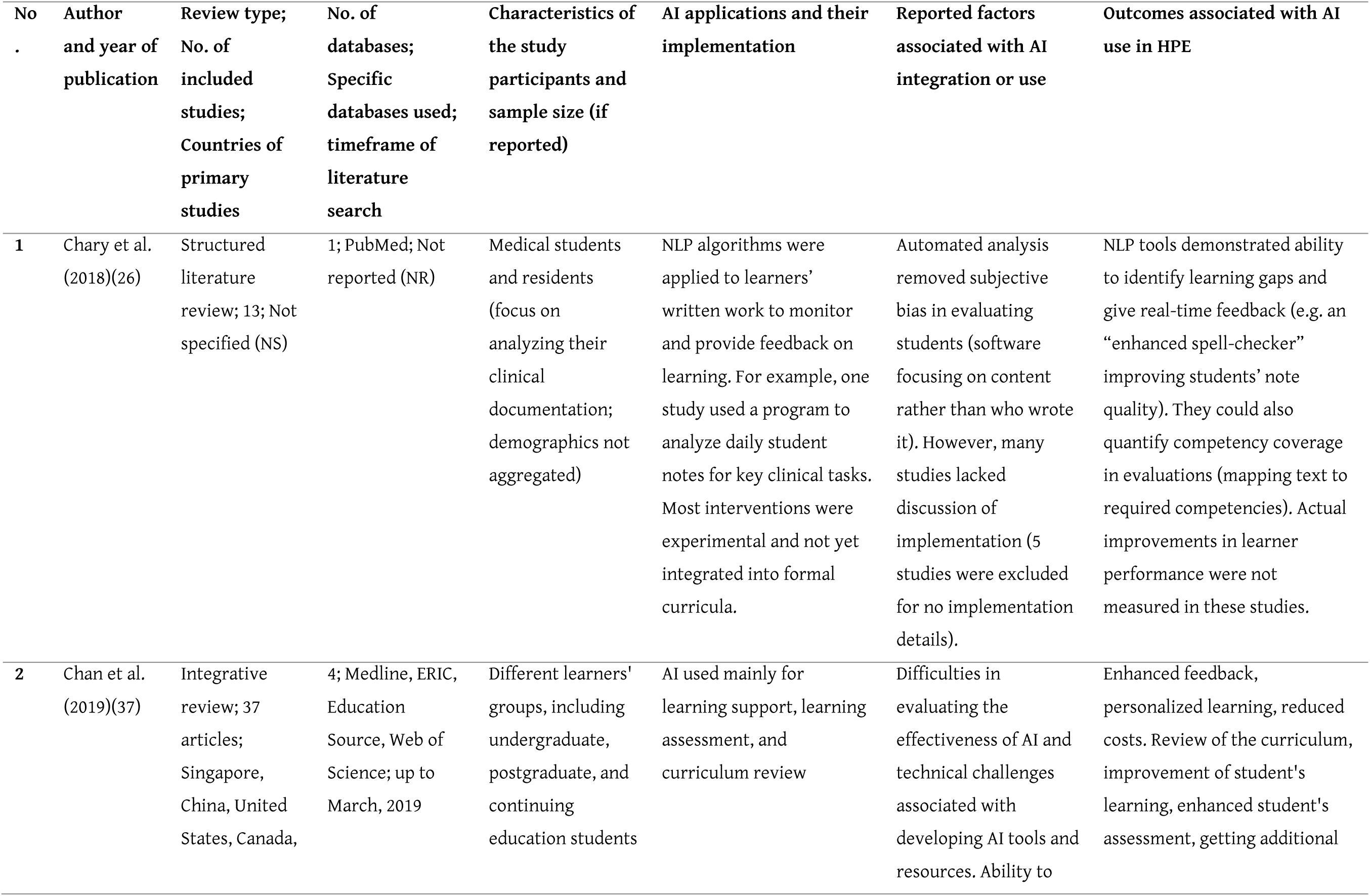
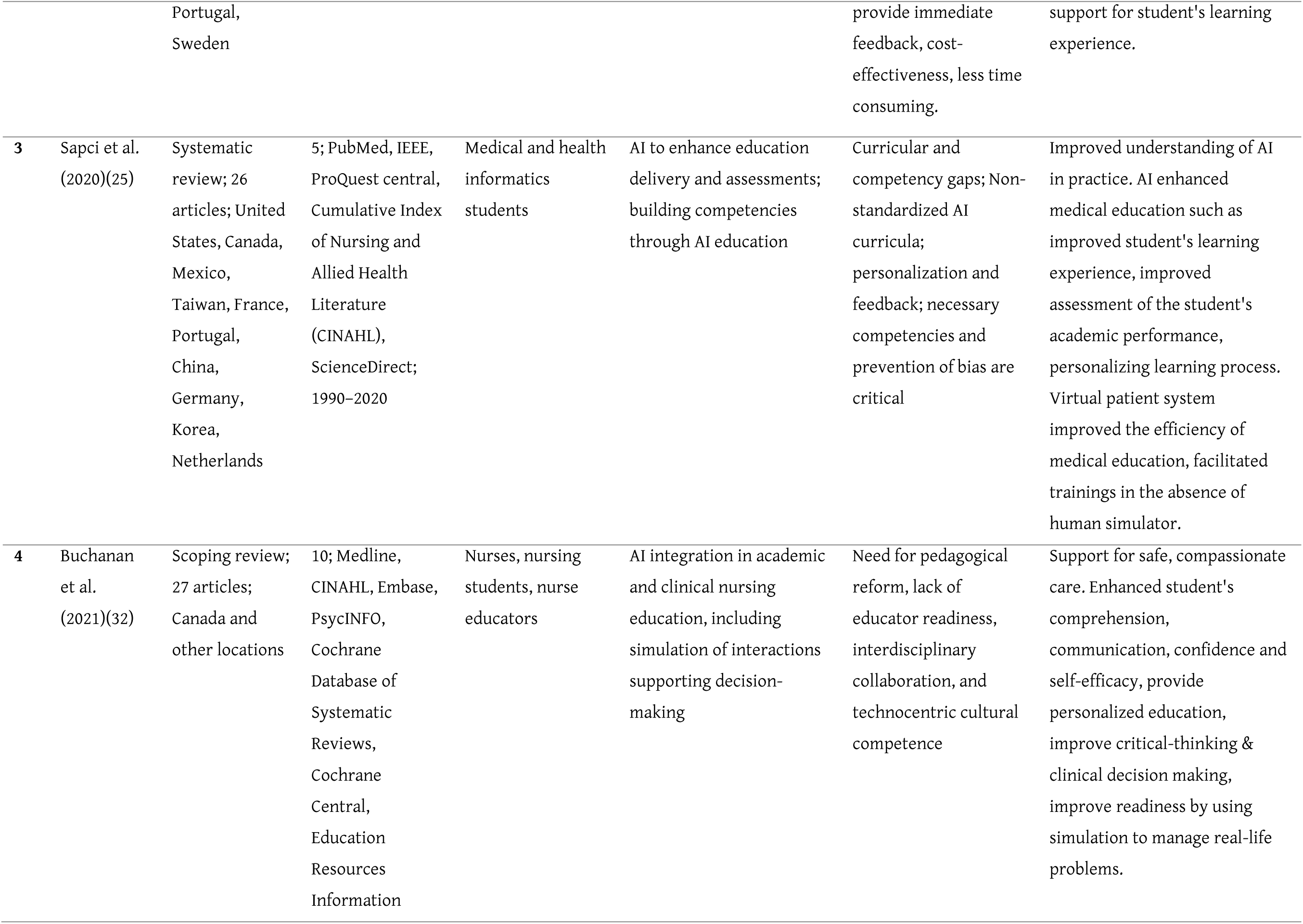
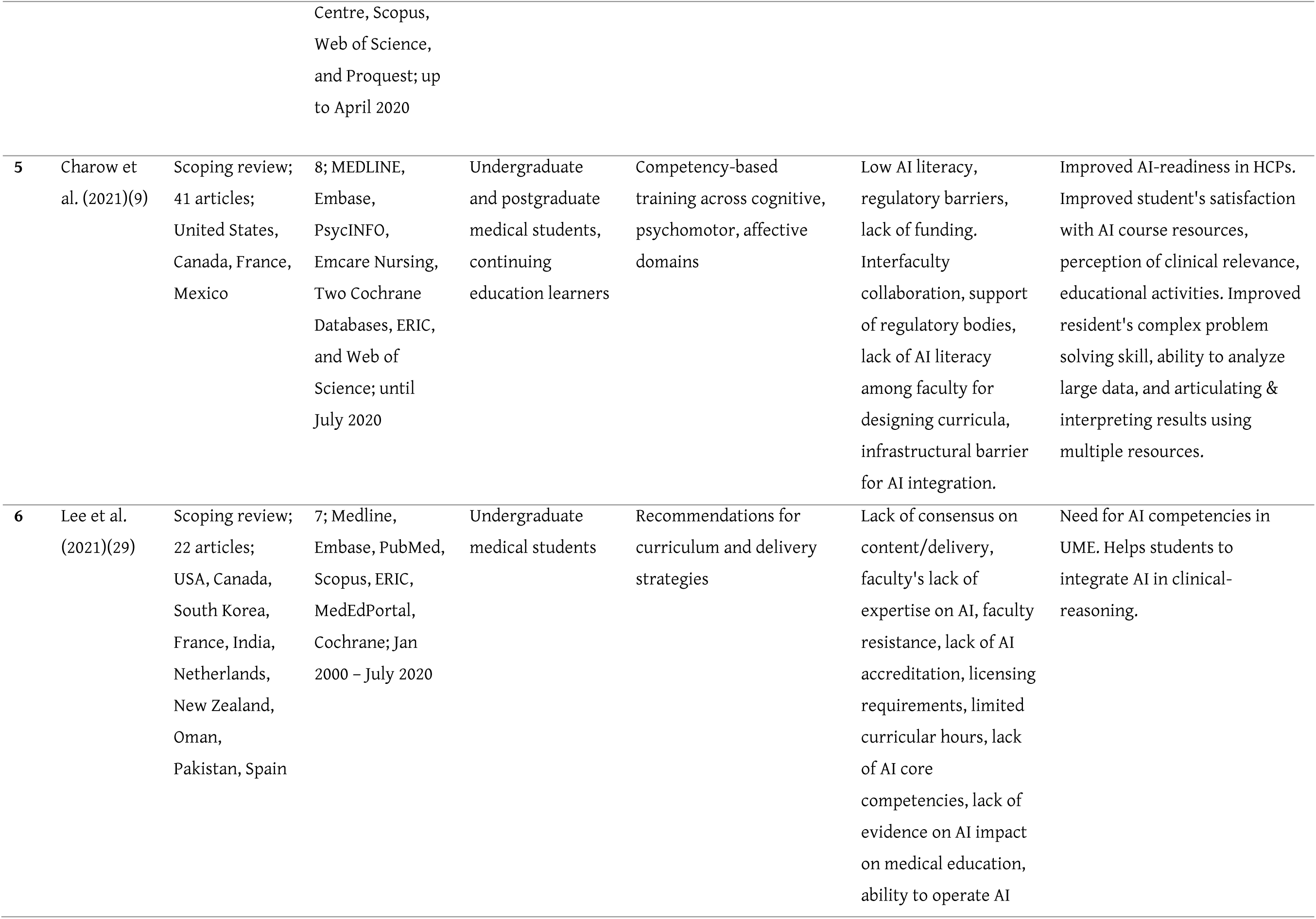
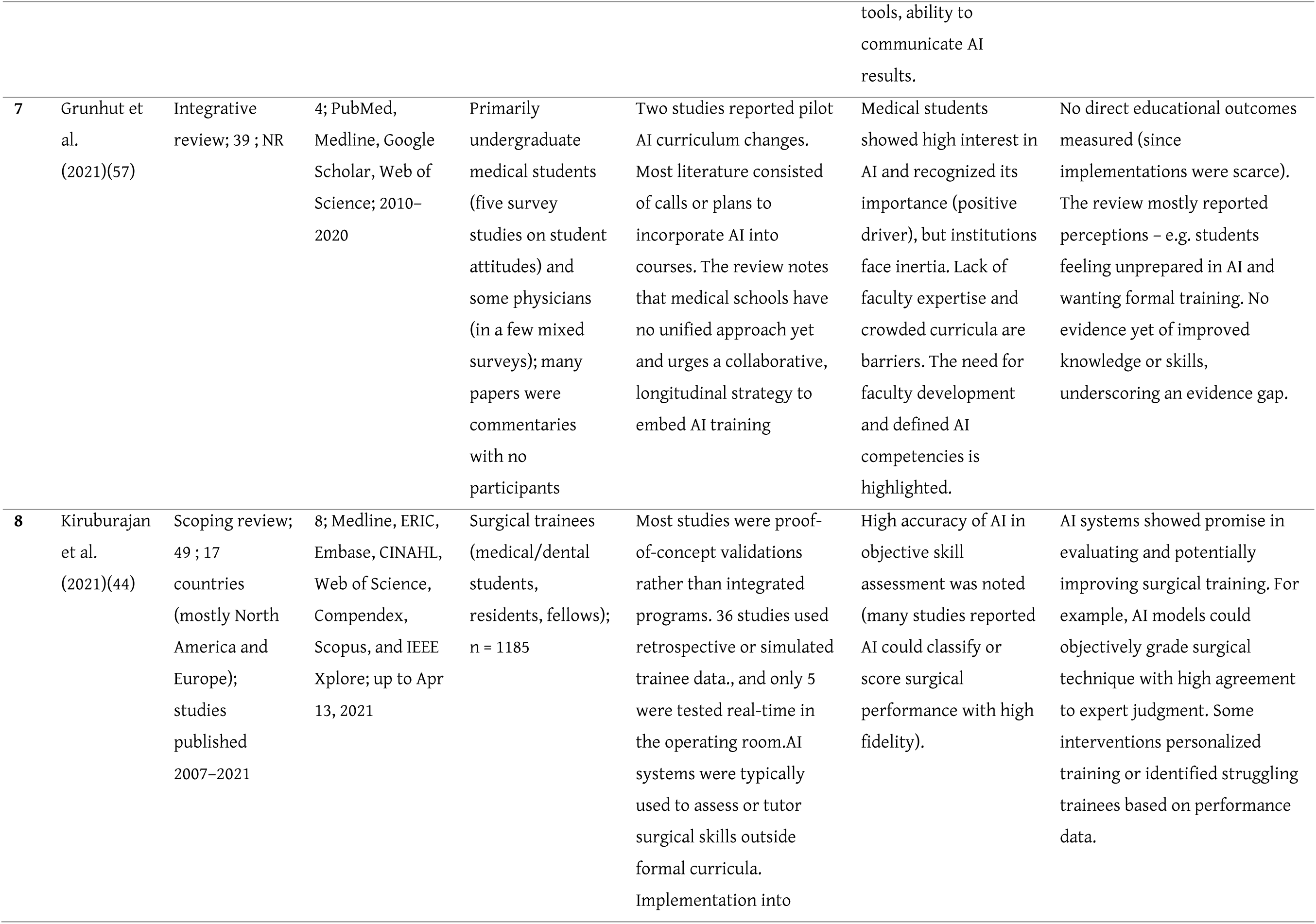
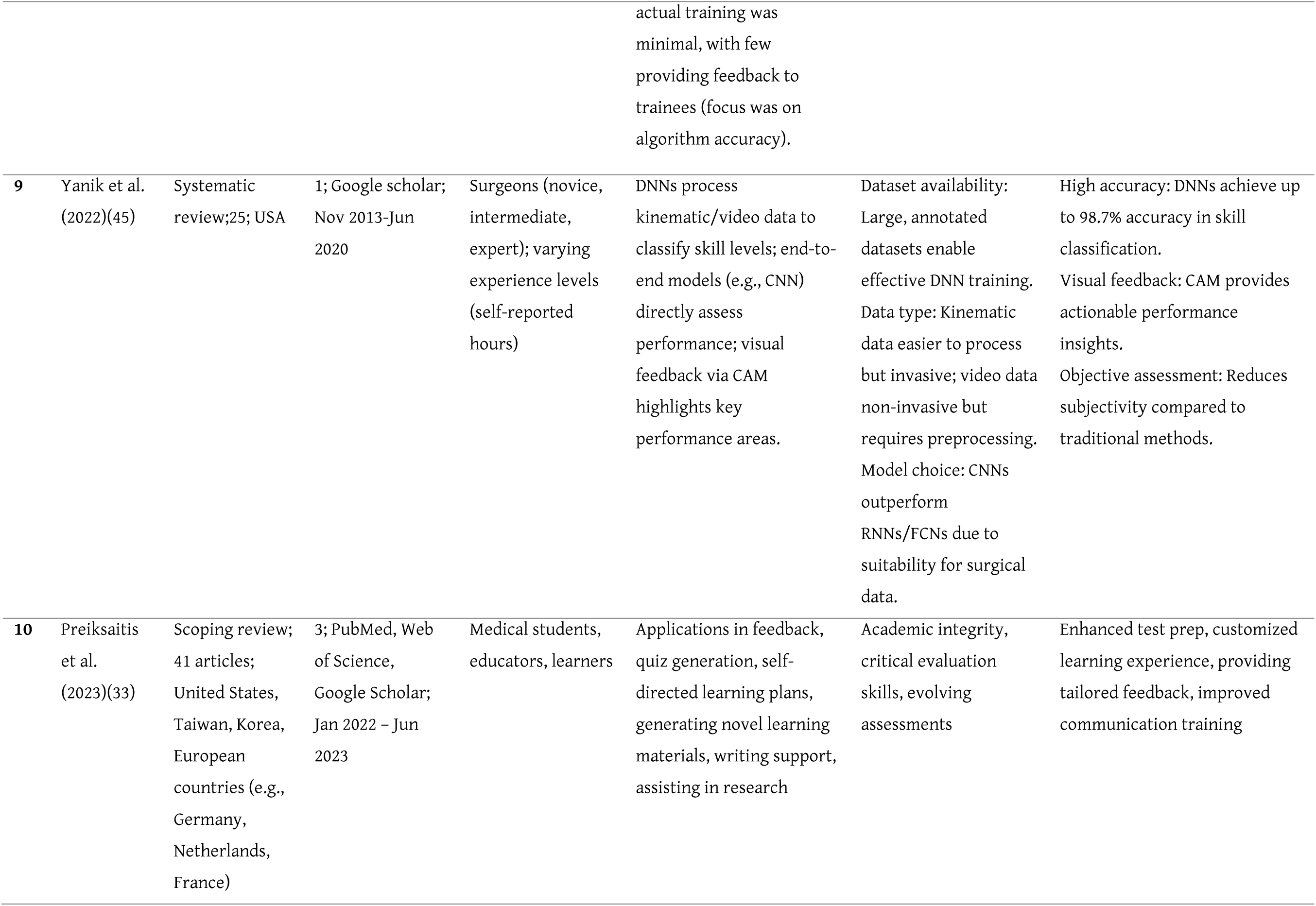
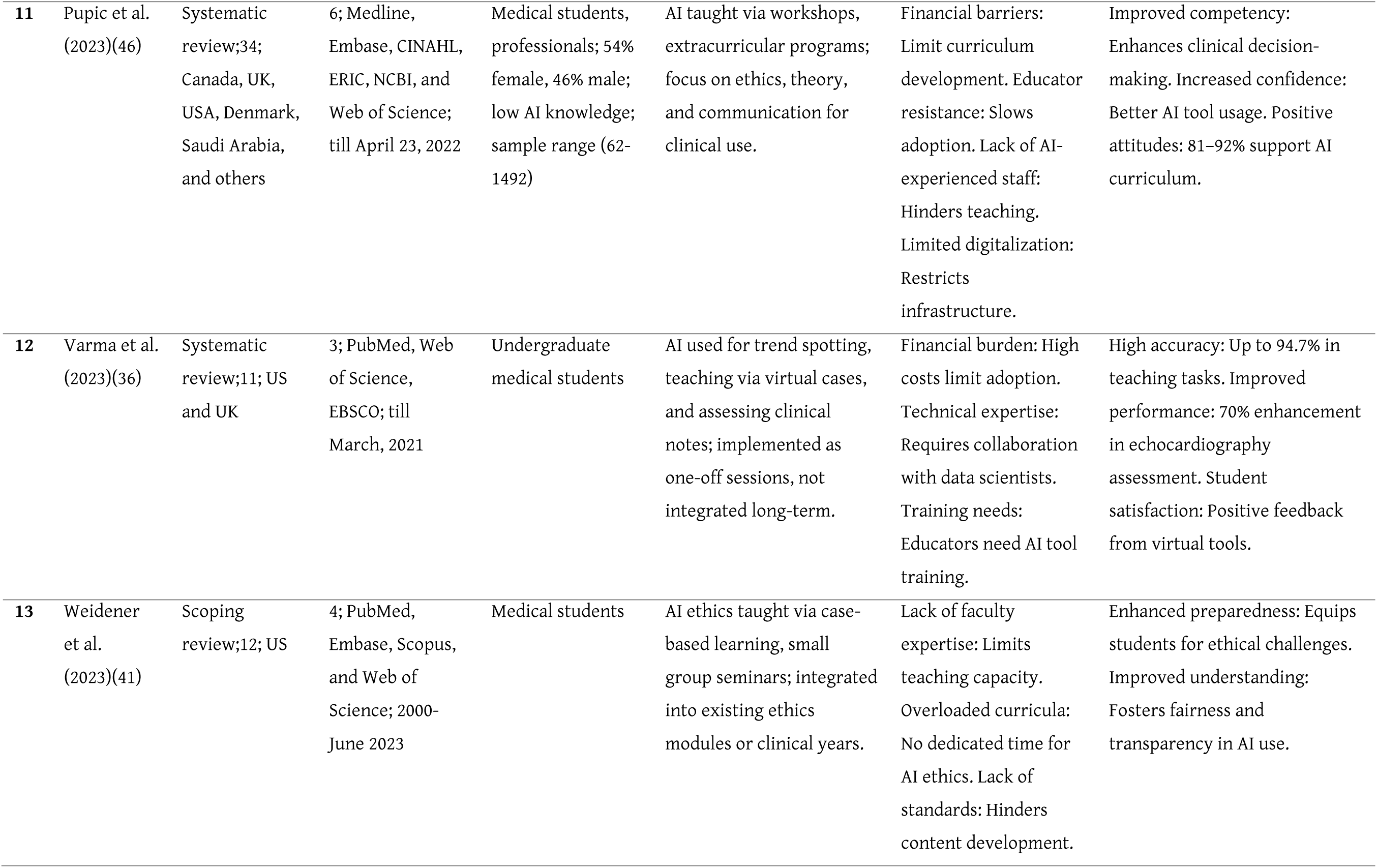
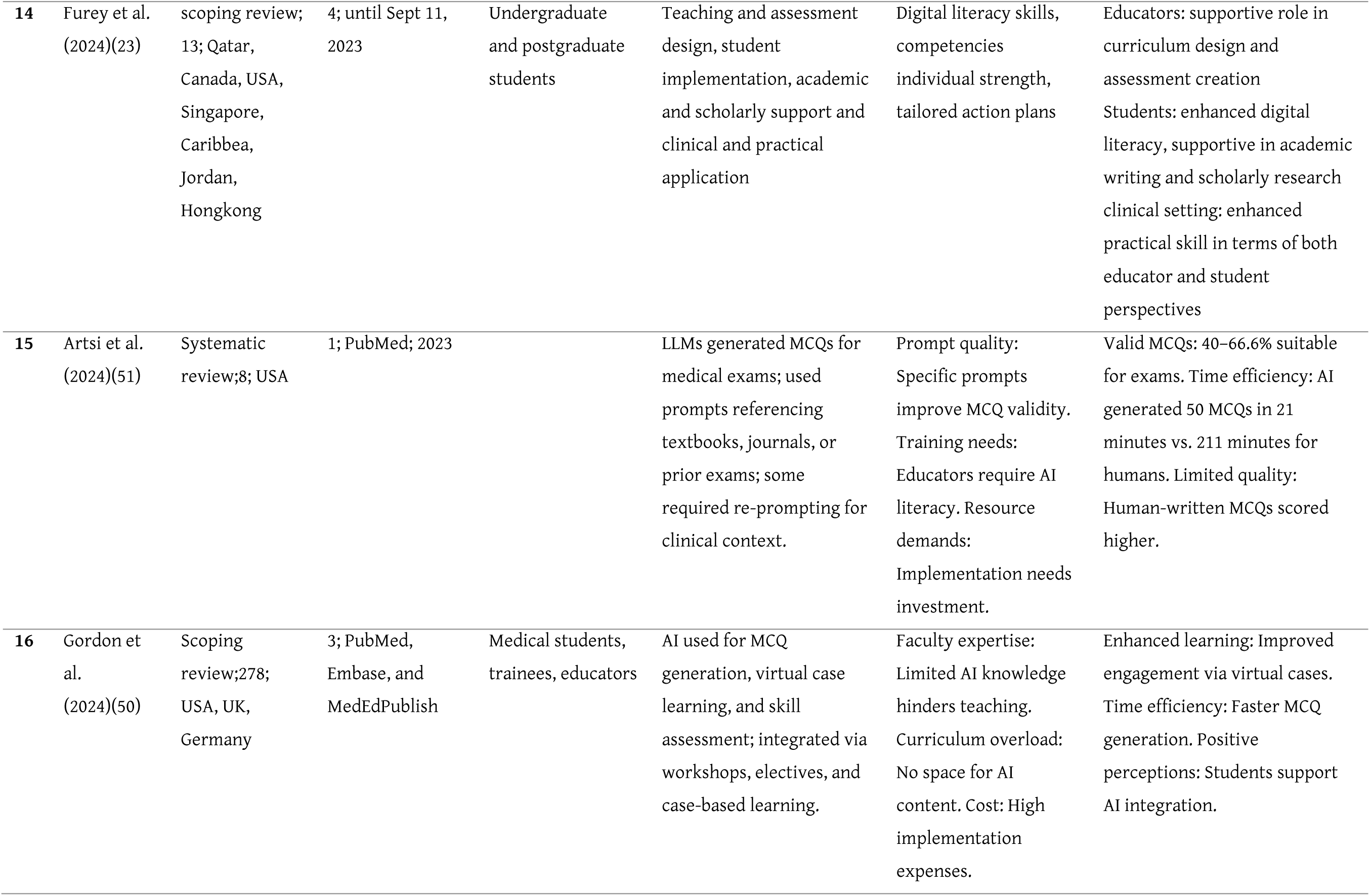
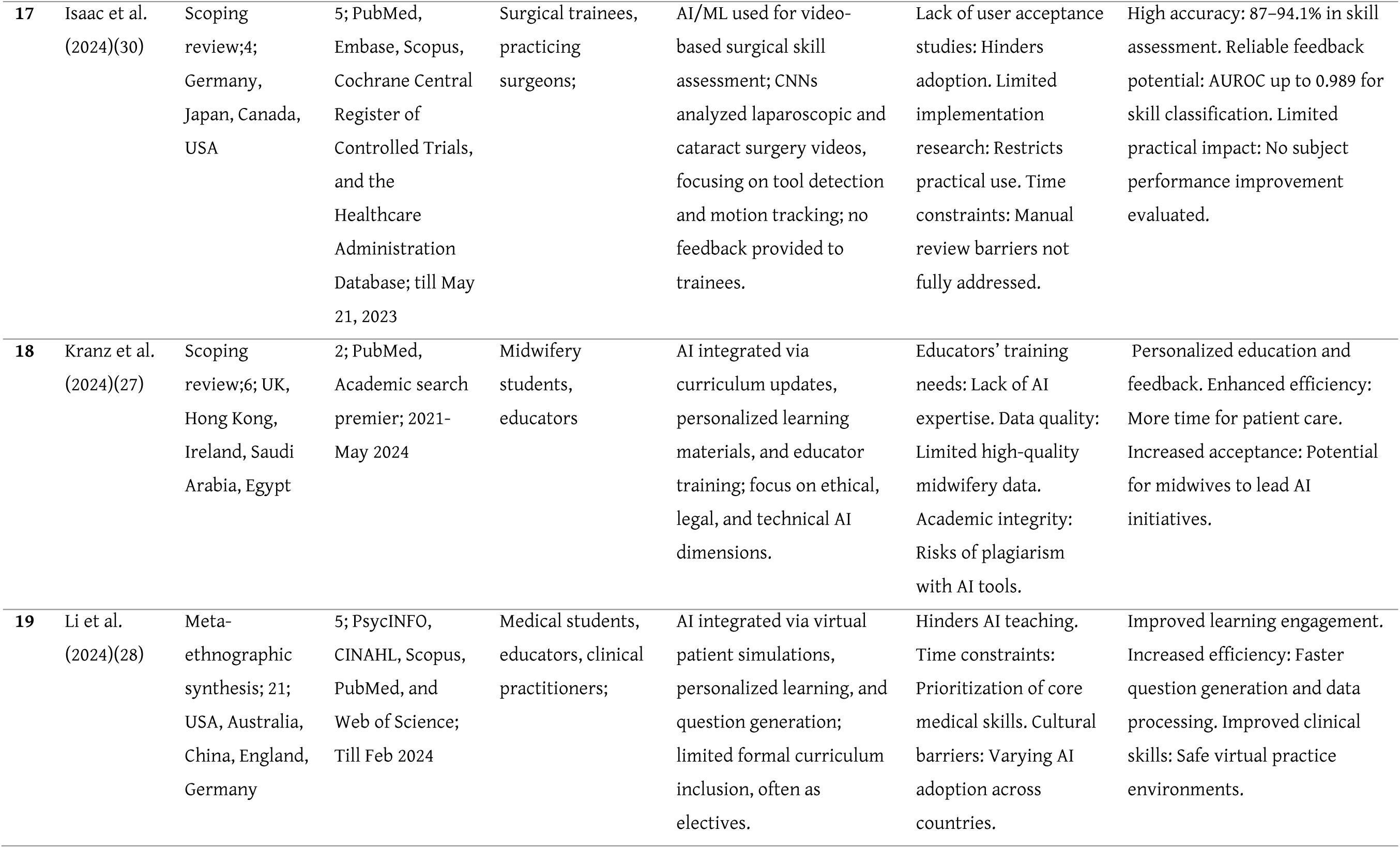
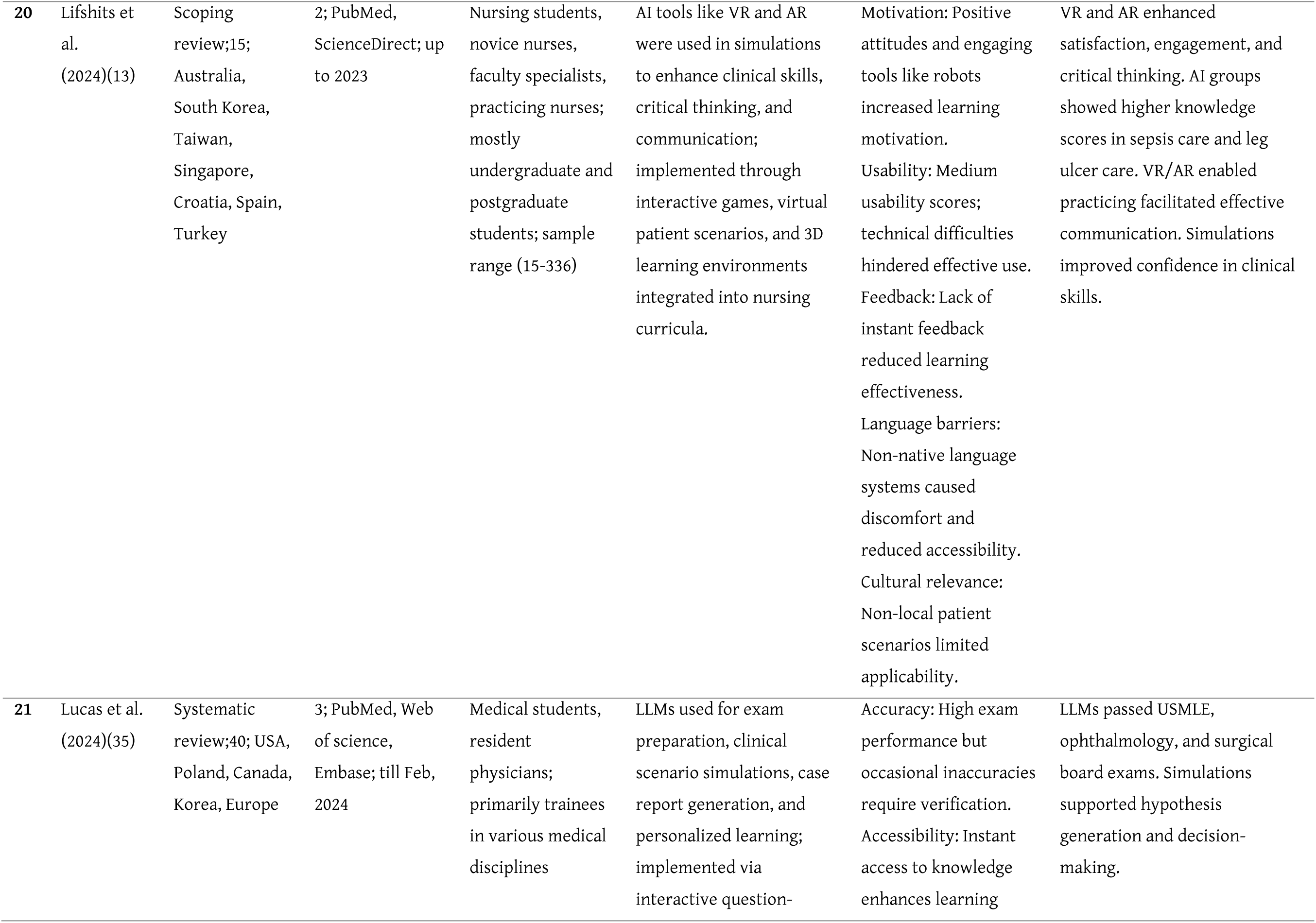
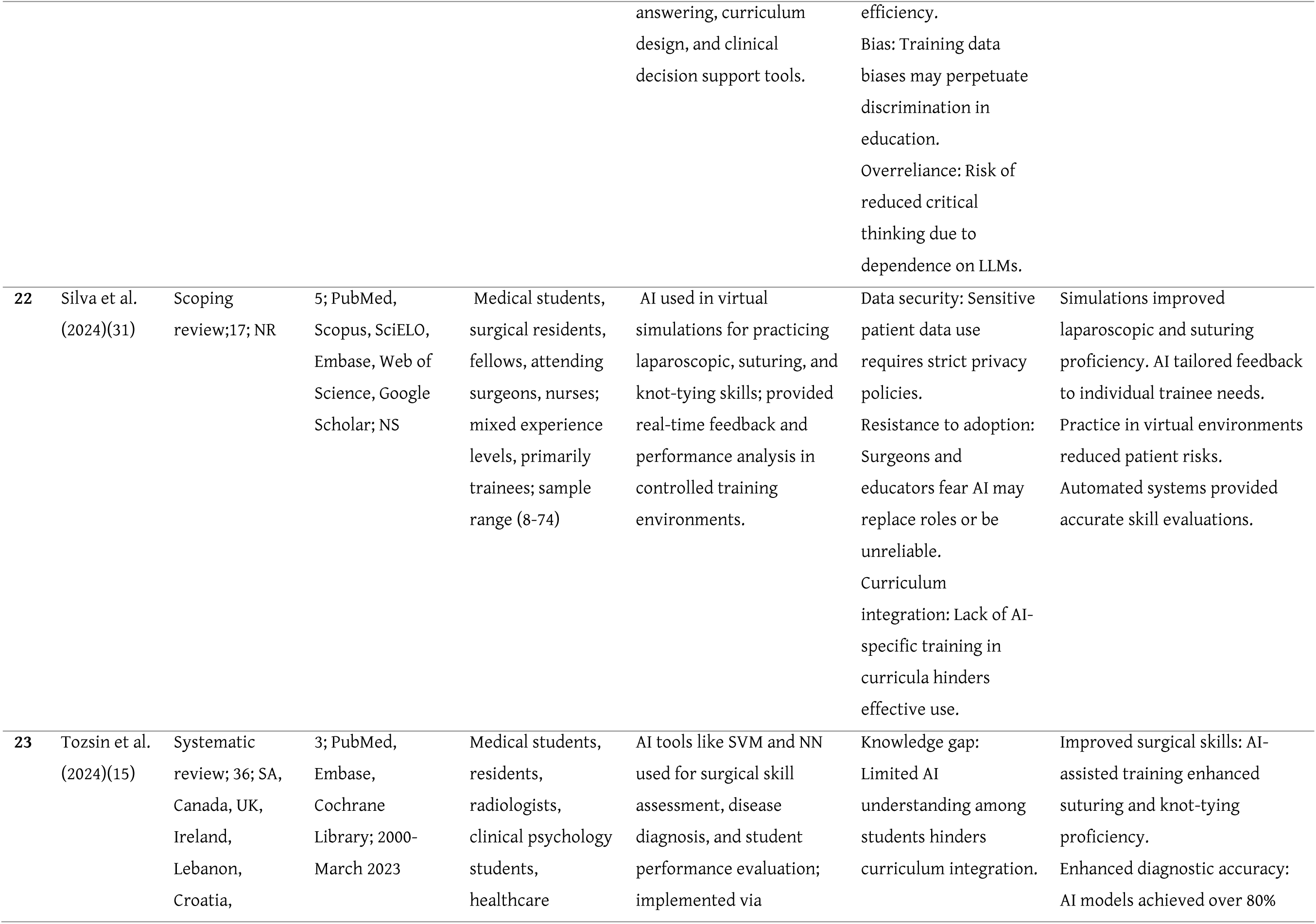
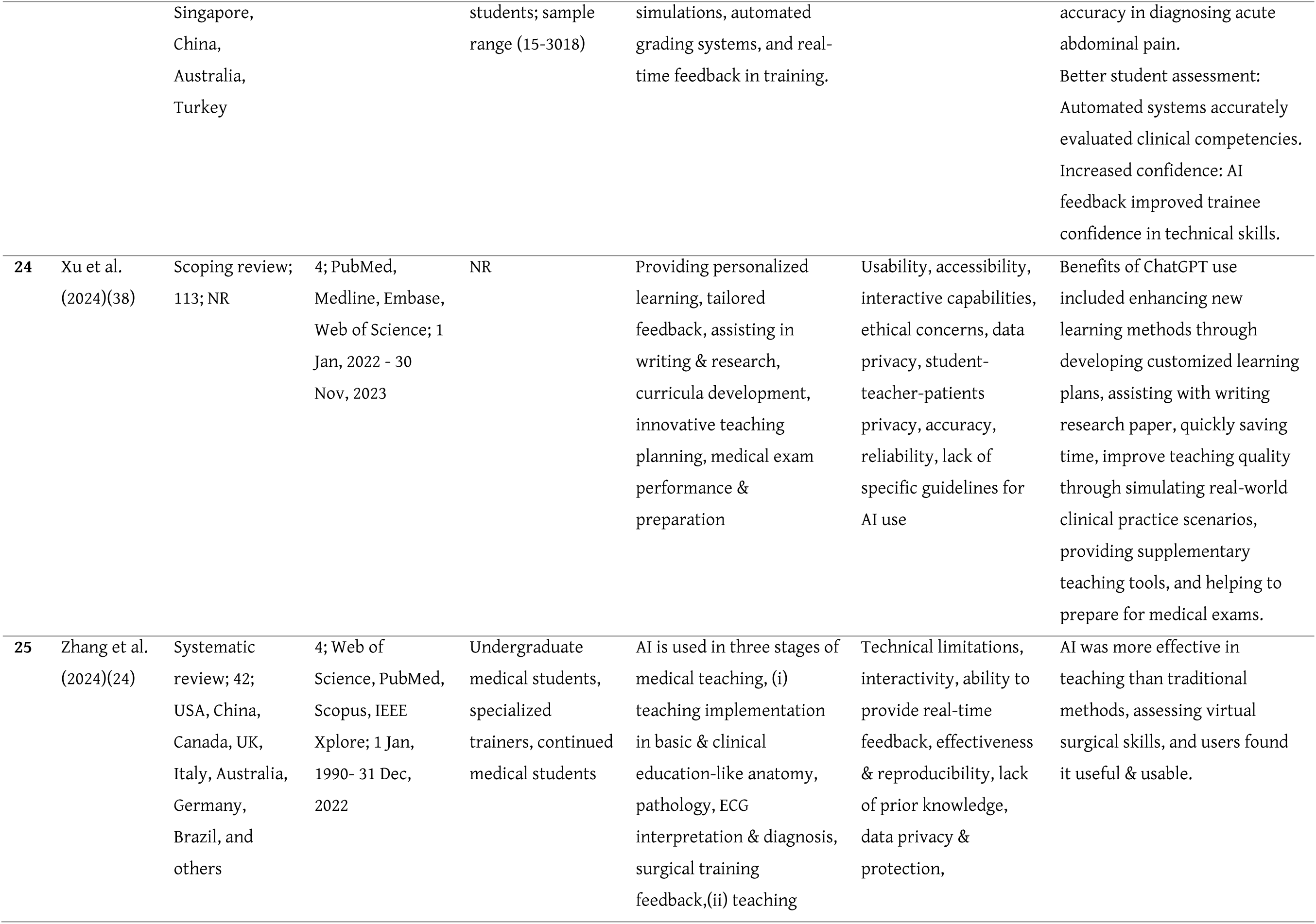
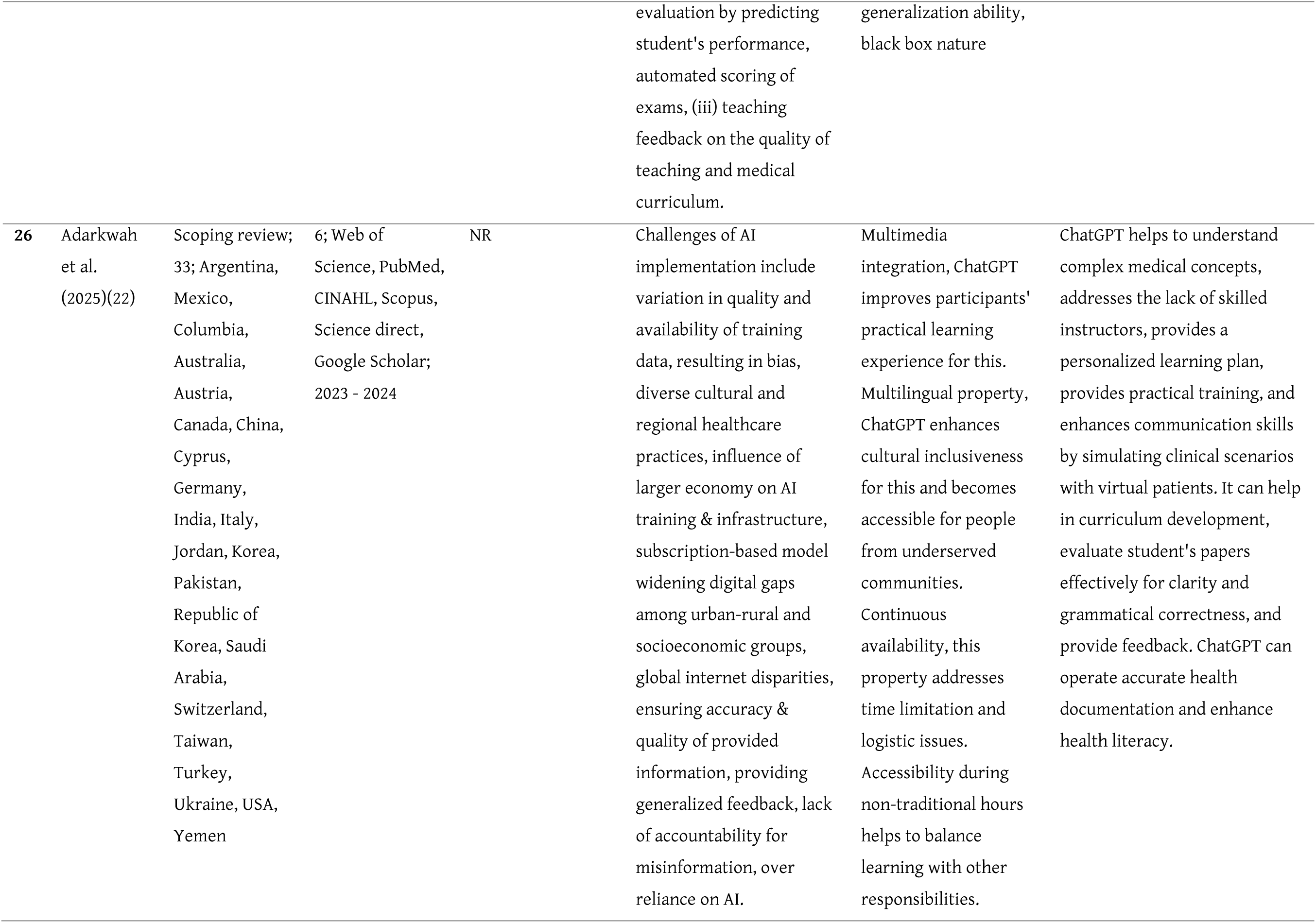
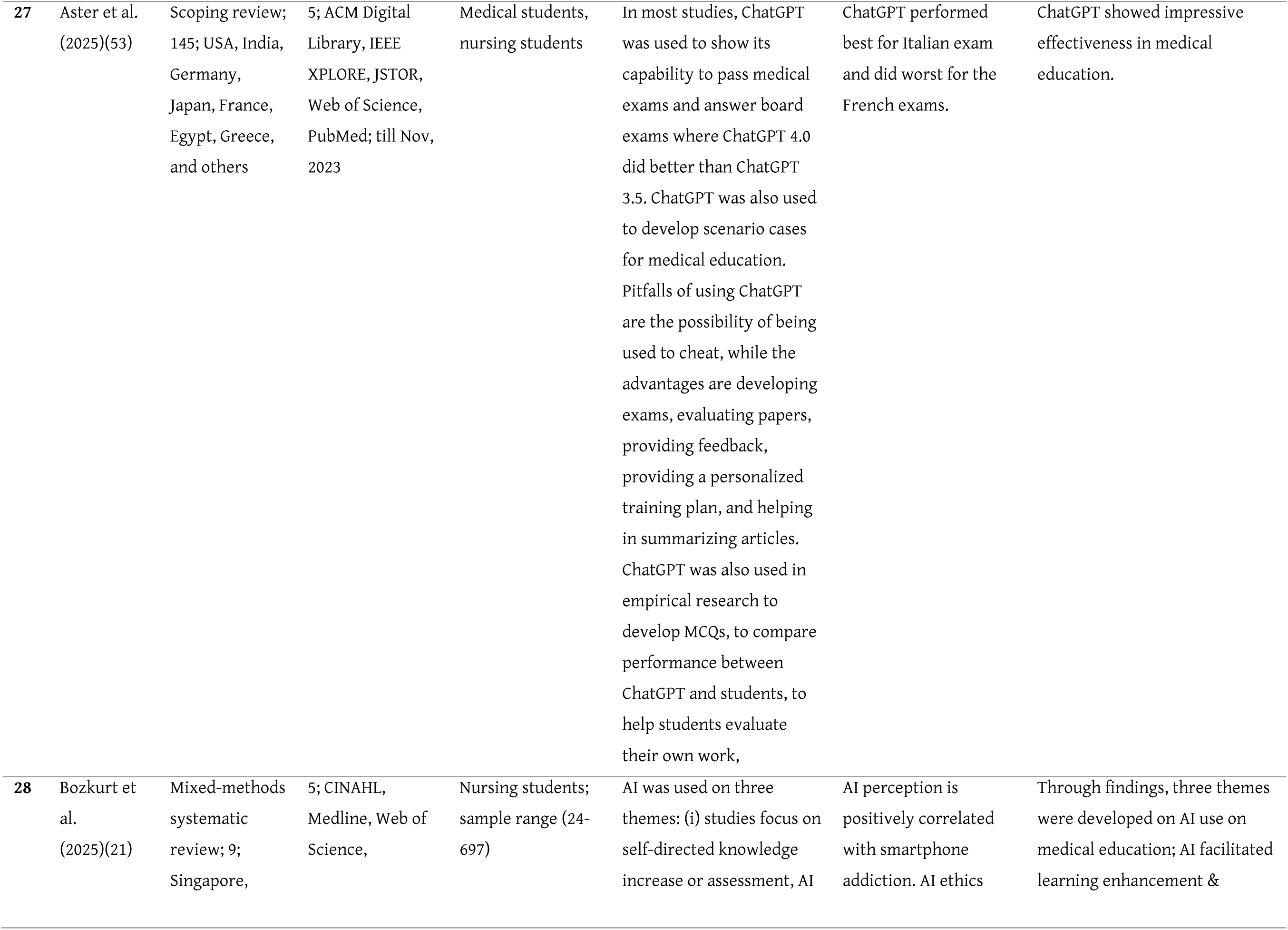
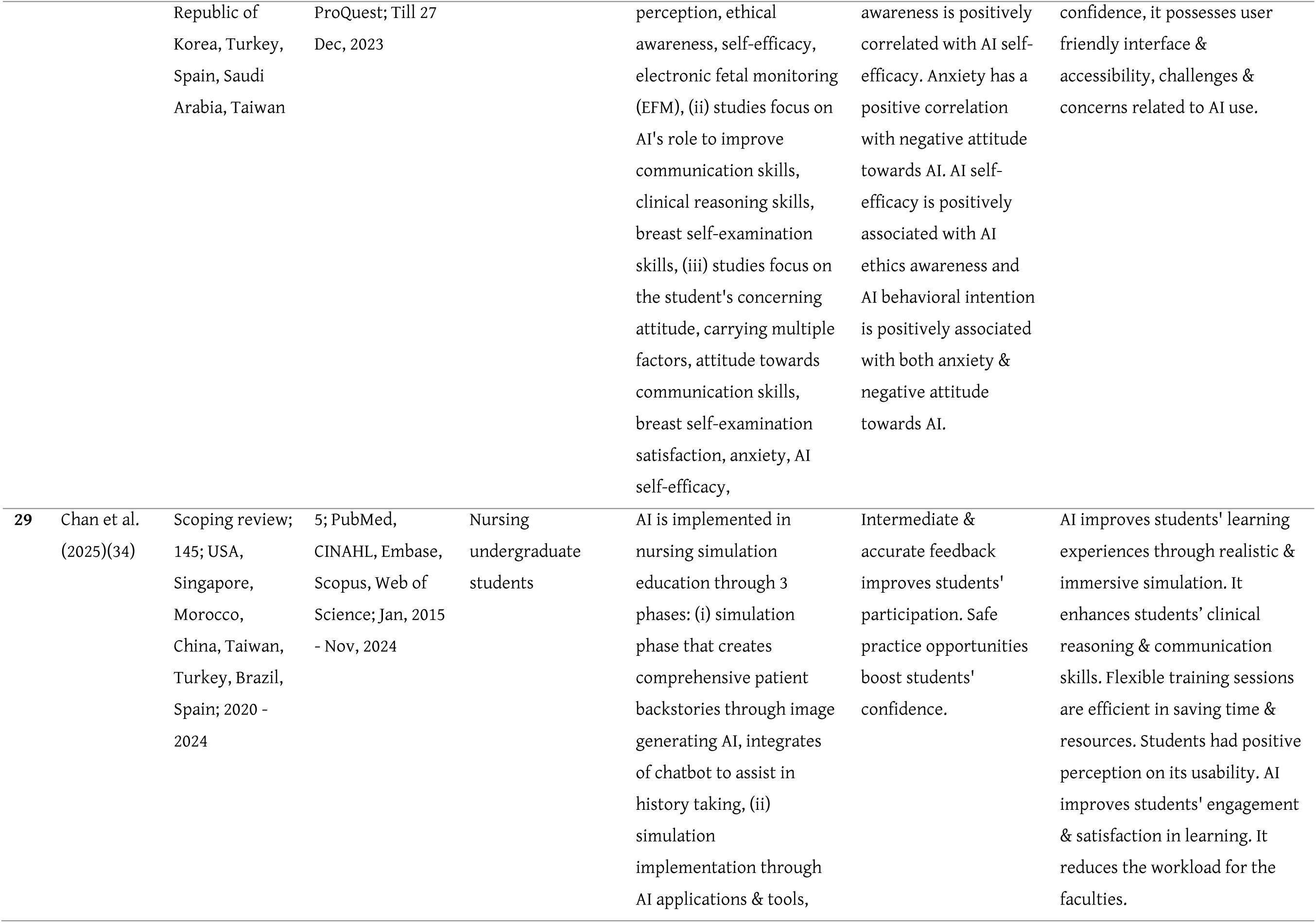
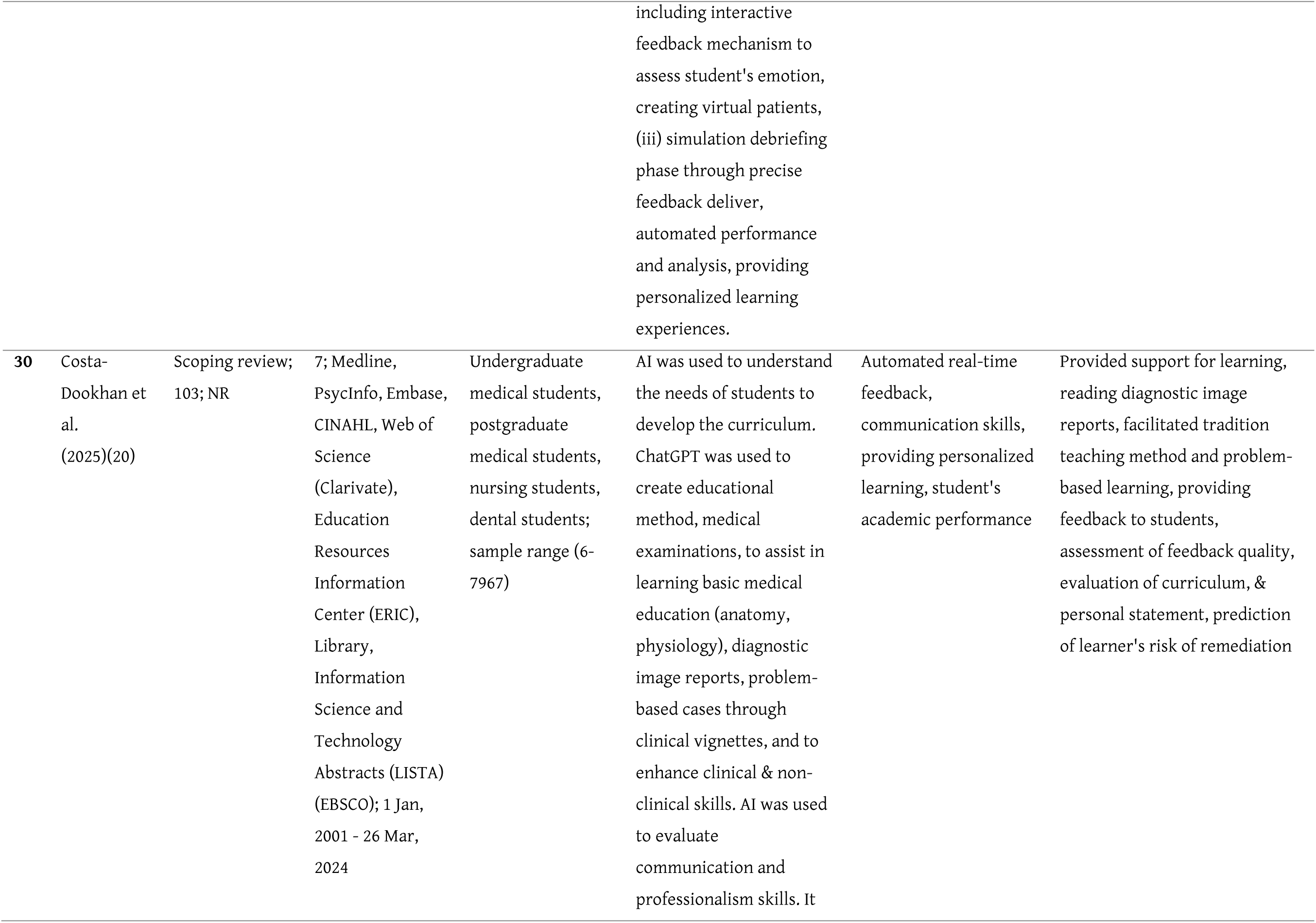
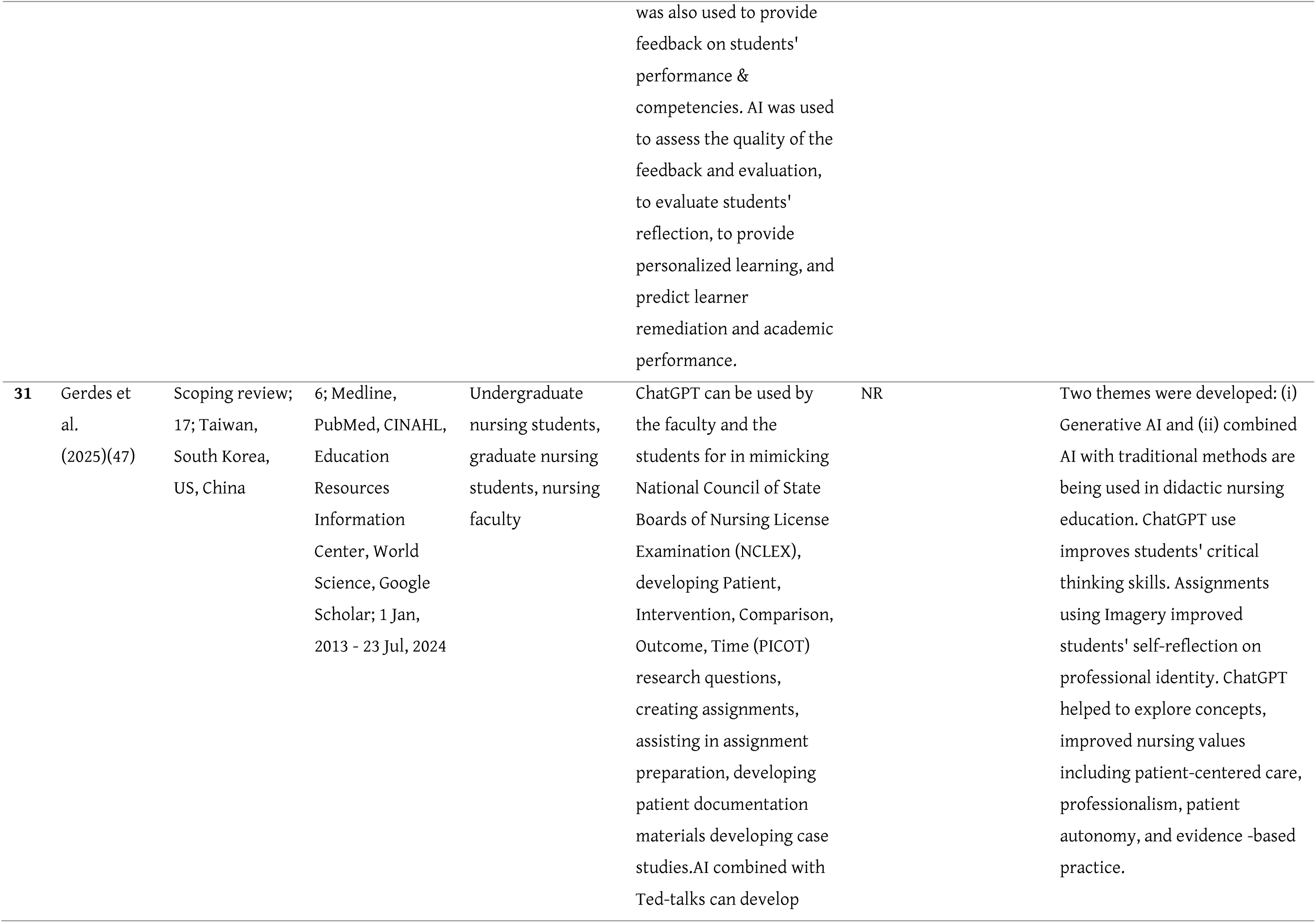
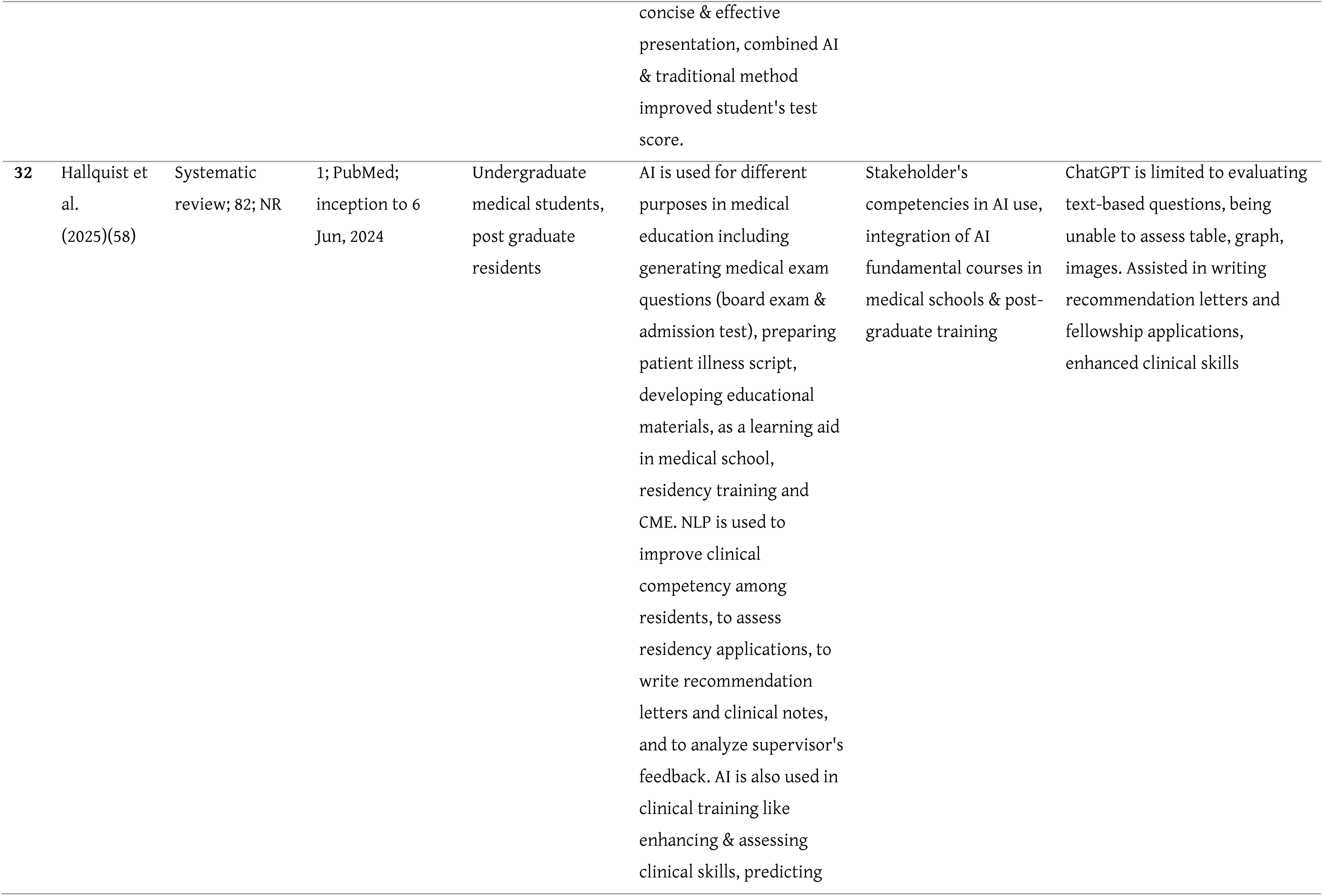
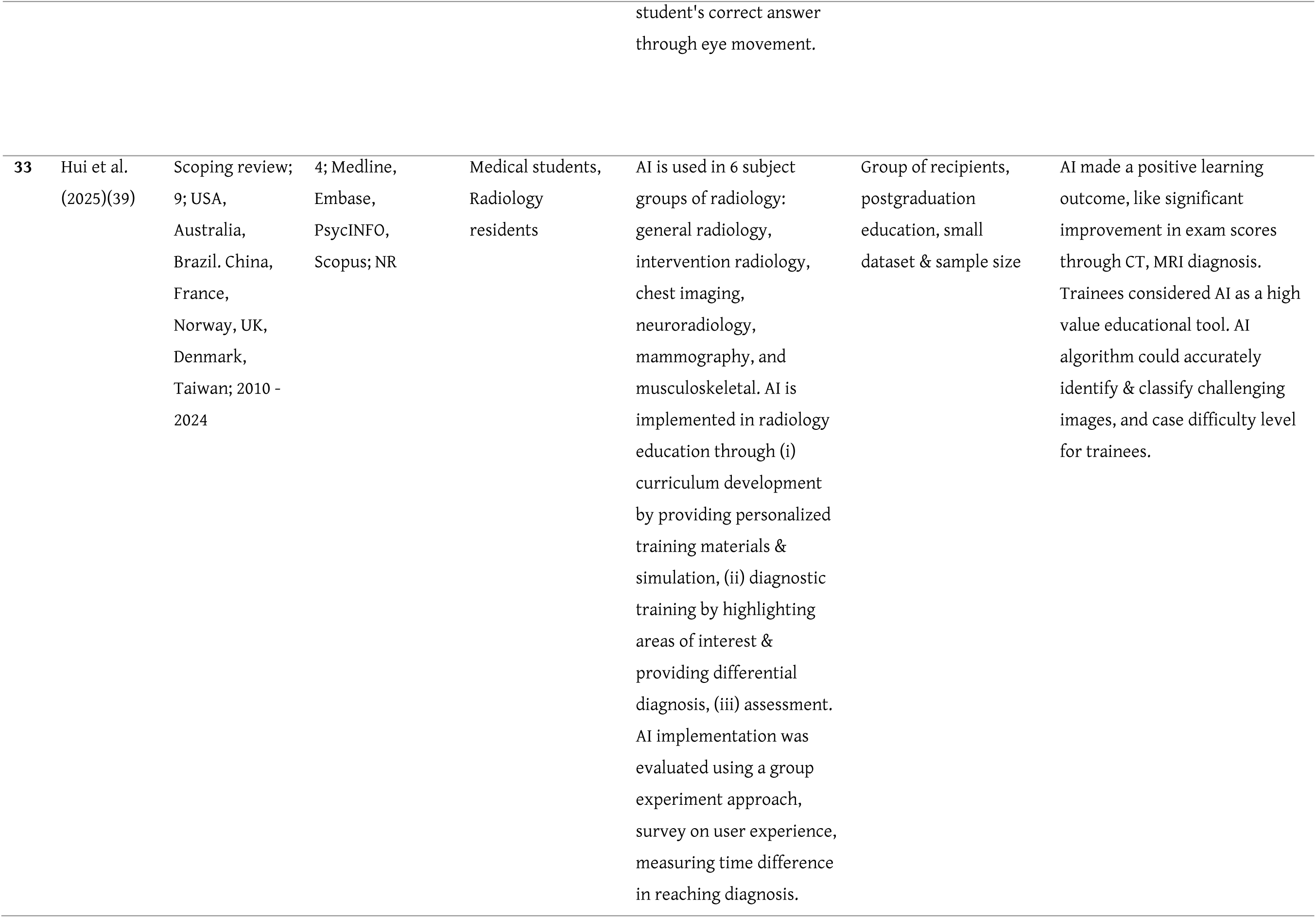
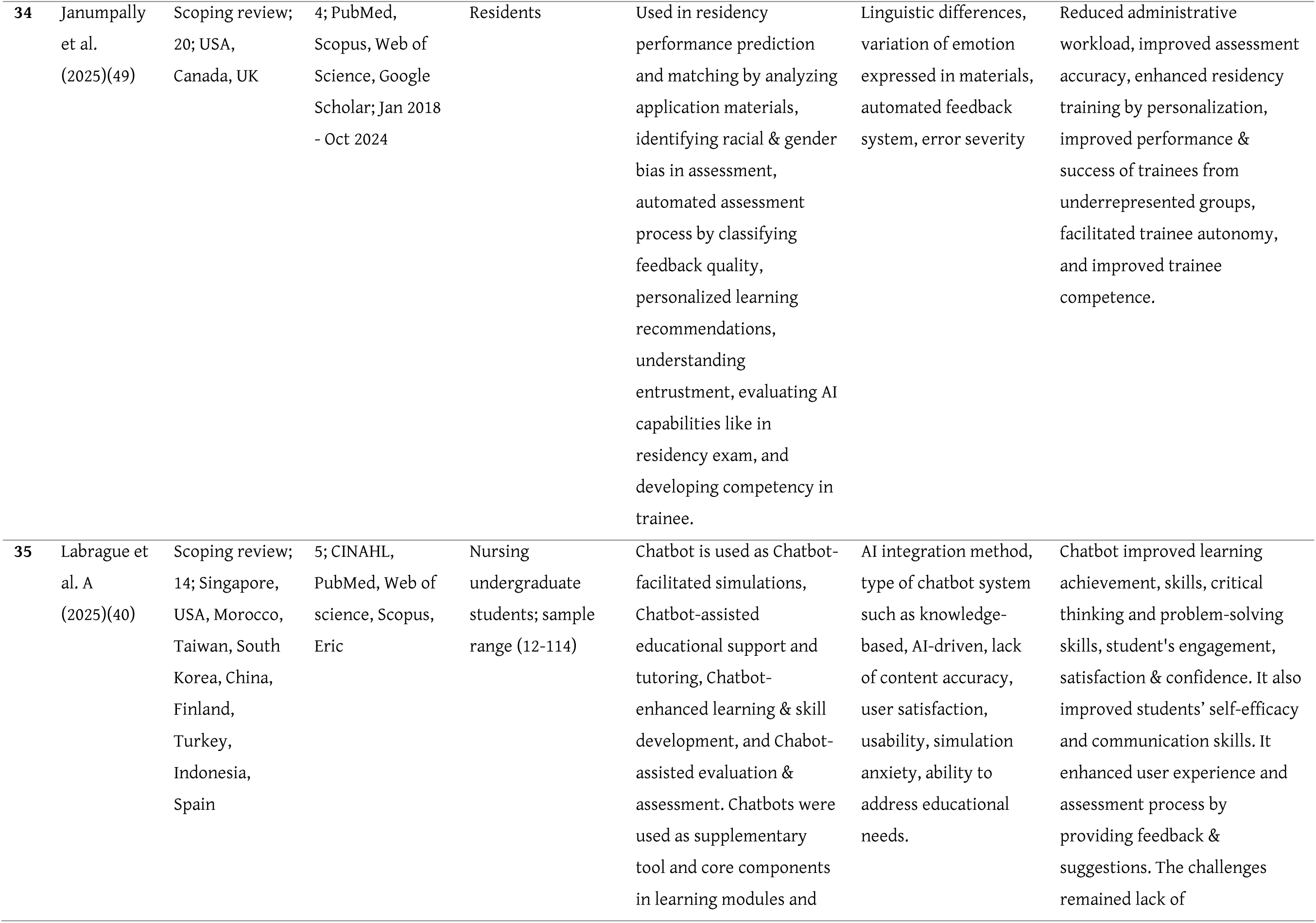
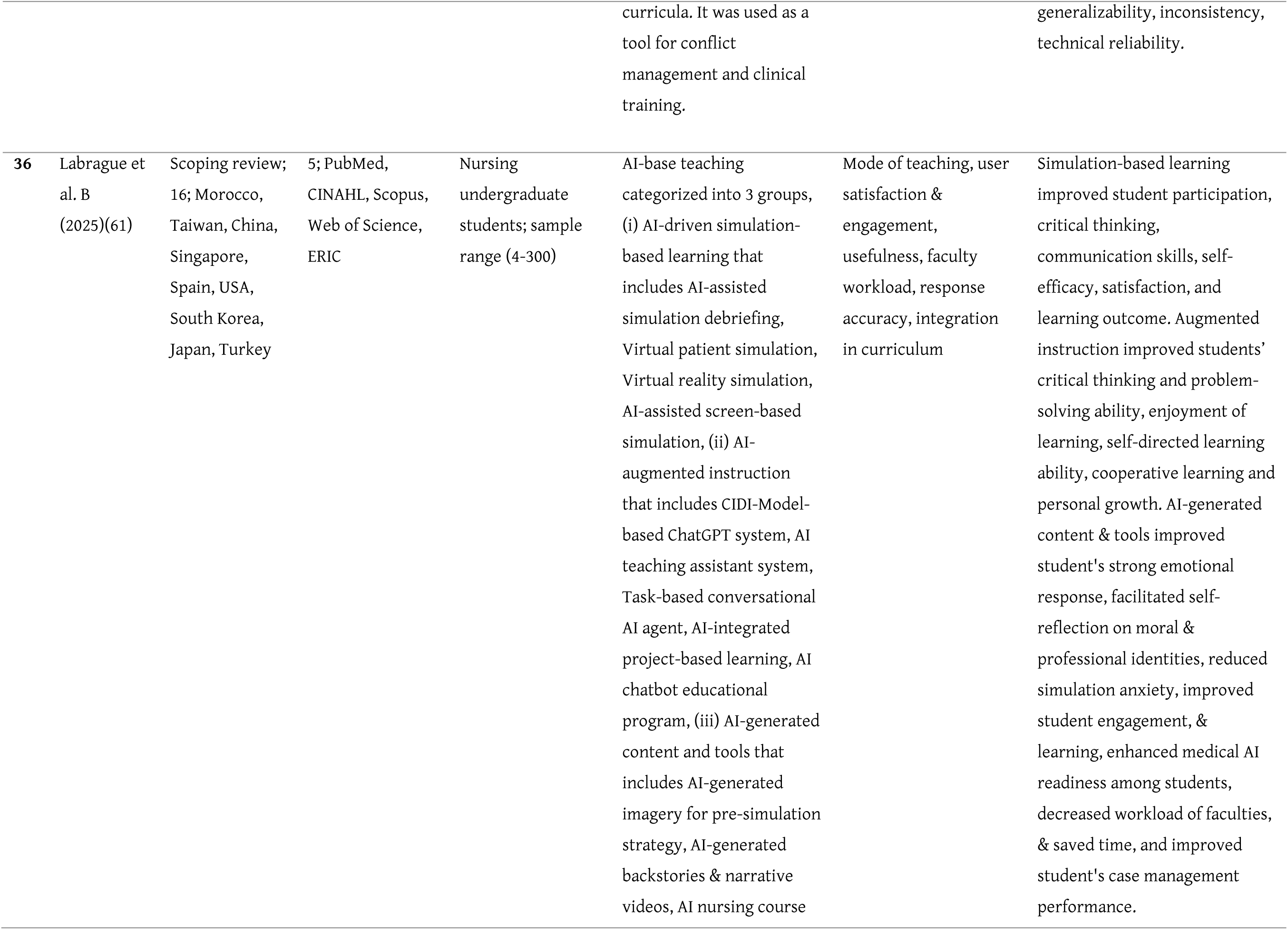
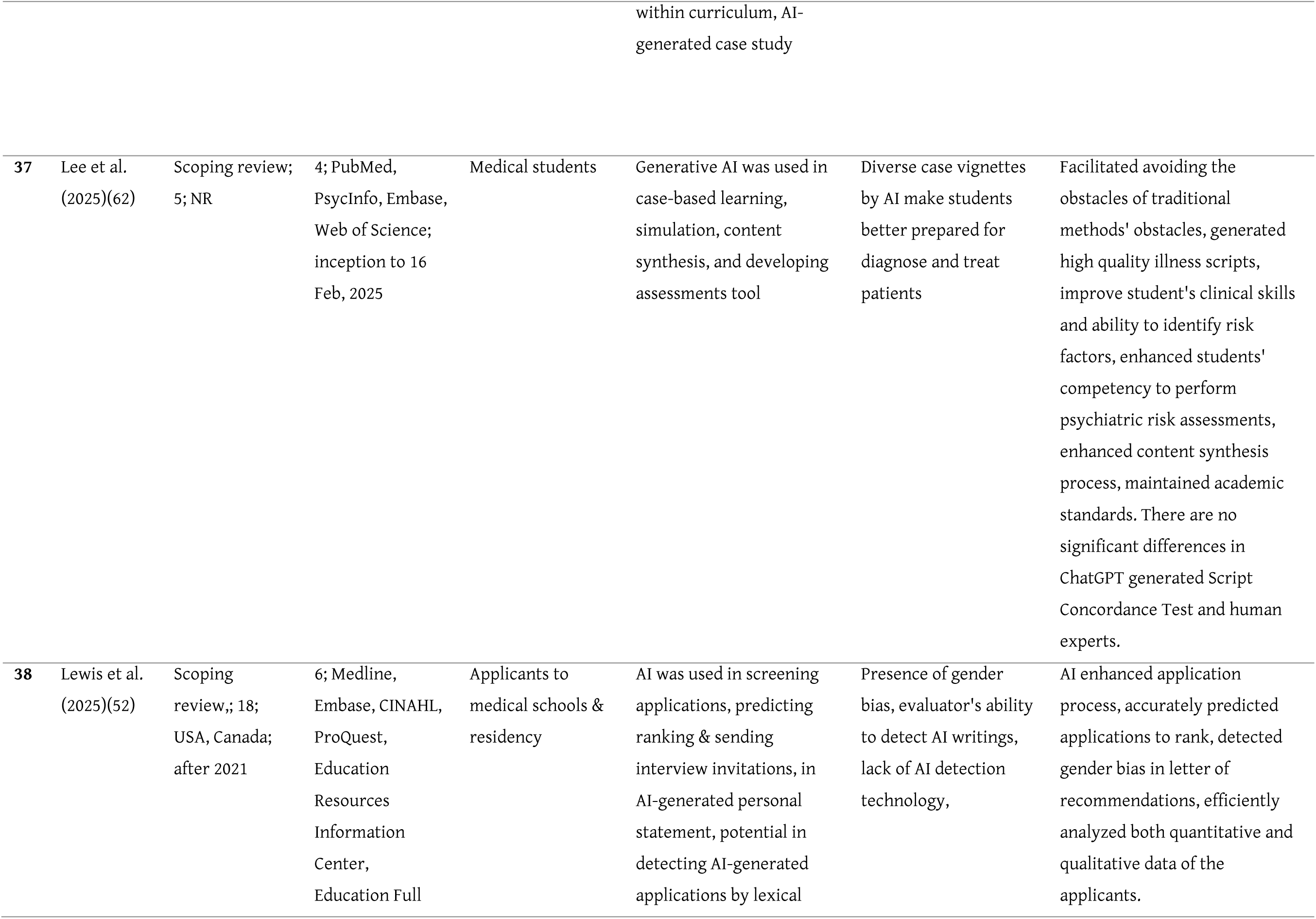
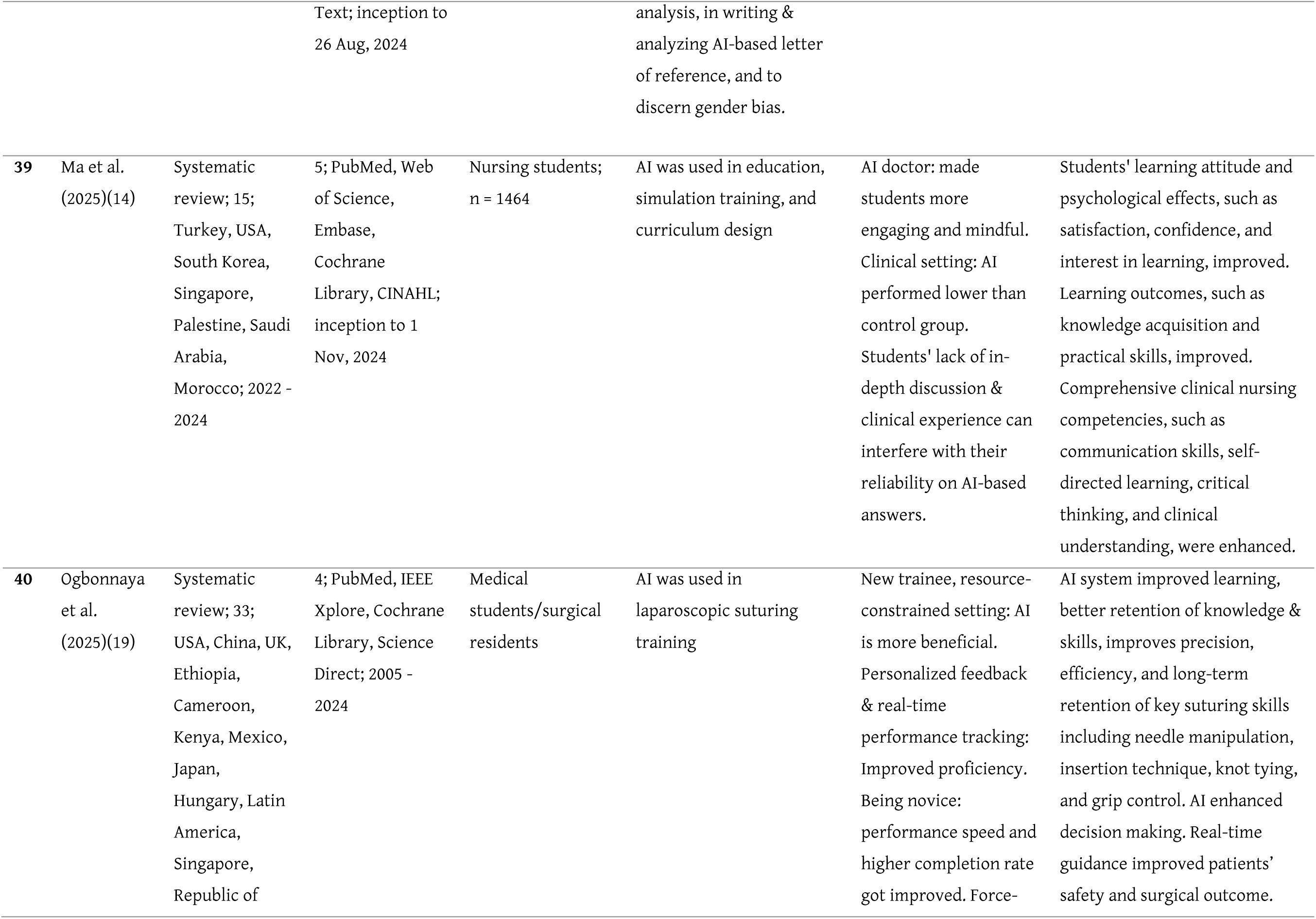
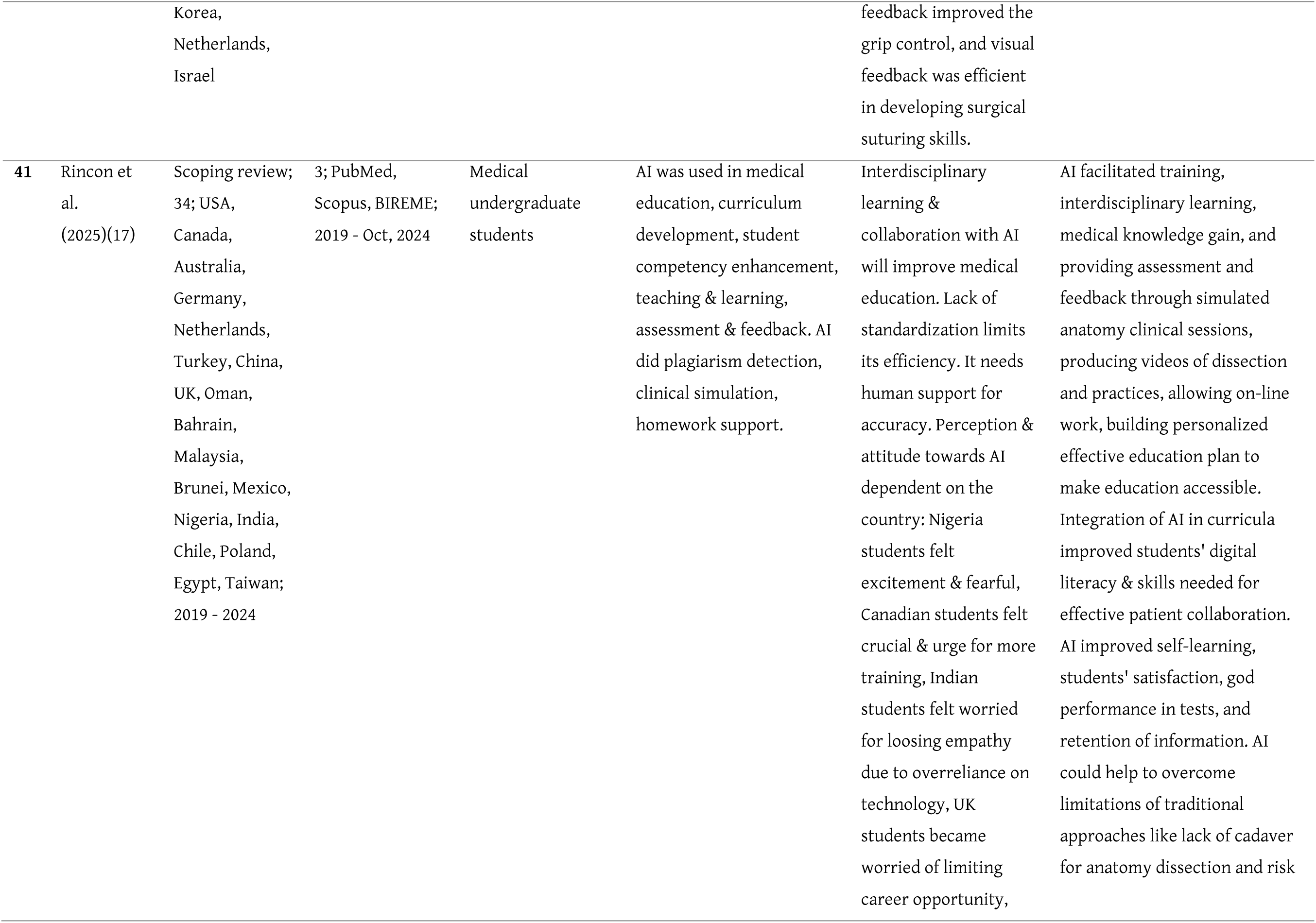
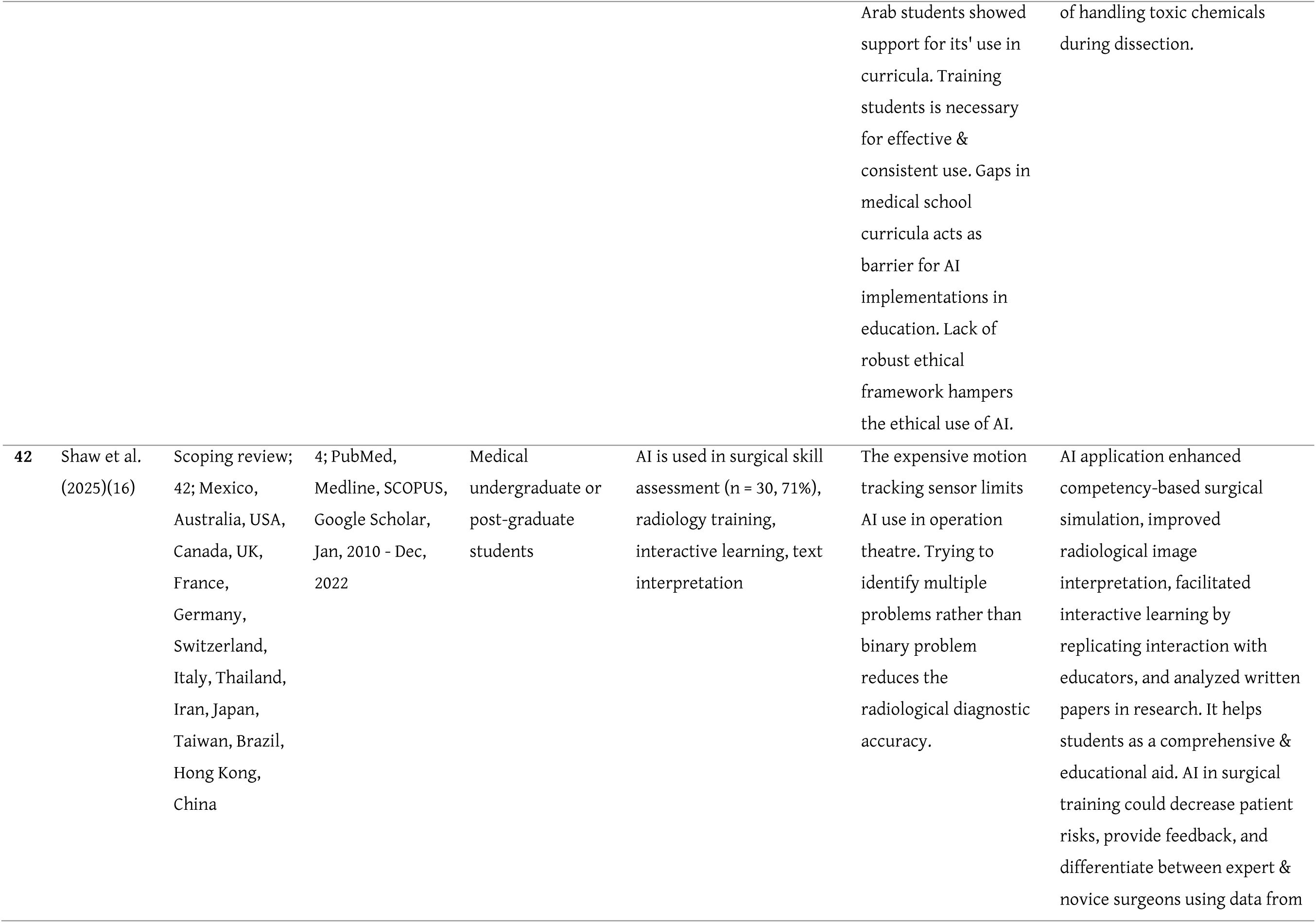
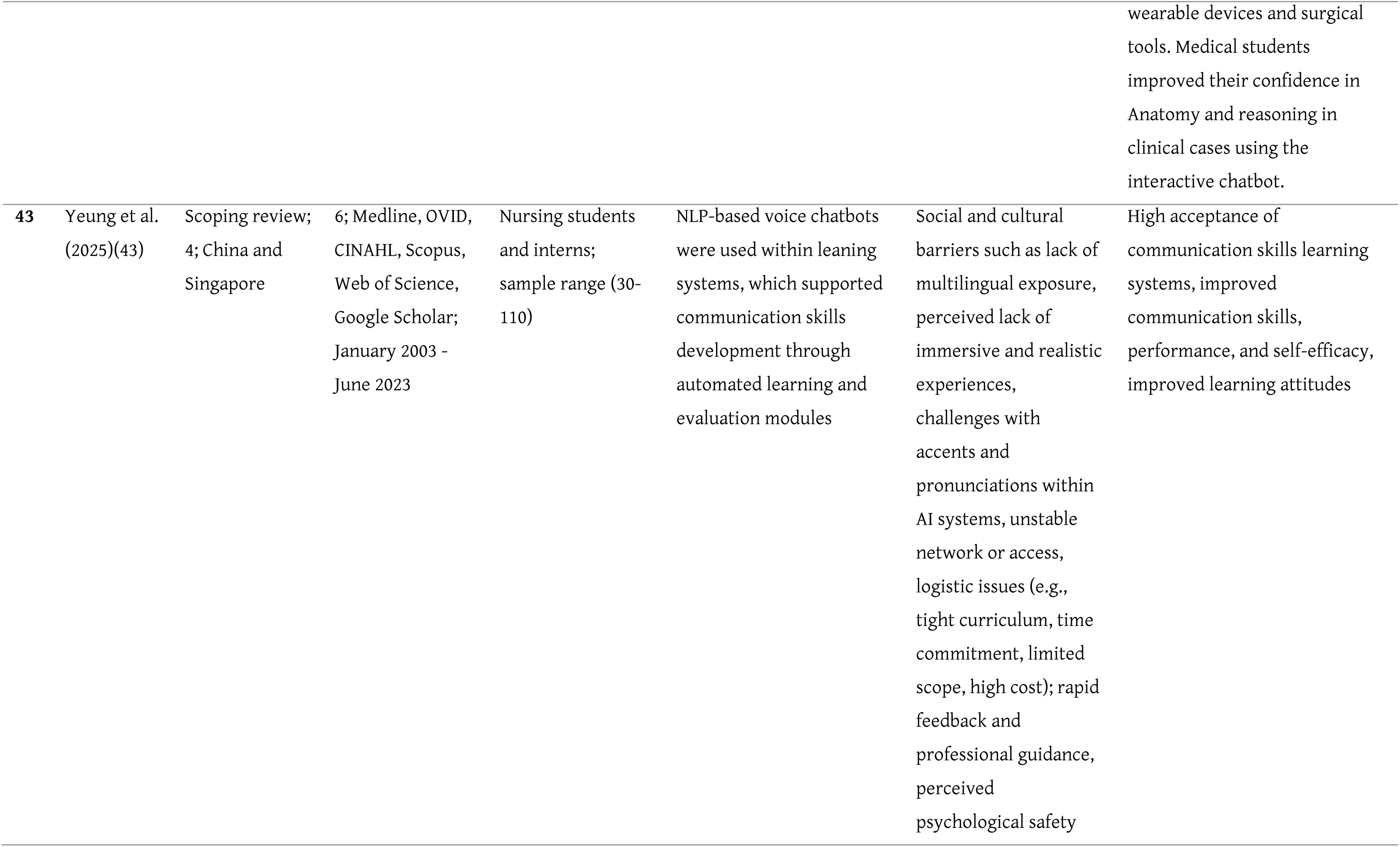
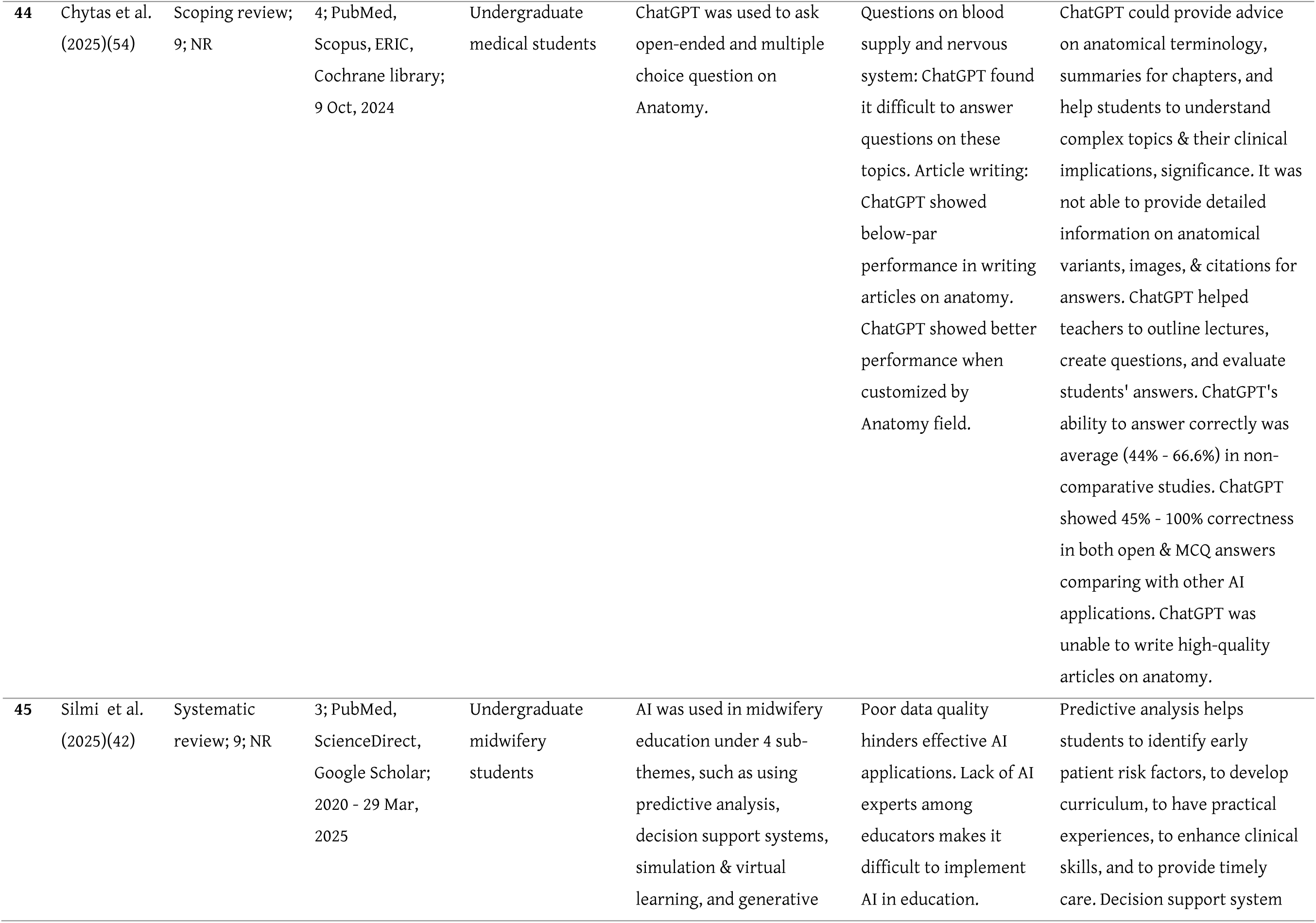
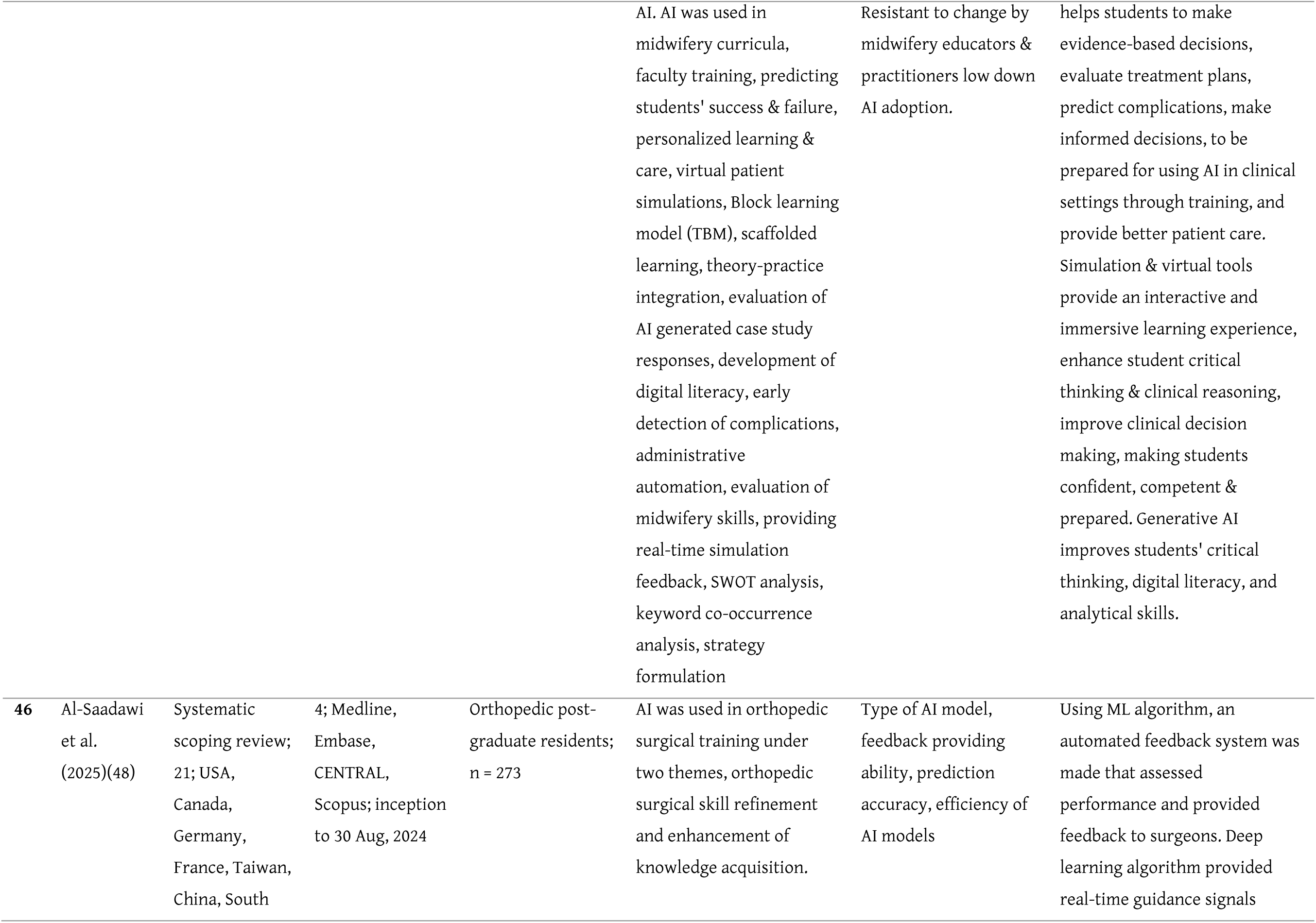
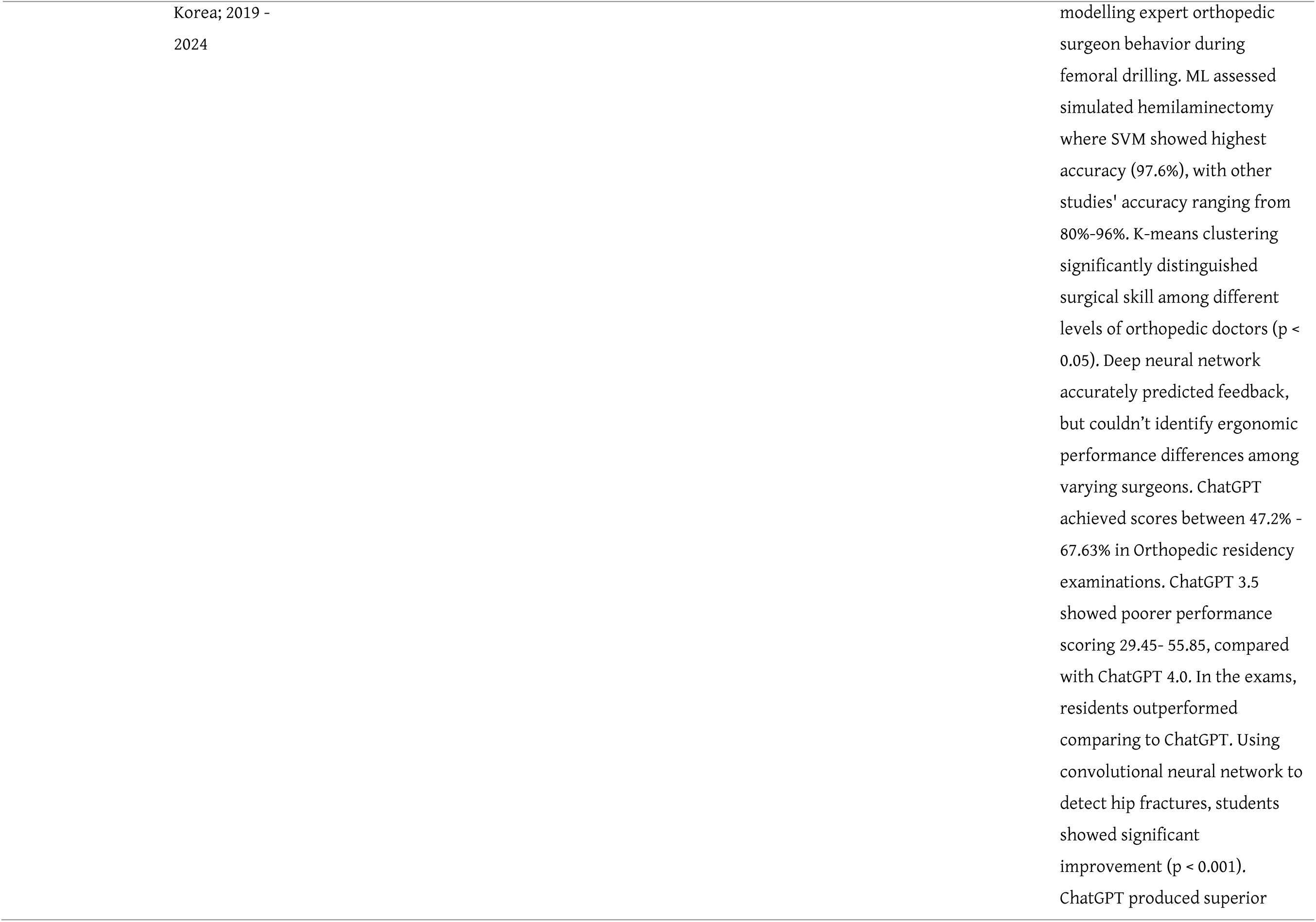
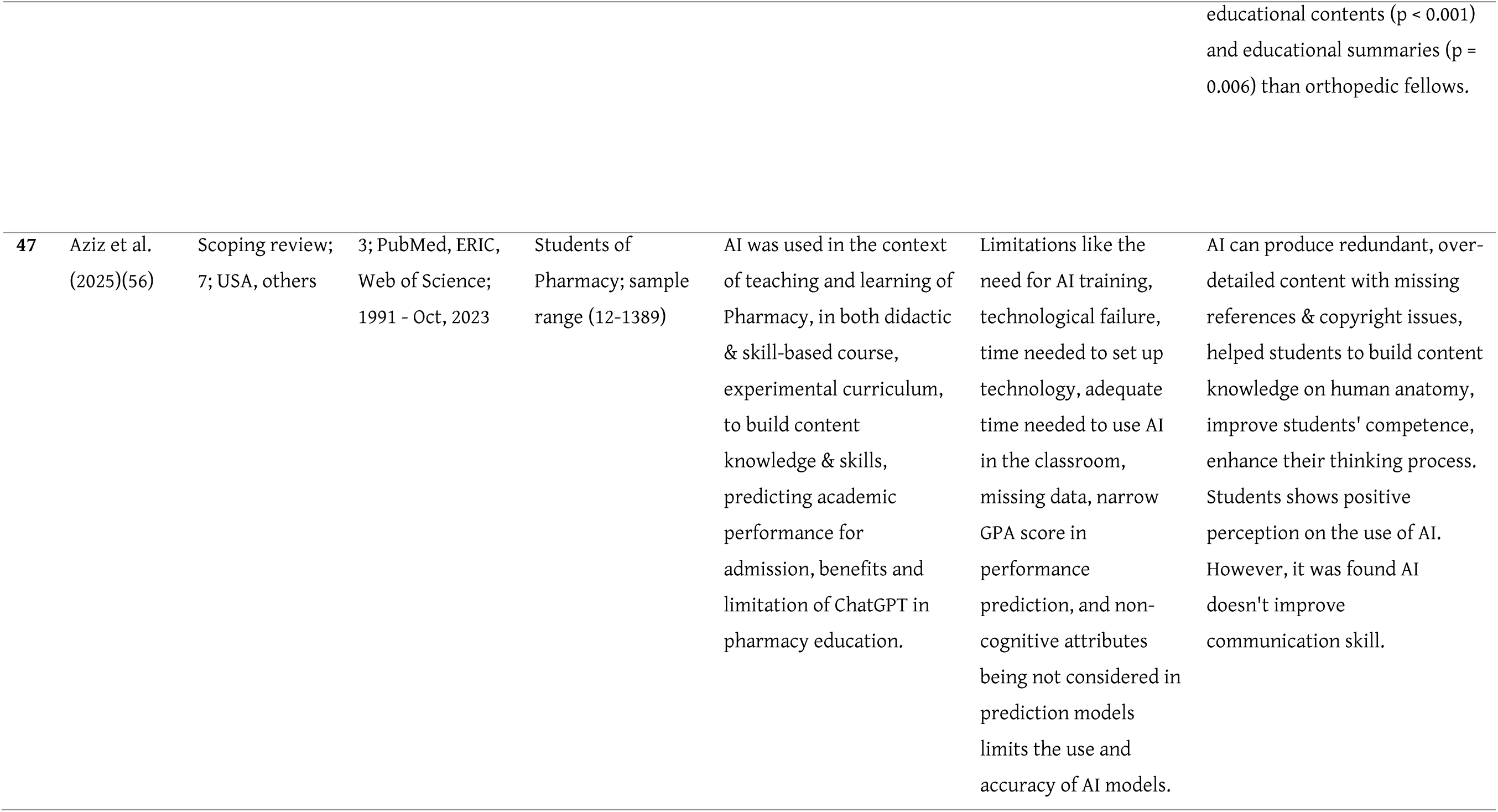
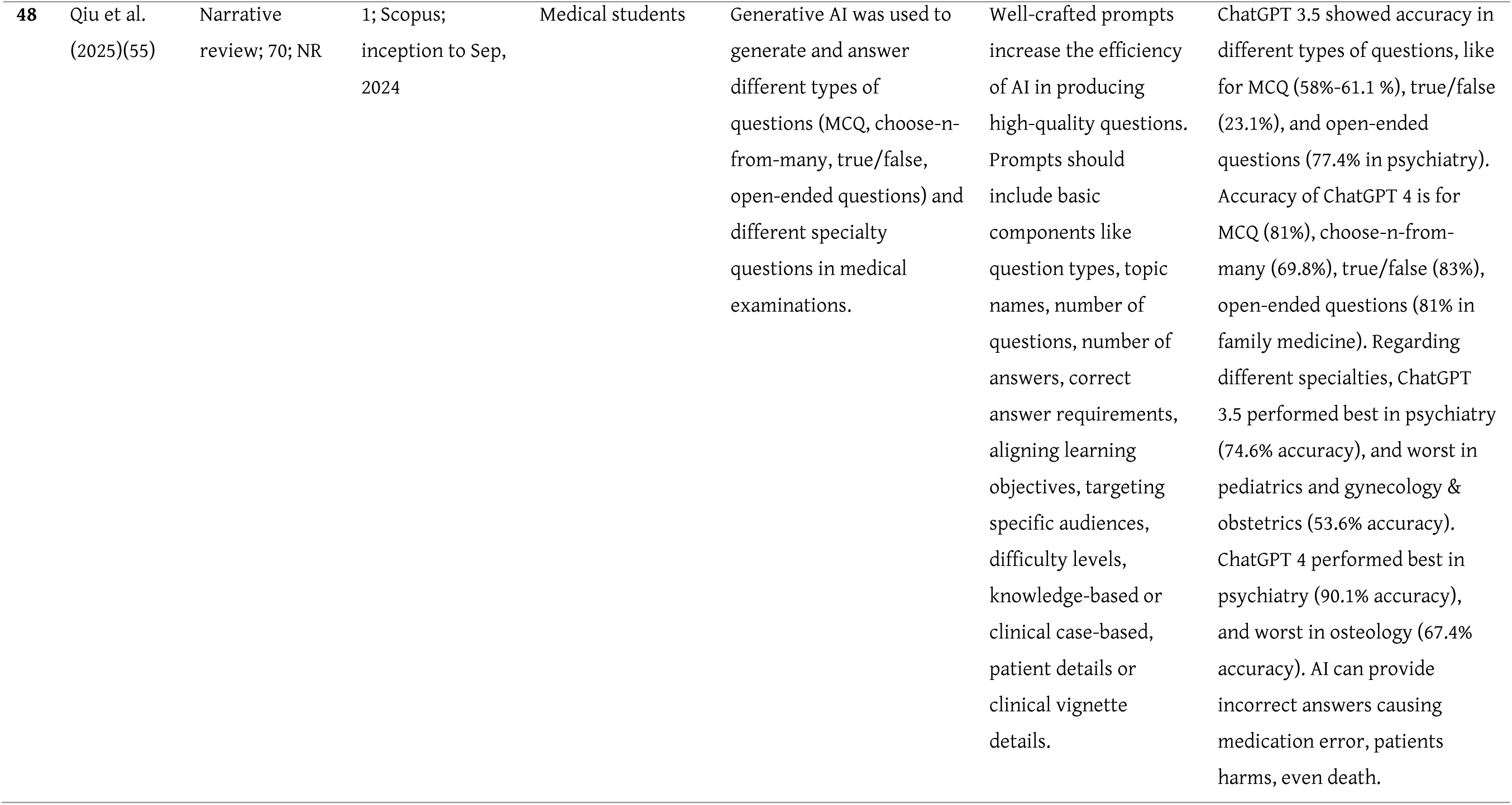
Summary of the included articles.

| No | Author<br>and year of<br>publication | Review type;<br>No. of<br>included<br>studies;<br>Countries of<br>primary<br>studies | No. of<br>databases;<br>Specific<br>databases used;<br>timeframe of<br>literature<br>search | Characteristics of<br>the study<br>participants and<br>sample size (if<br>reported) | AI applications and their<br>implementation | Reported factors<br>associated with AI<br>integration or use | Outcomes associated with AI<br>use in HPE |
| --- | --- | --- | --- | --- | --- | --- | --- |
| 1 | Chary et al.<br>(2018)(26) | Structured<br>literature<br>review; 13; Not<br>specified (NS) | 1; PubMed; Not<br>reported (NR) | Medical students<br>and residents<br>(focus on<br>analyzing their<br>clinical<br>documentation;<br>demographics not<br>aggregated) | NLP algorithms were<br>applied to learners'<br>written work to monitor<br>and provide feedback on<br>learning. For example, one<br>study used a program to<br>analyze daily student<br>notes for key clinical tasks.<br>Most interventions were<br>experimental and not yet<br>integrated into formal<br>curricula. | Automated analysis<br>removed subjective<br>bias in evaluating<br>students (software<br>focusing on content<br>rather than who wrote<br>it). However, many<br>studies lacked<br>discussion of<br>implementation (5<br>studies were excluded<br>for no implementation<br>details). | NLP tools demonstrated ability<br>to identify learning gaps and<br>give real-time feedback (e.g. an<br>“enhanced spell-checker”<br>improving students’ note<br>quality). They could also<br>quantify competency coverage<br>in evaluations (mapping text to<br>required competencies). Actual<br>improvements in learner<br>performance were not<br>measured in these studies. |
| 2 | Chan et al.<br>(2019)(37) | Integrative<br>review; 37<br>articles;<br>Singapore,<br>China, United<br>States, Canada, | 4; Medline, ERIC,<br>Education<br>Source, Web of<br>Science; up to<br>March, 2019 | Different learners'<br>groups, including<br>undergraduate,<br>postgraduate, and<br>continuing<br>education students | AI used mainly for<br>learning support, learning<br>assessment, and<br>curriculum review | Difficulties in<br>evaluating the<br>effectiveness of AI and<br>technical challenges<br>associated with<br>developing AI tools and<br>resources. Ability to | Enhanced feedback,<br>personalized learning, reduced<br>costs. Review of the curriculum,<br>improvement of student's<br>learning, enhanced student's<br>assessment, getting additional |
|  |  | Portugal,<br>Sweden |  |  |  | provide immediate<br>feedback, cost-<br>effectiveness, less time<br>consuming. | support for student's learning<br>experience. |
| 3 | Sapci et al.<br>(2020)(25) | Systematic<br>review; 26<br>articles; United<br>States, Canada,<br>Mexico,<br>Taiwan, France,<br>Portugal,<br>China,<br>Germany,<br>Korea,<br>Netherlands | 5; PubMed, IEEE,<br>ProQuest central,<br>Cumulative Index<br>of Nursing and<br>Allied Health<br>Literature<br>(CINAHL),<br>ScienceDirect;<br>1990–2020 | Medical and health<br>informatics<br>students | AI to enhance education<br>delivery and assessments;<br>building competencies<br>through AI education | Curricular and<br>competency gaps; Non-<br>standardized AI<br>curricula;<br>personalization and<br>feedback; necessary<br>competencies and<br>prevention of bias are<br>critical | Improved understanding of AI<br>in practice. AI enhanced<br>medical education such as<br>improved student's learning<br>experience, improved<br>assessment of the student's<br>academic performance,<br>personalizing learning process.<br>Virtual patient system<br>improved the efficiency of<br>medical education, facilitated<br>trainings in the absence of<br>human simulator. |
| 4 | Buchanan<br>et al.<br>(2021)(32) | Scoping review;<br>27 articles;<br>Canada and<br>other locations | 10; Medline,<br>CINAHL, Embase,<br>PsycINFO,<br>Cochrane<br>Database of<br>Systematic<br>Reviews,<br>Cochrane<br>Central,<br>Education<br>Resources<br>Information | Nurses, nursing<br>students, nurse<br>educators | AI integration in academic<br>and clinical nursing<br>education, including<br>simulation of interactions<br>supporting decision-making | Need for pedagogical<br>reform, lack of<br>educator readiness,<br>interdisciplinary<br>collaboration, and<br>technocentric cultural<br>competence | Support for safe, compassionate<br>care. Enhanced student's<br>comprehension,<br>communication, confidence and<br>self-efficacy, provide<br>personalized education,<br>improve critical-thinking &<br>clinical decision making,<br>improve readiness by using<br>simulation to manage real-life<br>problems. |
|  |  |  | Centre, Scopus, Web of Science, and Proquest; up to April 2020 |  |  |  |  |
| 5 | Charow et al. (2021)(9) | Scoping review; 41 articles; United States, Canada, France, Mexico | 8; MEDLINE, Embase, PsycINFO, Emcare Nursing, Two Cochrane Databases, ERIC, and Web of Science; until July 2020 | Undergraduate and postgraduate medical students, continuing education learners | Competency-based training across cognitive, psychomotor, affective domains | Low AI literacy, regulatory barriers, lack of funding. Interfaculty collaboration, support of regulatory bodies, lack of AI literacy among faculty for designing curricula, infrastructural barrier for AI integration. | Improved AI-readiness in HCPs. Improved student's satisfaction with AI course resources, perception of clinical relevance, educational activities. Improved resident's complex problem solving skill, ability to analyze large data, and articulating & interpreting results using multiple resources. |
| 6 | Lee et al. (2021)(29) | Scoping review; 22 articles; USA, Canada, South Korea, France, India, Netherlands, New Zealand, Oman, Pakistan, Spain | 7; Medline, Embase, PubMed, Scopus, ERIC, MedEdPortal, Cochrane; Jan 2000 – July 2020 | Undergraduate medical students | Recommendations for curriculum and delivery strategies | Lack of consensus on content/delivery, faculty's lack of expertise on AI, faculty resistance, lack of AI accreditation, licensing requirements, limited curricular hours, lack of AI core competencies, lack of evidence on AI impact on medical education, ability to operate AI tools, ability to communicate AI results. | Need for AI competencies in UME. Helps students to integrate AI in clinical-reasoning. |
| 7 | Grunhut et al. (2021)(57) | Integrative review; 39 ; NR | 4; PubMed, Medline, Google Scholar, Web of Science; 2010–2020 | Primarily undergraduate medical students (five survey studies on student attitudes) and some physicians (in a few mixed surveys); many papers were commentaries with no participants | Two studies reported pilot AI curriculum changes. Most literature consisted of calls or plans to incorporate AI into courses. The review notes that medical schools have no unified approach yet and urges a collaborative, longitudinal strategy to embed AI training | Medical students showed high interest in AI and recognized its importance (positive driver), but institutions face inertia. Lack of faculty expertise and crowded curricula are barriers. The need for faculty development and defined AI competencies is highlighted. | No direct educational outcomes measured (since implementations were scarce). The review mostly reported perceptions – e.g. students feeling unprepared in AI and wanting formal training. No evidence yet of improved knowledge or skills, underscoring an evidence gap. |
| 8 | Kiruburajan et al. (2021)(44) | Scoping review; 49 ; 17 countries (mostly North America and Europe); studies published 2007–2021 | 8; Medline, ERIC, Embase, CINAHL, Web of Science, Compendex, Scopus, and IEEE Xplore; up to Apr 13, 2021 | Surgical trainees (medical/dental students, residents, fellows); n = 1185 | Most studies were proof-of-concept validations rather than integrated programs. 36 studies used retrospective or simulated trainee data., and only 5 were tested real-time in the operating room. AI systems were typically used to assess or tutor surgical skills outside formal curricula. Implementation into actual training was minimal, with few providing feedback to trainees (focus was on algorithm accuracy). | High accuracy of AI in objective skill assessment was noted (many studies reported AI could classify or score surgical performance with high fidelity). | AI systems showed promise in evaluating and potentially improving surgical training. For example, AI models could objectively grade surgical technique with high agreement to expert judgment. Some interventions personalized training or identified struggling trainees based on performance data. |
| 9 | Yanik et al. (2022)(45) | Systematic review;25; USA | 1; Google scholar; Nov 2013-Jun 2020 | Surgeons (novice, intermediate, expert); varying experience levels (self-reported hours) | DNNs process kinematic/video data to classify skill levels; end-to-end models (e.g., CNN) directly assess performance; visual feedback via CAM highlights key performance areas. | Dataset availability: Large, annotated datasets enable effective DNN training. Data type: Kinematic data easier to process but invasive; video data non-invasive but requires preprocessing. Model choice: CNNs outperform RNNs/FCNs due to suitability for surgical data. | High accuracy: DNNs achieve up to 98.7% accuracy in skill classification. Visual feedback: CAM provides actionable performance insights. Objective assessment: Reduces subjectivity compared to traditional methods. |
| 10 | Preiksaitis et al. (2023)(33) | Scoping review; 41 articles; United States, Taiwan, Korea, European countries (e.g., Germany, Netherlands, France) | 3; PubMed, Web of Science, Google Scholar; Jan 2022 – Jun 2023 | Medical students, educators, learners | Applications in feedback, quiz generation, self-directed learning plans, generating novel learning materials, writing support, assisting in research | Academic integrity, critical evaluation skills, evolving assessments | Enhanced test prep, customized learning experience, providing tailored feedback, improved communication training |
| 11 | Pupic et al.<br>(2023)(46) | Systematic review;34; Canada, UK, USA, Denmark, Saudi Arabia, and others | 6; Medline, Embase, CINAHL, ERIC, NCBI, and Web of Science; till April 23, 2022 | Medical students, professionals; 54% female, 46% male; low AI knowledge; sample range (62-1492) | AI taught via workshops, extracurricular programs; focus on ethics, theory, and communication for clinical use. | Financial barriers: Limit curriculum development. Educator resistance: Slows adoption. Lack of AI-experienced staff: Hinders teaching. Limited digitalization: Restricts infrastructure. | Improved competency: Enhances clinical decision-making. Increased confidence: Better AI tool usage. Positive attitudes: 81–92% support AI curriculum. |
| 12 | Varma et al.<br>(2023)(36) | Systematic review;11; US and UK | 3; PubMed, Web of Science, EBSCO; till March, 2021 | Undergraduate medical students | AI used for trend spotting, teaching via virtual cases, and assessing clinical notes; implemented as one-off sessions, not integrated long-term. | Financial burden: High costs limit adoption. Technical expertise: Requires collaboration with data scientists. Training needs: Educators need AI tool training. | High accuracy: Up to 94.7% in teaching tasks. Improved performance: 70% enhancement in echocardiography assessment. Student satisfaction: Positive feedback from virtual tools. |
| 13 | Weidener et al.<br>(2023)(41) | Scoping review;12; US | 4; PubMed, Embase, Scopus, and Web of Science; 2000-June 2023 | Medical students | AI ethics taught via case-based learning, small group seminars; integrated into existing ethics modules or clinical years. | Lack of faculty expertise: Limits teaching capacity. Overloaded curricula: No dedicated time for AI ethics. Lack of standards: Hinders content development. | Enhanced preparedness: Equips students for ethical challenges. Improved understanding: Fosters fairness and transparency in AI use. |
| 14 | Furey et al. (2024)(23) | scoping review; 13; Qatar, Canada, USA, Singapore, Caribbea, Jordan, Hongkong | 4; until Sept 11, 2023 | Undergraduate and postgraduate students | Teaching and assessment design, student implementation, academic and scholarly support and clinical and practical application | Digital literacy skills, competencies individual strength, tailored action plans | Educators: supportive role in curriculum design and assessment creation<br>Students: enhanced digital literacy, supportive in academic writing and scholarly research clinical setting: enhanced practical skill in terms of both educator and student perspectives |
| 15 | Artsi et al. (2024)(51) | Systematic review;8; USA | 1; PubMed; 2023 |  | LLMs generated MCQs for medical exams; used prompts referencing textbooks, journals, or prior exams; some required re-prompting for clinical context. | Prompt quality: Specific prompts improve MCQ validity. Training needs: Educators require AI literacy. Resource demands: Implementation needs investment. | Valid MCQs: 40–66.6% suitable for exams. Time efficiency: AI generated 50 MCQs in 21 minutes vs. 211 minutes for humans. Limited quality: Human-written MCQs scored higher. |
| 16 | Gordon et al. (2024)(50) | Scoping review;278; USA, UK, Germany | 3; PubMed, Embase, and MedEdPublish | Medical students, trainees, educators | AI used for MCQ generation, virtual case learning, and skill assessment; integrated via workshops, electives, and case-based learning. | Faculty expertise: Limited AI knowledge hinders teaching. Curriculum overload: No space for AI content. Cost: High implementation expenses. | Enhanced learning: Improved engagement via virtual cases. Time efficiency: Faster MCQ generation. Positive perceptions: Students support AI integration. |
| 17 | Isaac et al. (2024)(30) | Scoping review;4; Germany, Japan, Canada, USA | 5; PubMed, Embase, Scopus, Cochrane Central Register of Controlled Trials, and the Healthcare Administration Database; till May 21, 2023 | Surgical trainees, practicing surgeons; | AI/ML used for video-based surgical skill assessment; CNNs analyzed laparoscopic and cataract surgery videos, focusing on tool detection and motion tracking; no feedback provided to trainees. | Lack of user acceptance studies: Hinders adoption. Limited implementation research: Restricts practical use. Time constraints: Manual review barriers not fully addressed. | High accuracy: 87–94.1% in skill assessment. Reliable feedback potential: AUROC up to 0.989 for skill classification. Limited practical impact: No subject performance improvement evaluated. |
| 18 | Kranz et al. (2024)(27) | Scoping review;6; UK, Hong Kong, Ireland, Saudi Arabia, Egypt | 2; PubMed, Academic search premier; 2021-May 2024 | Midwifery students, educators | AI integrated via curriculum updates, personalized learning materials, and educator training; focus on ethical, legal, and technical AI dimensions. | Educators' training needs: Lack of AI expertise. Data quality: Limited high-quality midwifery data. Academic integrity: Risks of plagiarism with AI tools. | Personalized education and feedback. Enhanced efficiency: More time for patient care. Increased acceptance: Potential for midwives to lead AI initiatives. |
| 19 | Li et al. (2024)(28) | Meta-ethnographic synthesis; 21; USA, Australia, China, England, Germany | 5; PsycINFO, CINAHL, Scopus, PubMed, and Web of Science; Till Feb 2024 | Medical students, educators, clinical practitioners; | AI integrated via virtual patient simulations, personalized learning, and question generation; limited formal curriculum inclusion, often as electives. | Hinders AI teaching. Time constraints: Prioritization of core medical skills. Cultural barriers: Varying AI adoption across countries. | Improved learning engagement. Increased efficiency: Faster question generation and data processing. Improved clinical skills: Safe virtual practice environments. |
| 20 | Lifshits et al. (2024)(13) | Scoping review;15; Australia, South Korea, Taiwan, Singapore, Croatia, Spain, Turkey | 2; PubMed, ScienceDirect; up to 2023 | Nursing students, novice nurses, faculty specialists, practicing nurses; mostly undergraduate and postgraduate students; sample range (15-336) | AI tools like VR and AR were used in simulations to enhance clinical skills, critical thinking, and communication; implemented through interactive games, virtual patient scenarios, and 3D learning environments integrated into nursing curricula. | Motivation: Positive attitudes and engaging tools like robots increased learning motivation. Usability: Medium usability scores; technical difficulties hindered effective use. Feedback: Lack of instant feedback reduced learning effectiveness. Language barriers: Non-native language systems caused discomfort and reduced accessibility. Cultural relevance: Non-local patient scenarios limited applicability. | VR and AR enhanced satisfaction, engagement, and critical thinking. AI groups showed higher knowledge scores in sepsis care and leg ulcer care. VR/AR enabled practicing facilitated effective communication. Simulations improved confidence in clinical skills. |
| 21 | Lucas et al. (2024)(35) | Systematic review;40; USA, Poland, Canada, Korea, Europe | 3; PubMed, Web of science, Embase; till Feb, 2024 | Medical students, resident physicians; primarily trainees in various medical disciplines | LLMs used for exam preparation, clinical scenario simulations, case report generation, and personalized learning; implemented via interactive question- answering, curriculum design, and clinical decision support tools. | Accuracy: High exam performance but occasional inaccuracies require verification. Accessibility: Instant access to knowledge enhances learning efficiency.<br>Bias: Training data biases may perpetuate discrimination in education.<br>Overreliance: Risk of reduced critical thinking due to dependence on LLMs. | LLMs passed USMLE, ophthalmology, and surgical board exams. Simulations supported hypothesis generation and decision-making. |
| 22 | Silva et al. (2024)(31) | Scoping review;17; NR | 5; PubMed, Scopus, SciELO, Embase, Web of Science, Google Scholar; NS | Medical students, surgical residents, fellows, attending surgeons, nurses; mixed experience levels, primarily trainees; sample range (8-74) | AI used in virtual simulations for practicing laparoscopic, suturing, and knot-tying skills; provided real-time feedback and performance analysis in controlled training environments. | Data security: Sensitive patient data use requires strict privacy policies.<br>Resistance to adoption: Surgeons and educators fear AI may replace roles or be unreliable.<br>Curriculum integration: Lack of AI-specific training in curricula hinders effective use. | Simulations improved laparoscopic and suturing proficiency. AI tailored feedback to individual trainee needs.<br>Practice in virtual environments reduced patient risks.<br>Automated systems provided accurate skill evaluations. |
| 23 | Tozsin et al. (2024)(15) | Systematic review; 36; SA, Canada, UK, Ireland, Lebanon, Croatia, | 3; PubMed, Embase, Cochrane Library; 2000-March 2023 | Medical students, residents, radiologists, clinical psychology students, healthcare | AI tools like SVM and NN used for surgical skill assessment, disease diagnosis, and student performance evaluation; implemented via | Knowledge gap: Limited AI understanding among students hinders curriculum integration. | Improved surgical skills: AI-assisted training enhanced suturing and knot-tying proficiency.<br>Enhanced diagnostic accuracy: AI models achieved over 80% |
|  |  | Singapore,<br>China,<br>Australia,<br>Turkey |  | students; sample<br>range (15-3018) | simulations, automated<br>grading systems, and real-time feedback in training. |  | accuracy in diagnosing acute<br>abdominal pain.<br>Better student assessment:<br>Automated systems accurately<br>evaluated clinical competencies.<br>Increased confidence: AI<br>feedback improved trainee<br>confidence in technical skills. |
| 24 | Xu et al.<br>(2024)(38) | Scoping review;<br>113; NR | 4; PubMed,<br>Medline, Embase,<br>Web of Science; 1<br>Jan, 2022 - 30<br>Nov, 2023 | NR | Providing personalized<br>learning, tailored<br>feedback, assisting in<br>writing & research,<br>curricula development,<br>innovative teaching<br>planning, medical exam<br>performance &<br>preparation | Usability, accessibility,<br>interactive capabilities,<br>ethical concerns, data<br>privacy, student-<br>teacher-patients<br>privacy, accuracy,<br>reliability, lack of<br>specific guidelines for<br>AI use | Benefits of ChatGPT use<br>included enhancing new<br>learning methods through<br>developing customized learning<br>plans, assisting with writing<br>research paper, quickly saving<br>time, improve teaching quality<br>through simulating real-world<br>clinical practice scenarios,<br>providing supplementary<br>teaching tools, and helping to<br>prepare for medical exams. |
| 25 | Zhang et al.<br>(2024)(24) | Systematic<br>review; 42;<br>USA, China,<br>Canada, UK,<br>Italy, Australia,<br>Germany,<br>Brazil, and<br>others | 4; Web of<br>Science, PubMed,<br>Scopus, IEEE<br>Xplore; 1 Jan,<br>1990- 31 Dec,<br>2022 | Undergraduate<br>medical students,<br>specialized<br>trainers, continued<br>medical students | AI is used in three stages of<br>medical teaching, (i)<br>teaching implementation<br>in basic & clinical<br>education-like anatomy,<br>pathology, ECG<br>interpretation & diagnosis,<br>surgical training<br>feedback,(ii) teaching evaluation by predicting student's performance, automated scoring of exams, (iii) teaching feedback on the quality of teaching and medical curriculum. | Technical limitations,<br>interactivity, ability to<br>provide real-time<br>feedback, effectiveness<br>& reproducibility, lack<br>of prior knowledge,<br>data privacy &<br>protection, generalization ability, black box nature | AI was more effective in<br>teaching than traditional<br>methods, assessing virtual<br>surgical skills, and users found<br>it useful & usable. |
| 26 | Adarkwah et al. (2025)(22) | Scoping review; 33; Argentina, Mexico, Columbia, Australia, Austria, Canada, China, Cyprus, Germany, India, Italy, Jordan, Korea, Pakistan, Republic of Korea, Saudi Arabia, Switzerland, Taiwan, Turkey, Ukraine, USA, Yemen | 6; Web of Science, PubMed, CINAHL, Scopus, Science direct, Google Scholar; 2023 - 2024 | NR | Challenges of AI implementation include variation in quality and availability of training data, resulting in bias, diverse cultural and regional healthcare practices, influence of larger economy on AI training & infrastructure, subscription-based model widening digital gaps among urban-rural and socioeconomic groups, global internet disparities, ensuring accuracy & quality of provided information, providing generalized feedback, lack of accountability for misinformation, over reliance on AI. | Multimedia integration, ChatGPT improves participants' practical learning experience for this. Multilingual property, ChatGPT enhances cultural inclusiveness for this and becomes accessible for people from underserved communities. Continuous availability, this property addresses time limitation and logistic issues. Accessibility during non-traditional hours helps to balance learning with other responsibilities. | ChatGPT helps to understand complex medical concepts, addresses the lack of skilled instructors, provides a personalized learning plan, provides practical training, and enhances communication skills by simulating clinical scenarios with virtual patients. It can help in curriculum development, evaluate student's papers effectively for clarity and grammatical correctness, and provide feedback. ChatGPT can operate accurate health documentation and enhance health literacy. |
| 27 | Aster et al.<br>(2025)(53) | Scoping review;<br>145; USA, India,<br>Germany,<br>Japan, France,<br>Egypt, Greece,<br>and others | 5; ACM Digital<br>Library, IEEE<br>XPLORE, JSTOR,<br>Web of Science,<br>PubMed; till Nov,<br>2023 | Medical students,<br>nursing students | In most studies, ChatGPT<br>was used to show its<br>capability to pass medical<br>exams and answer board<br>exams where ChatGPT 4.0<br>did better than ChatGPT<br>3.5. ChatGPT was also used<br>to develop scenario cases<br>for medical education.<br>Pitfalls of using ChatGPT<br>are the possibility of being<br>used to cheat, while the<br>advantages are developing<br>exams, evaluating papers,<br>providing feedback,<br>providing a personalized<br>training plan, and helping<br>in summarizing articles.<br>ChatGPT was also used in<br>empirical research to<br>develop MCQs, to compare<br>performance between<br>ChatGPT and students, to<br>help students evaluate<br>their own work, | ChatGPT performed<br>best for Italian exam<br>and did worst for the<br>French exams. | ChatGPT showed impressive<br>effectiveness in medical<br>education. |
| 28 | Bozkurt et<br>al.<br>(2025)(21) | Mixed-methods<br>systematic<br>review; 9;<br>Singapore, | 5; CINAHL,<br>Medline, Web of<br>Science, | Nursing students;<br>sample range (24-697) | AI was used on three<br>themes: (i) studies focus on<br>self-directed knowledge<br>increase or assessment, AI | AI perception is<br>positively correlated<br>with smartphone<br>addiction. AI ethics | Through findings, three themes<br>were developed on AI use on<br>medical education; AI facilitated<br>learning enhancement & |
|  |  | Republic of<br>Korea, Turkey,<br>Spain, Saudi<br>Arabia, Taiwan | ProQuest; Till 27<br>Dec, 2023 |  | perception, ethical<br>awareness, self-efficacy,<br>electronic fetal monitoring<br>(EFM), (ii) studies focus on<br>AI's role to improve<br>communication skills,<br>clinical reasoning skills,<br>breast self-examination<br>skills, (iii) studies focus on<br>the student's concerning<br>attitude, carrying multiple<br>factors, attitude towards<br>communication skills,<br>breast self-examination<br>satisfaction, anxiety, AI<br>self-efficacy, | awareness is positively<br>correlated with AI self-efficacy. Anxiety has a<br>positive correlation<br>with negative attitude<br>towards AI. AI self-efficacy is positively<br>associated with AI<br>ethics awareness and<br>AI behavioral intention<br>is positively associated<br>with both anxiety &<br>negative attitude<br>towards AI. | confidence, it possesses user<br>friendly interface &<br>accessibility, challenges &<br>concerns related to AI use. |
| 29 | Chan et al.<br>(2025)(34) | Scoping review;<br>145; USA,<br>Singapore,<br>Morocco,<br>China, Taiwan,<br>Turkey, Brazil,<br>Spain; 2020 -<br>2024 | 5; PubMed,<br>CINAHL, Embase,<br>Scopus, Web of<br>Science; Jan, 2015<br>- Nov, 2024 | Nursing<br>undergraduate<br>students | AI is implemented in<br>nursing simulation<br>education through 3<br>phases: (i) simulation<br>phase that creates<br>comprehensive patient<br>backstories through image<br>generating AI, integrates<br>of chatbot to assist in<br>history taking, (ii)<br>simulation<br>implementation through<br>AI applications & tools, including interactive feedback mechanism to assess student's emotion, creating virtual patients, (iii) simulation debriefing phase through precise feedback deliver, automated performance and analysis, providing personalized learning experiences. | Intermediate &<br>accurate feedback<br>improves students'<br>participation. Safe<br>practice opportunities<br>boost students'<br>confidence. | AI improves students' learning<br>experiences through realistic &<br>immersive simulation. It<br>enhances students' clinical<br>reasoning & communication<br>skills. Flexible training sessions<br>are efficient in saving time &<br>resources. Students had positive<br>perception on its usability. AI<br>improves students' engagement<br>& satisfaction in learning. It<br>reduces the workload for the<br>faculties. |
| 30 | Costa-Dookhan et al. (2025)(20) | Scoping review; 103; NR | 7; Medline, PsycInfo, Embase, CINAHL, Web of Science (Clarivate), Education Resources Information Center (ERIC), Library, Information Science and Technology Abstracts (LISTA) (EBSCO); 1 Jan, 2001 - 26 Mar, 2024 | Undergraduate medical students, postgraduate medical students, nursing students, dental students; sample range (6-7967) | AI was used to understand the needs of students to develop the curriculum. ChatGPT was used to create educational method, medical examinations, to assist in learning basic medical education (anatomy, physiology), diagnostic image reports, problem-based cases through clinical vignettes, and to enhance clinical & non-clinical skills. AI was used to evaluate communication and professionalism skills. It was also used to provide feedback on students' performance & competencies. AI was used to assess the quality of the feedback and evaluation, to evaluate students' reflection, to provide personalized learning, and predict learner remediation and academic performance. | Automated real-time feedback, communication skills, providing personalized learning, student's academic performance | Provided support for learning, reading diagnostic image reports, facilitated tradition teaching method and problem-based learning, providing feedback to students, assessment of feedback quality, evaluation of curriculum, & personal statement, prediction of learner's risk of remediation |
| 31 | Gerdes et al. (2025)(47) | Scoping review; 17; Taiwan, South Korea, US, China | 6; Medline, PubMed, CINAHL, Education Resources Information Center, World Science, Google Scholar; 1 Jan, 2013 - 23 Jul, 2024 | Undergraduate nursing students, graduate nursing students, nursing faculty | ChatGPT can be used by the faculty and the students for in mimicking National Council of State Boards of Nursing License Examination (NCLEX), developing Patient, Intervention, Comparison, Outcome, Time (PICOT) research questions, creating assignments, assisting in assignment preparation, developing patient documentation materials developing case studies.AI combined with Ted-talks can develop concise & effective presentation, combined AI & traditional method improved student's test score. | NR | Two themes were developed: (i) Generative AI and (ii) combined AI with traditional methods are being used in didactic nursing education. ChatGPT use improves students' critical thinking skills. Assignments using Imagery improved students' self-reflection on professional identity. ChatGPT helped to explore concepts, improved nursing values including patient-centered care, professionalism, patient autonomy, and evidence -based practice. |
| 32 | Hallquist et al. (2025)(58) | Systematic review; 82; NR | 1; PubMed; inception to 6 Jun, 2024 | Undergraduate medical students, post graduate residents | AI is used for different purposes in medical education including generating medical exam questions (board exam & admission test), preparing patient illness script, developing educational materials, as a learning aid in medical school, residency training and CME. NLP is used to improve clinical competency among residents, to assess residency applications, to write recommendation letters and clinical notes, and to analyze supervisor's feedback. AI is also used in clinical training like enhancing & assessing clinical skills, predicting student's correct answer through eye movement. | Stakeholder's competencies in AI use, integration of AI fundamental courses in medical schools & post-graduate training | ChatGPT is limited to evaluating text-based questions, being unable to assess table, graph, images. Assisted in writing recommendation letters and fellowship applications, enhanced clinical skills |
| 33 | Hui et al. (2025)(39) | Scoping review; 9; USA, Australia, Brazil. China, France, Norway, UK, Denmark, Taiwan; 2010 - 2024 | 4; Medline, Embase, PsycINFO, Scopus; NR | Medical students, Radiology residents | AI is used in 6 subject groups of radiology: general radiology, intervention radiology, chest imaging, neuroradiology, mammography, and musculoskeletal. AI is implemented in radiology education through (i) curriculum development by providing personalized training materials & simulation, (ii) diagnostic training by highlighting areas of interest & providing differential diagnosis, (iii) assessment. AI implementation was evaluated using a group experiment approach, survey on user experience, measuring time difference in reaching diagnosis. | Group of recipients, postgraduation education, small dataset & sample size | AI made a positive learning outcome, like significant improvement in exam scores through CT, MRI diagnosis. Trainees considered AI as a high value educational tool. AI algorithm could accurately identify & classify challenging images, and case difficulty level for trainees. |
| 34 | Janumpally et al. (2025)(49) | Scoping review; 20; USA, Canada, UK | 4; PubMed, Scopus, Web of Science, Google Scholar; Jan 2018 - Oct 2024 | Residents | Used in residency performance prediction and matching by analyzing application materials, identifying racial & gender bias in assessment, automated assessment process by classifying feedback quality, personalized learning recommendations, understanding entrustment, evaluating AI capabilities like in residency exam, and developing competency in trainee. | Linguistic differences, variation of emotion expressed in materials, automated feedback system, error severity | Reduced administrative workload, improved assessment accuracy, enhanced residency training by personalization, improved performance & success of trainees from underrepresented groups, facilitated trainee autonomy, and improved trainee competence. |
| 35 | Labrague et al. A (2025)(40) | Scoping review; 14; Singapore, USA, Morocco, Taiwan, South Korea, China, Finland, Turkey, Indonesia, Spain | 5; CINAHL, PubMed, Web of science, Scopus, Eric | Nursing undergraduate students; sample range (12-114) | Chatbot is used as Chatbot-facilitated simulations, Chatbot-assisted educational support and tutoring, Chatbot-enhanced learning & skill development, and Chatbot-assisted evaluation & assessment. Chatbots were used as supplementary tool and core components in learning modules and curricula. It was used as a tool for conflict management and clinical training. | AI integration method, type of chatbot system such as knowledge-based, AI-driven, lack of content accuracy, user satisfaction, usability, simulation anxiety, ability to address educational needs. | Chatbot improved learning achievement, skills, critical thinking and problem-solving skills, student's engagement, satisfaction & confidence. It also improved students' self-efficacy and communication skills. It enhanced user experience and assessment process by providing feedback & suggestions. The challenges remained lack of generalizability, inconsistency, technical reliability. |
| 36 | Labrague et al. B (2025)(61) | Scoping review; 16; Morocco, Taiwan, China, Singapore, Spain, USA, South Korea, Japan, Turkey | 5; PubMed, CINAHL, Scopus, Web of Science, ERIC | Nursing undergraduate students; sample range (4-300) | AI-base teaching categorized into 3 groups, (i) AI-driven simulation-based learning that includes AI-assisted simulation debriefing, Virtual patient simulation, Virtual reality simulation, AI-assisted screen-based simulation, (ii) AI-augmented instruction that includes CIDI-Model-based ChatGPT system, AI teaching assistant system, Task-based conversational AI agent, AI-integrated project-based learning, AI chatbot educational program, (iii) AI-generated content and tools that includes AI-generated imagery for pre-simulation strategy, AI-generated backstories & narrative videos, AI nursing course within curriculum, AI-generated case study | Mode of teaching, user satisfaction & engagement, usefulness, faculty workload, response accuracy, integration in curriculum | Simulation-based learning improved student participation, critical thinking, communication skills, self-efficacy, satisfaction, and learning outcome. Augmented instruction improved students' critical thinking and problem-solving ability, enjoyment of learning, self-directed learning ability, cooperative learning and personal growth. AI-generated content & tools improved student's strong emotional response, facilitated self-reflection on moral & professional identities, reduced simulation anxiety, improved student engagement, & learning, enhanced medical AI readiness among students, decreased workload of faculties, & saved time, and improved student's case management performance. |
| 37 | Lee et al.<br>(2025)(62) | Scoping review;<br>5; NR | 4; PubMed,<br>PsycInfo, Embase,<br>Web of Science;<br>inception to 16<br>Feb, 2025 | Medical students | Generative AI was used in<br>case-based learning,<br>simulation, content<br>synthesis, and developing<br>assessments tool | Diverse case vignettes<br>by AI make students<br>better prepared for<br>diagnose and treat<br>patients | Facilitated avoiding the<br>obstacles of traditional<br>methods' obstacles, generated<br>high quality illness scripts,<br>improve student's clinical skills<br>and ability to identify risk<br>factors, enhanced students'<br>competency to perform<br>psychiatric risk assessments,<br>enhanced content synthesis<br>process, maintained academic<br>standards. There are no<br>significant differences in<br>ChatGPT generated Script<br>Concordance Test and human<br>experts. |
| 38 | Lewis et al.<br>(2025)(52) | Scoping<br>review,; 18;<br>USA, Canada;<br>after 2021 | 6; Medline,<br>Embase, CINAHL,<br>ProQuest,<br>Education<br>Resources<br>Information<br>Center,<br>Education Full | Applicants to<br>medical schools &<br>residency | AI was used in screening<br>applications, predicting<br>ranking & sending<br>interview invitations, in<br>AI-generated personal<br>statement, potential in<br>detecting AI-generated<br>applications by lexical | Presence of gender<br>bias, evaluator's ability<br>to detect AI writings,<br>lack of AI detection<br>technology, | AI enhanced application<br>process, accurately predicted<br>applications to rank, detected<br>gender bias in letter of<br>recommendations, efficiently<br>analyzed both quantitative and<br>qualitative data of the<br>applicants. |
|  |  |  | Text; inception to 26 Aug, 2024 |  | analysis, in writing & analyzing AI-based letter of reference, and to discern gender bias. |  |  |
| 39 | Ma et al. (2025)(14) | Systematic review; 15; Turkey, USA, South Korea, Singapore, Palestine, Saudi Arabia, Morocco; 2022 - 2024 | 5; PubMed, Web of Science, Embase, Cochrane Library, CINAHL; inception to 1 Nov, 2024 | Nursing students; n = 1464 | AI was used in education, simulation training, and curriculum design | AI doctor: made students more engaging and mindful. Clinical setting: AI performed lower than control group. Students' lack of in-depth discussion & clinical experience can interfere with their reliability on AI-based answers. | Students' learning attitude and psychological effects, such as satisfaction, confidence, and interest in learning, improved. Learning outcomes, such as knowledge acquisition and practical skills, improved. Comprehensive clinical nursing competencies, such as communication skills, self-directed learning, critical thinking, and clinical understanding, were enhanced. |
| 40 | Ogbonnaya et al. (2025)(19) | Systematic review; 33; USA, China, UK, Ethiopia, Cameroon, Kenya, Mexico, Japan, Hungary, Latin America, Singapore, Republic of | 4; PubMed, IEEE Xplore, Cochrane Library, Science Direct; 2005 - 2024 | Medical students/surgical residents | AI was used in laparoscopic suturing training | New trainee, resource-constrained setting: AI is more beneficial. Personalized feedback & real-time performance tracking: Improved proficiency. Being novice: performance speed and higher completion rate got improved. Force- | AI system improved learning, better retention of knowledge & skills, improves precision, efficiency, and long-term retention of key suturing skills including needle manipulation, insertion technique, knot tying, and grip control. AI enhanced decision making. Real-time guidance improved patients' safety and surgical outcome. |
|  |  | Korea,<br>Netherlands,<br>Israel |  |  |  | feedback improved the<br>grip control, and visual<br>feedback was efficient<br>in developing surgical<br>suturing skills. |  |
| 41 | Rincon et<br>al.<br>(2025)(17) | Scoping review;<br>34; USA,<br>Canada,<br>Australia,<br>Germany,<br>Netherlands,<br>Turkey, China,<br>UK, Oman,<br>Bahrain,<br>Malaysia,<br>Brunei, Mexico,<br>Nigeria, India,<br>Chile, Poland,<br>Egypt, Taiwan;<br>2019 - 2024 | 3; PubMed,<br>Scopus, BIREME;<br>2019 - Oct, 2024 | Medical<br>undergraduate<br>students | AI was used in medical<br>education, curriculum<br>development, student<br>competency enhancement,<br>teaching & learning,<br>assessment & feedback. AI<br>did plagiarism detection,<br>clinical simulation,<br>homework support. | Interdisciplinary<br>learning &<br>collaboration with AI<br>will improve medical<br>education. Lack of<br>standardization limits<br>its efficiency. It needs<br>human support for<br>accuracy. Perception &<br>attitude towards AI<br>dependent on the<br>country: Nigeria<br>students felt<br>excitement & fearful,<br>Canadian students felt<br>crucial & urge for more<br>training, Indian<br>students felt worried<br>for losing empathy<br>due to overreliance on<br>technology, UK<br>students became<br>worried of limiting<br>career opportunity, | AI facilitated training,<br>interdisciplinary learning,<br>medical knowledge gain, and<br>providing assessment and<br>feedback through simulated<br>anatomy clinical sessions,<br>producing videos of dissection<br>and practices, allowing on-line<br>work, building personalized<br>effective education plan to<br>make education accessible.<br>Integration of AI in curricula<br>improved students' digital<br>literacy & skills needed for<br>effective patient collaboration.<br>AI improved self-learning,<br>students' satisfaction, good<br>performance in tests, and<br>retention of information. AI<br>could help to overcome<br>limitations of traditional<br>approaches like lack of cadaver<br>for anatomy dissection and risk |
|  |  |  |  |  |  | Arab students showed support for its' use in curricula. Training students is necessary for effective & consistent use. Gaps in medical school curricula acts as barrier for AI implementations in education. Lack of robust ethical framework hampers the ethical use of AI. | of handling toxic chemicals during dissection. |
| 42 | Shaw et al. (2025)(16) | Scoping review; 42; Mexico, Australia, USA, Canada, UK, France, Germany, Switzerland, Italy, Thailand, Iran, Japan, Taiwan, Brazil, Hong Kong, China | 4; PubMed, Medline, SCOPUS, Google Scholar, Jan, 2010 - Dec, 2022 | Medical undergraduate or post-graduate students | AI is used in surgical skill assessment (n = 30, 71%), radiology training, interactive learning, text interpretation | The expensive motion tracking sensor limits AI use in operation theatre. Trying to identify multiple problems rather than binary problem reduces the radiological diagnostic accuracy. | AI application enhanced competency-based surgical simulation, improved radiological image interpretation, facilitated interactive learning by replicating interaction with educators, and analyzed written papers in research. It helps students as a comprehensive & educational aid. AI in surgical training could decrease patient risks, provide feedback, and differentiate between expert & novice surgeons using data from wearable devices and surgical tools. Medical students improved their confidence in Anatomy and reasoning in clinical cases using the interactive chatbot. |
| 43 | Yeung et al. (2025)(43) | Scoping review; 4; China and Singapore | 6; Medline, OVID, CINAHL, Scopus, Web of Science, Google Scholar; January 2003 - June 2023 | Nursing students and interns; sample range (30-110) | NLP-based voice chatbots were used within leaning systems, which supported communication skills development through automated learning and evaluation modules | Social and cultural barriers such as lack of multilingual exposure, perceived lack of immersive and realistic experiences, challenges with accents and pronunciations within AI systems, unstable network or access, logistic issues (e.g., tight curriculum, time commitment, limited scope, high cost); rapid feedback and professional guidance, perceived psychological safety | High acceptance of communication skills learning systems, improved communication skills, performance, and self-efficacy, improved learning attitudes |
| 44 | Chytas et al. (2025)(54) | Scoping review; 9; NR | 4; PubMed, Scopus, ERIC, Cochrane library; 9 Oct, 2024 | Undergraduate medical students | ChatGPT was used to ask open-ended and multiple choice question on Anatomy. | Questions on blood supply and nervous system: ChatGPT found it difficult to answer questions on these topics. Article writing: ChatGPT showed below-par performance in writing articles on anatomy. ChatGPT showed better performance when customized by Anatomy field. | ChatGPT could provide advice on anatomical terminology, summaries for chapters, and help students to understand complex topics & their clinical implications, significance. It was not able to provide detailed information on anatomical variants, images, & citations for answers. ChatGPT helped teachers to outline lectures, create questions, and evaluate students' answers. ChatGPT's ability to answer correctly was average (44% - 66.6%) in non-comparative studies. ChatGPT showed 45% - 100% correctness in both open & MCQ answers comparing with other AI applications. ChatGPT was unable to write high-quality articles on anatomy. |
| 45 | Silmi et al. (2025)(42) | Systematic review; 9; NR | 3; PubMed, ScienceDirect, Google Scholar; 2020 - 29 Mar, 2025 | Undergraduate midwifery students | AI was used in midwifery education under 4 sub-themes, such as using predictive analysis, decision support systems, simulation & virtual learning, and generative | Poor data quality hinders effective AI applications. Lack of AI experts among educators makes it difficult to implement AI in education. | Predictive analysis helps students to identify early patient risk factors, to develop curriculum, to have practical experiences, to enhance clinical skills, and to provide timely care. Decision support system |
|  |  |  |  |  | AI. AI was used in midwifery curricula, faculty training, predicting students' success & failure, personalized learning & care, virtual patient simulations, Block learning model (TBM), scaffolded learning, theory-practice integration, evaluation of AI generated case study responses, development of digital literacy, early detection of complications, administrative automation, evaluation of midwifery skills, providing real-time simulation feedback, SWOT analysis, keyword co-occurrence analysis, strategy formulation | Resistant to change by midwifery educators & practitioners low down AI adoption. | helps students to make evidence-based decisions, evaluate treatment plans, predict complications, make informed decisions, to be prepared for using AI in clinical settings through training, and provide better patient care. Simulation & virtual tools provide an interactive and immersive learning experience, enhance student critical thinking & clinical reasoning, improve clinical decision making, making students confident, competent & prepared. Generative AI improves students' critical thinking, digital literacy, and analytical skills. |
| 46 | Al-Saadawi et al. (2025)(48) | Systematic scoping review; 21; USA, Canada, Germany, France, Taiwan, China, South | 4; Medline, Embase, CENTRAL, Scopus; inception to 30 Aug, 2024 | Orthopedic post-graduate residents; n = 273 | AI was used in orthopedic surgical training under two themes, orthopedic surgical skill refinement and enhancement of knowledge acquisition. | Type of AI model, feedback providing ability, prediction accuracy, efficiency of AI models | Using ML algorithm, an automated feedback system was made that assessed performance and provided feedback to surgeons. Deep learning algorithm provided real-time guidance signals educational contents ($p < 0.001$) and educational summaries ($p = 0.006$) than orthopedic fellows. |
| 47 | Aziz et al. (2025)(56) | Scoping review; 7; USA, others | 3; PubMed, ERIC, Web of Science; 1991 - Oct, 2023 | Students of Pharmacy; sample range (12-1389) | AI was used in the context of teaching and learning of Pharmacy, in both didactic & skill-based course, experimental curriculum, to build content knowledge & skills, predicting academic performance for admission, benefits and limitation of ChatGPT in pharmacy education. | Limitations like the need for AI training, technological failure, time needed to set up technology, adequate time needed to use AI in the classroom, missing data, narrow GPA score in performance prediction, and non-cognitive attributes being not considered in prediction models limits the use and accuracy of AI models. | AI can produce redundant, over-detailed content with missing references & copyright issues, helped students to build content knowledge on human anatomy, improve students' competence, enhance their thinking process. Students shows positive perception on the use of AI. However, it was found AI doesn't improve communication skill. |
| 48 | Qiu et al. (2025)(55) | Narrative review; 70; NR | 1; Scopus; inception to Sep, 2024 | Medical students | Generative AI was used to generate and answer different types of questions (MCQ, choose-n-from-many, true/false, open-ended questions) and different specialty questions in medical examinations. | Well-crafted prompts increase the efficiency of AI in producing high-quality questions. Prompts should include basic components like question types, topic names, number of questions, number of answers, correct answer requirements, aligning learning objectives, targeting specific audiences, difficulty levels, knowledge-based or clinical case-based, patient details or clinical vignette details. | ChatGPT 3.5 showed accuracy in different types of questions, like for MCQ (58%-61.1 %), true/false (23.1%), and open-ended questions (77.4% in psychiatry). Accuracy of ChatGPT 4 is for MCQ (81%), choose-n-from-many (69.8%), true/false (83%), open-ended questions (81% in family medicine). Regarding different specialties, ChatGPT 3.5 performed best in psychiatry (74.6% accuracy), and worst in pediatrics and gynecology & obstetrics (53.6% accuracy). ChatGPT 4 performed best in psychiatry (90.1% accuracy), and worst in osteology (67.4% accuracy). AI can provide incorrect answers causing medication error, patients harms, even death. |

Among the 48 included reviews, 29 were scoping reviews, 15 systematic reviews, two integrative reviews, one narrative review, and one meta-ethnographic analysis. The sample size for the included articles was wide in range (minimum four articles, maximum 278 articles). The study participants included medical school applicants, medical students (undergraduate and postgraduate, continuing education), residents and surgical trainees, educators, health informatics students, pharmacy students, nursing students, midwifery students, surgeons, nurse educators, and midwifery educators.

### Application of different AI technologies

Various AI tools and technologies were utilized in the education of health professionals (please see Appendix A). The majority of the tools and resources were based on generative AI, natural language processing, and machine learning technology (20–26). The tools ranged from wearable technology to virtual patient simulators, indicating the variety of ways AI technology was being used in the education and training of health professionals.

In terms of use of AI in health profession education, some examples included simulated clinical scenarios, intelligent tutoring systems, virtual patients, and virtual and augmented reality (13,27,28). In case of integrating AI in training and practice, authors mentioned various tools, for example, diagnostic algorithms, precision medicine, AI robotics, surgical AI systems, and AI-driven surgical robots (29–31). This review also found that AI technology was being used to identify resources such as fellowships and journals in various educational institutions (9).

### The outcomes of AI applications in HPE

The reviews included in this study identified various outcomes associated with the use of AI in HPE. Many of the included reviews identified that the use of AI in medical education helped to improve the learning environment and skills training through simulation, and AI was more accessible and provided flexibility in learning and training (13,15,32–35). For educators, AI played a supportive role in the design of curriculum and assessments (23). It also helped to evaluate learning progress, identify learning gaps, and assess achievements for accreditation or certification (26,33).

For medical students, the use of AI developed customized learning plans, provided them with feedback and personalized training, addressed the lack of skilled instructors, supported them in academic writing and research, and helped to improve digital literacy (22,23,36–38). AI use supported radiology trainees in identifying and classifying challenging images and complex cases (16,39). Use of virtual and augmented reality and simulation-based training enabled students to practice effective communication and was found to support critical thinking, engagement and self-efficacy (13,40). AI use also improved the understanding of AI in practice, prepared students to identify ethical challenges, fostered transparency in AI use, and built the competency and confidence of the students (9,25,41).

In one of the reviews on midwifery students and educators, it was found that AI improved learning engagement and efficiency, and allowed them more time for patient care (27). Decision-support systems for midwifery students helped them to make informed and evidence-based decisions and evaluate treatment plans, as well as prepared them to use AI in their own practice (42). In nursing education and practice, AI use helped to improve self-efficacy, communication skills and performance of nursing students (43). However, another review found that it was important to be equipped with knowledge and skills to assess the use of AI to provide appropriate care that was person-centered and compassionate (32).

In clinical settings, the use of AI offered opportunities for improving practical skills for both educators and students (e.g. improved suturing and knot-tying skills), and supported decision-making. It provided personalized training based on performance data, and could provide a more objective assessment compared to traditional methods (15,17,19,23,26,29,31,44–46). AI use combined with conventional methods could improve learning and critical thinking (39,47). Machine learning algorithms provided real-time feedback to orthopedic trainees (48). In addition, AI use reduced administrative workload for residents, improved assessment and enhanced training for trainees from underrepresented groups, and facilitated autonomy and competence (49).

### Factors influencing the applications of AI in HPE

The included papers stated that AI was being used as a learning support, in teaching and curriculum design, AI-assisted simulation, virtual simulation, AI-augmented instruction, assessment and evaluation of learning, critical thinking, clinical and communication skills, in academic writing and scholarly research, and clinical and practical settings to improve skills and build competence and confidence for students, trainees, educators and practitioners (14,16,21,23,26,29,35,37,40,42,50,51). AI was also utilized in evaluating teaching and getting feedback (24). In administrative procedures, the use of AI helped to screen applications, predict ranking, send interview invitations, and generate personal statements, while at the same time, it helped to detect AI-generated applications and plagiarism (17,52). There were a wide range of multilevel factors influencing the above-mentioned AI applications in different HPE contexts.

Positive attitude and engaging AI tools could improve learning motivation (13), and instant access to feedback and knowledge could enhance learning efficiency (35). However, the effectiveness of AI depended on the context and the levels of the trainees. In resource-constrained settings, novice trainees benefited more from the use of AI as it provided feedback and real-time performance monitoring (19). The effectiveness of AI in HPE was influenced by the language of learning (53), the subject matter (54) and the interest of educators and practitioners in adopting AI (31,42,46,50).

Challenges of AI implementation included constrained training data in terms of quality and availability (22,45). Use of ChatGPT to answer different types of questions showed moderate accuracy in various disciplines (55). Moreover, the responses were found to be redundant, overly-detailed, missing references, and susceptible to copyright issues (56). It was challenging to ensure accuracy and quality of the provided information, and misinformation and bias in training data could perpetuate discrimination and inaccuracies (22,31). Adopting AI in HPE posed difficulties because of the variety in cultural and regional healthcare practices and the high cost associated with getting access to AI tools, which could limit curriculum development and cause resistance among the educators (22,36,43,46,50). Use of AI could result in widening digital gaps and disparity among the different socioeconomic groups of people both within and across countries (22). Although the majority of the articles mentioned that the use of AI helped critical thinking, overreliance and dependence on large language models could also hamper critical thinking (35). In addition, there were multitudes of ethical challenges associated with the use of AI that could hamper academic integrity, induce bias, and create risks of plagiarism (33,53).

Lack of standardized AI curricula and parameters for AI competency, difficulty in evaluating the effectiveness of AI, and various technical challenges associated with developing AI tools and resources posed additional challenges (15,25,31,37,56). AI was differently adopted in HPE across countries due to cultural barriers, prioritization for medical skills and time constraints (28). Moreover, the institutions and educators were not ready for an AI-integrated curriculum (27,32,41,50,57). There was a lack of interdisciplinary collaboration for consensus on content and delivery, regulatory barriers, and a lack of funding (9,29,32).

## Discussion

This meta-review synthesized evidence on AI applications in HPE from 48 SLRs published in recent years, suggesting a growing interest of the HPE community in different use cases of AI, such as providing learning support and feedback, performing knowledge and skills assessment, and complementing other digital resources used in HPE. The use of AI in HPE was associated with better learning environments, customizing learning strategies, addressing learning gaps, advancing self-efficacy and communication skills, and promoting critical thinking among study participants. Also, this meta-review found that many HPE programs are adopting AI-related curriculum facilitating meaningful use of AI in HPE among learners and educators. The synthesized evidence informs recent developments in capacity-building and deployment of AI-based learning strategies, alongside user-centered and structural challenges of AI use in HPE.

The available evidence suggests that AI is increasingly being used both at the individual and institutional levels in HPE. Students using AI tools such as GenAI may benefit from simulated learning exercises that may result in better learning engagement (33,36). Moreover, augmenting learning plans using AI can possibly provide more time and opportunities to focus on patient-centered communication and personalizing patient care strategies. On the other hand, HPE educators can use AI to provide detailed feedback and optimize instructional delivery, which may improve the effectiveness of educational resources and activities (24,27). Also, AI tools can facilitate clinical simulations that can be used stand alone or in combination with other digital learning tools such as virtual or augmented reality (13,40).

This meta-review identified critical factors associated with AI use in HPE. At the user level, the attitude and interest of HPE students and educators may influence how AI resources can be adopted in HPE practices. The existing instructional practices and cultural issues may impact on how AI innovations are viewed by end users, which can affect their implementation in HPE (46,50). Moreover, digital literacy and competencies related to AI use can influence user adoption behavior. In addition, institutional readiness for AI deployment is a significant determinant of early adoption and sustainability of AI tools in HPE (21,37). The availability or lack of strategic frameworks, supporting educational resources such as AI curricula and content, and interdisciplinary collaborations can affect how HPE programs can benefit from emerging AI resources (17,25,26,32,36,46).

The findings of this review have several implications for informed development and implementation of AI-related resources in HPE. Many AI tools have shown promising abilities to provide feedback on academic activities, which can be further optimized to promote critical thinking, interpretive abilities, and higher order cognitive skills among HPE learners (33,51). Future use cases of AI tools need to be person-centered, with provisions for personalizing knowledge delivery approaches considering unique learning needs among HPE learners (20,27). Moreover, synergistic use of AI and non-AI digital resources can contribute to coordinated digital learning in HPE, as demonstrated in the use of augmented reality and other technologies alongside AI tools (13,37). In addition, most AI tools are widely used in task-based learning activities, whereas HPE learners can possibly benefit from brainstorming and reflecting on their broader learning and career goals with AI support (15). Compared to traditional learning spaces that primarily focus on task-based learning, AI-enabled systems may facilitate data-driven and reflective learning emphasizing individual personality traits, learning styles, motivations, grit, current academic standings, long term career goals, and optimizing academic and professional pursuits while ensuring broader biopsychosocial wellness (51). Identifying best practices for such integrative and diverse use of AI-based educational technologies may promote the digital transformation of HPE practices.

HPE users may experience complex challenges in adopting AI in learning activities, specifically if they have limited AI literacy and inadequate exposure to digital technologies in respective HPE contexts (28,58). It is necessary to address such gaps for the effective implementation of AI-based resources. All HPE users, including learners, educators, and administrators, should be supported with educational resources enabling AI literacy and application skills relevant to their HPE-related activities. This would require changes to accreditation standards, investments from governments and foundations, and publicly available educational content. Training the trainers on AI is a well-recognized approach to support HPE educators, whereas HPE students may benefit from AI-related educational exercises prior to deploying AI in HPE practices. Also, AI-related education should include perspectives on ethical use of technologies, which can be part of or complemented by medical humanities education (24,41). In addition, the HPE community needs to consider academic integrity-related issues in the context of AI use in higher education and develop strategic approaches for ethical and meaningful AI use in HPE (35).

Effective and sustainable use of AI in HPE would require institutional commitment and strategic measures, ensuring evidence-based adoption of AI tools maximizing user benefits. Instructional planners and academic leaders need to evaluate current learning management systems and practices and identify opportunities for cost-effective integration of AI in HPE. Educational leadership should strategically evaluate the scope of AI in HPE and broader health professions practices, which may contribute to learning outcomes as well as advancing health services and health-related outcomes in patients and populations. Moreover, pedagogical reform can be necessary considering the versatile use of AI in different levels of instructional delivery (32). Also, organizational capacities, including computational resources and human skills, need to be strengthened for optimal development and application of AI resources (37). Context-specific needs assessment and participatory development of AI use priorities can help academic communities to promote responsible use of AI. Moreover, strategies such as resource mobilization and optimization, exploring funding opportunities, partnering with AI service providers, and collaborating with other HPE institutions may contribute to institutional capacity building for AI. Furthermore, HPE-related professional academies and societies may advocate for local and regional coalitions supporting AI use in HPE. Recent institutional discourses on AI use in healthcare professionals and students appear to be early steps in this direction (59,60).

The findings of this meta-review have several implications for future research. Most of the included SLRs offered narrative synthesis that represented fewer longitudinal or experimental evidence, which is common in early-stage research on a phenomenon or intervention of interest such as AU resources. A lack of representative research from the global south indicates a relative paucity of research developments on AI use in HPE in limited resource settings. Moreover, most studies recruited medical and nursing students, whereas evidence representing other disciplines of HPE, such as public health sciences, nutrition, and allied health professions remains limited. Future studies should explore AI integration in diverse HPE contexts for broader and impactful use of those technologies in HPE practice. Also, the perspectives of HPE educators, administrators, and other stakeholders supporting digital tools integration in HPE could provide further clarity on AI implementation in the context of HPE practices. In addition, descriptive evidence on early use of AI in HPE provides limited insights on the possible mechanisms explaining how AI-based resources specifically contribute to HPE outcomes and support learners with varying learning needs and preferences, which could inform personalizing AI resources meeting their expectations and supporting sustainable adoption of future AI technologies in HPE institutions (28,30,50). Future research may address these issues and facilitate evidence-based practices on AI integration in HPE globally.

There are several limitations of this meta-review, which can inform the scope of this work and the interpretation of the synthesized findings. As meta-review methods consider review articles as unit of analysis, they provide a high-level overview of knowledge in a given topic or domain, limiting study-level insights that might be lost in evidence aggregation. Moreover, this meta-review included all possible types of SLRs, which provided heterogenous findings emerging from diverse HPE contexts. Therefore, the findings of this review illustrate summarized knowledge on AI use in overall HPE. In contrast, specific HPE user groups (e.g., medicine, nursing, dentistry) may need to assess the appropriateness of cross-disciplinary insights in their respective contexts. Furthermore, we included articles published in peer-reviewed journals, which could exclude reports, dissertations, or monographs, which appeared in non-journal sources. This possible selection bias, alongside publication bias limiting the exclusion of unpublished materials, may affect the generalizability of the findings of this meta-review. These limitations should be considered while interpreting our findings and addressed in future meta-research, whenever possible.

## Conclusions

This meta-review informs recent progress in AI use in different HPE contexts, factors associated with AI implementation, and outcomes among HPE users. While most reviews reported educational outcomes such as facilitating academic practices and supporting AI-driven digital learning among students, there are critical gaps in evidence that require careful consideration and strategic implementation of AI in HPE. Meaningful use of AI requires understanding user-centered issues and ethical perspectives supporting effective learning processes and outcomes. As AI technologies evolve over time, scholars and practitioners need to adopt practices using the best available evidence on AI integration in HPE.

## Conflicts of interest

The authors declare there are no conflicts of interest related to conducting this study.

## Funding sources

This review did not receive any specific funding from any sources. Dr Hossain is supported by the Presidential Frontier Faculty Grant by the University of Houston, TX.

## Authorship statement (according to CrediT roles)

Study conceptualization: MMH, PH, TJR, PM, WL; data curation: MMH, TJR, JD, ST; formal analysis: MMH, PH, TJR, JD, ST; funding acquisition: not relevant; investigation: MMH, PH, PM, JD, WL; methodology: MMH, PH, JD, ST, PM; project administration: MMH, JD, ST, WL; resources: MMH, PM, WL; software: MMH, TJR, JD, ST; supervision: PM, WL; validation: MMH, PH, ST, JD; visualization: MMH, PH, TJR; writing-original draft: MMH, PH, TJR, JD; writing-review editing: PH, ST, PM, WL; final revision and approval of the work: MMH, PH, TJR, JD, ST, PM, WL; agreement to be accountable for all aspects of this meta-review: MMH, PH, TJR, JD, ST, PM, WL.

## Data Availability

All data produced in the present meta-review are contained in the manuscript and additional data related to the referred articles are available in the published literature.

---

**Appendix A:**
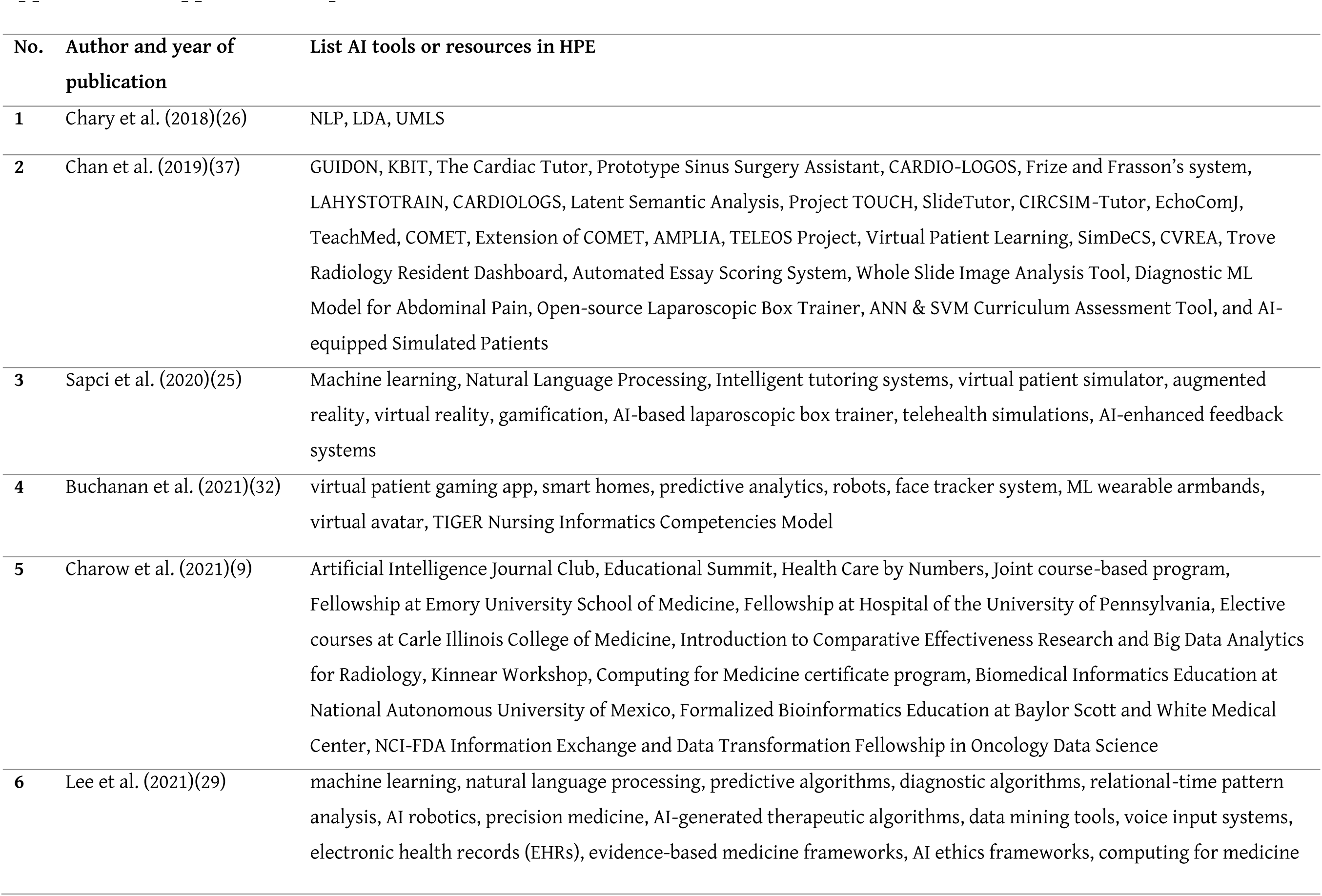
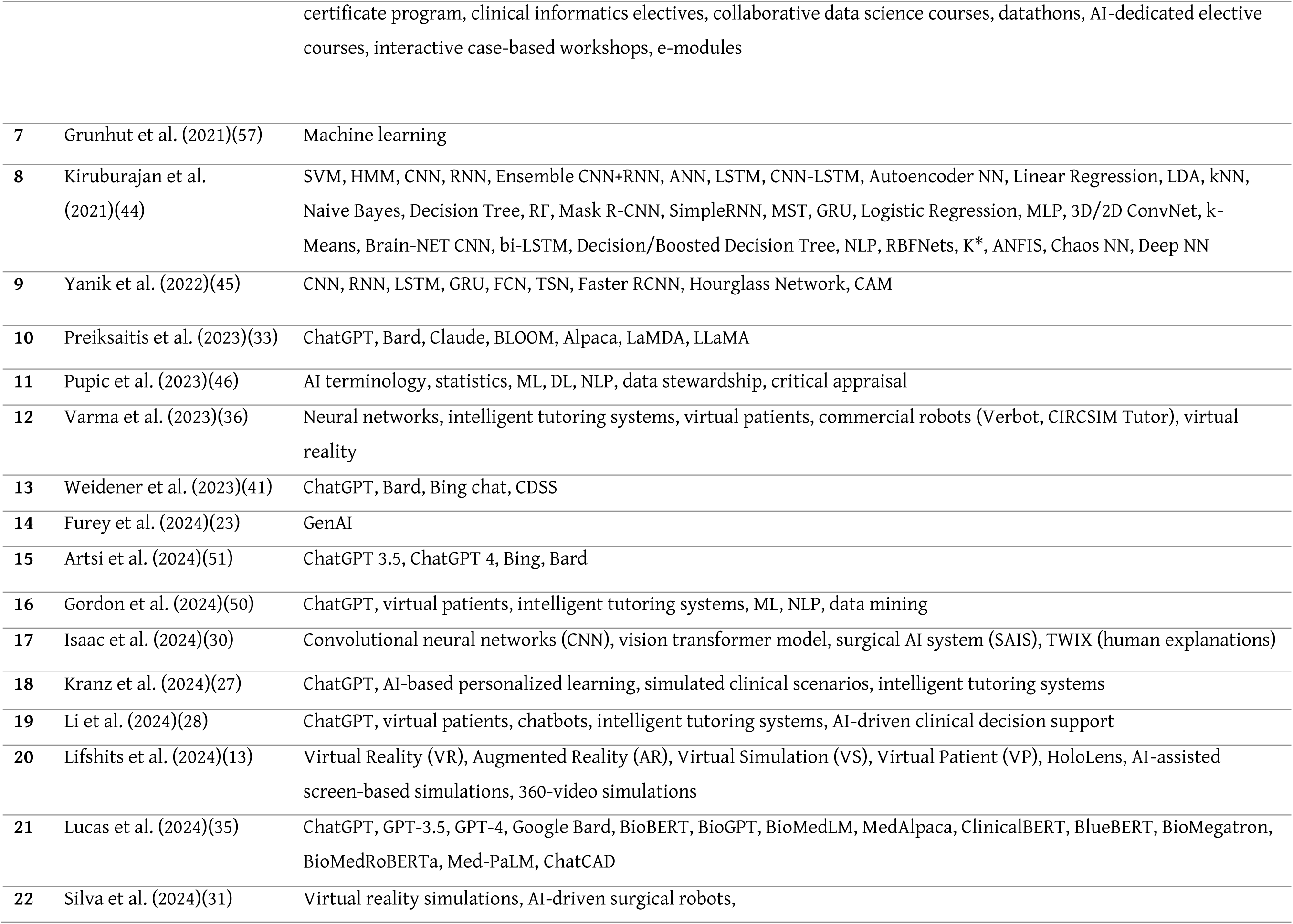
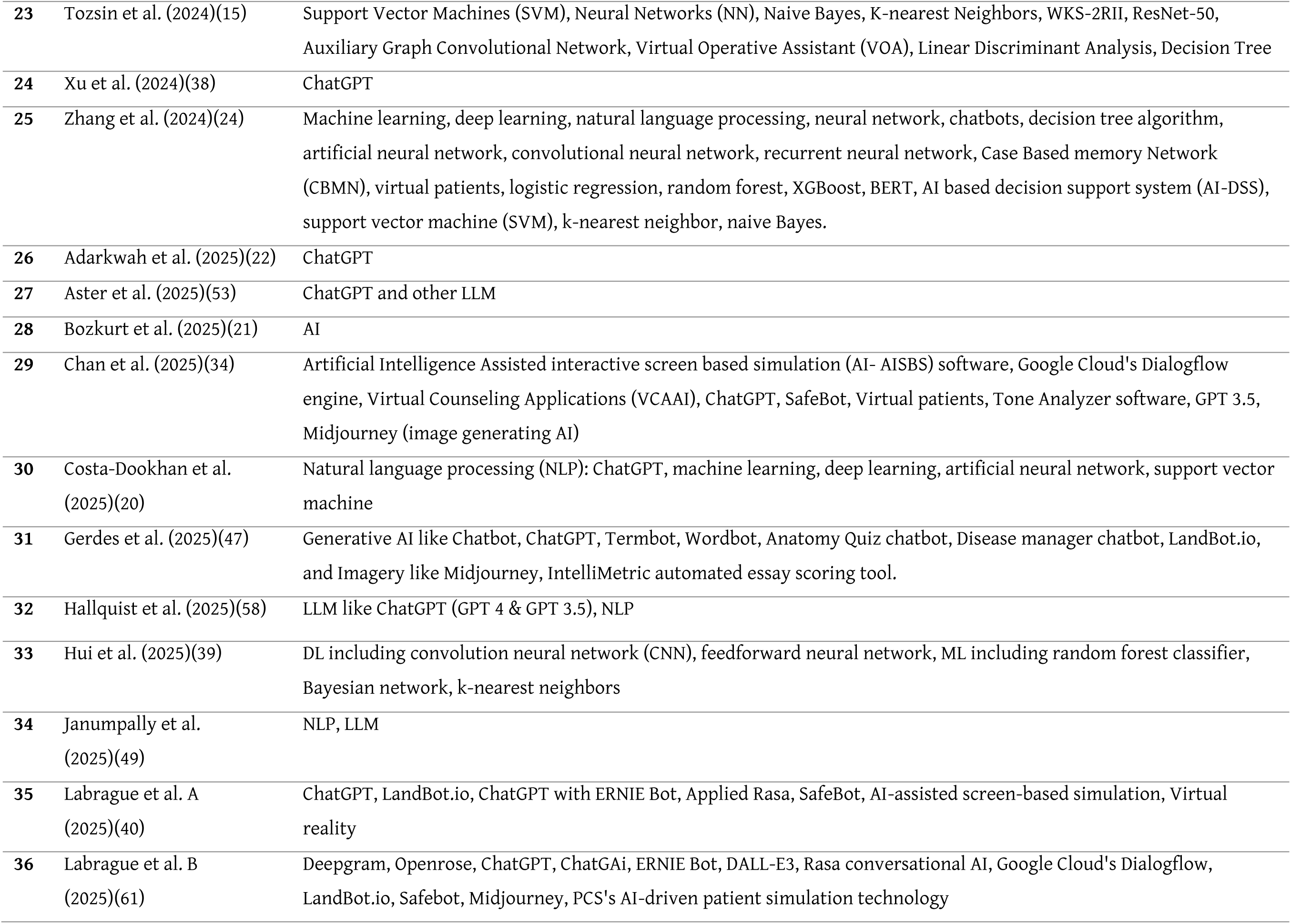
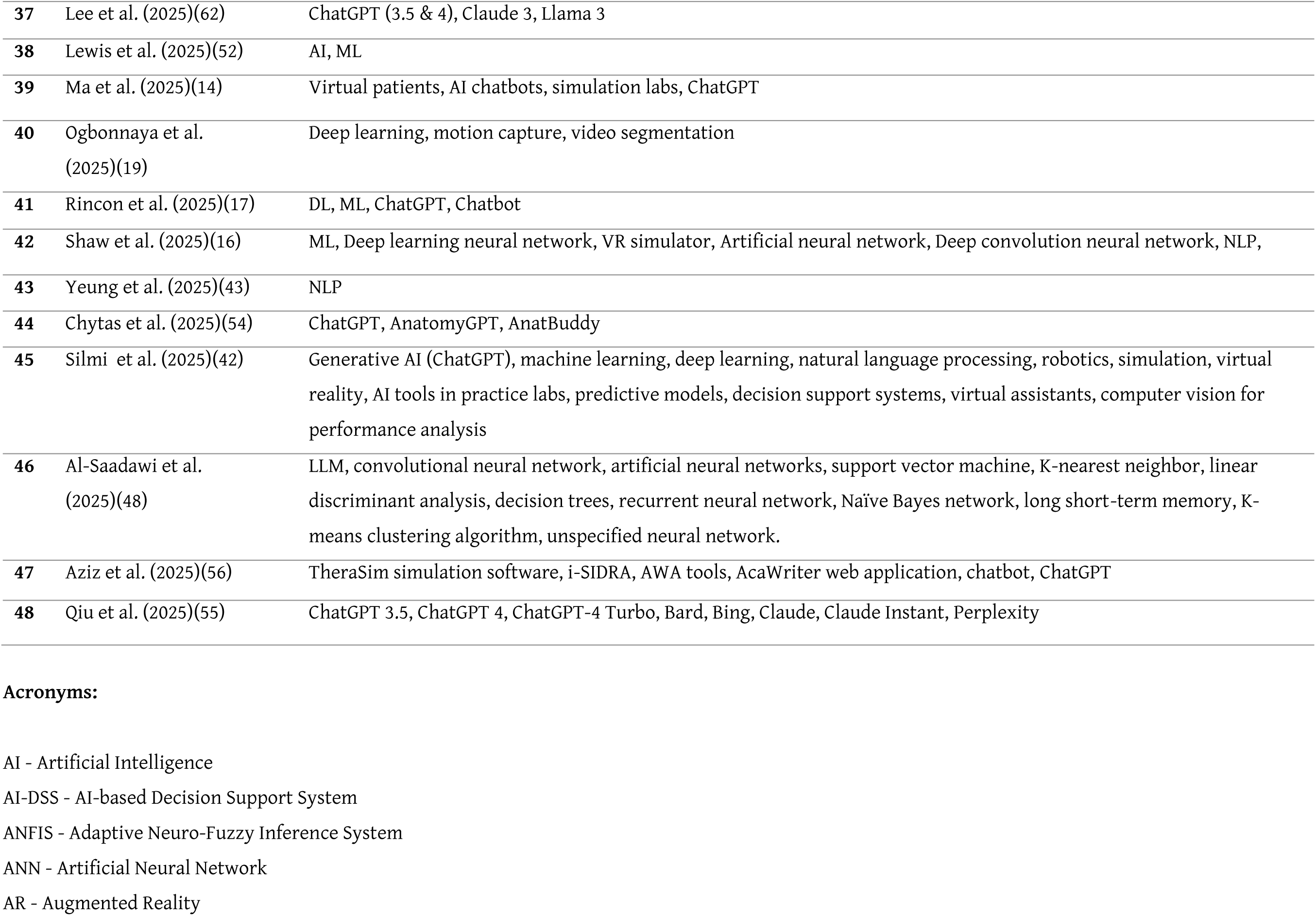
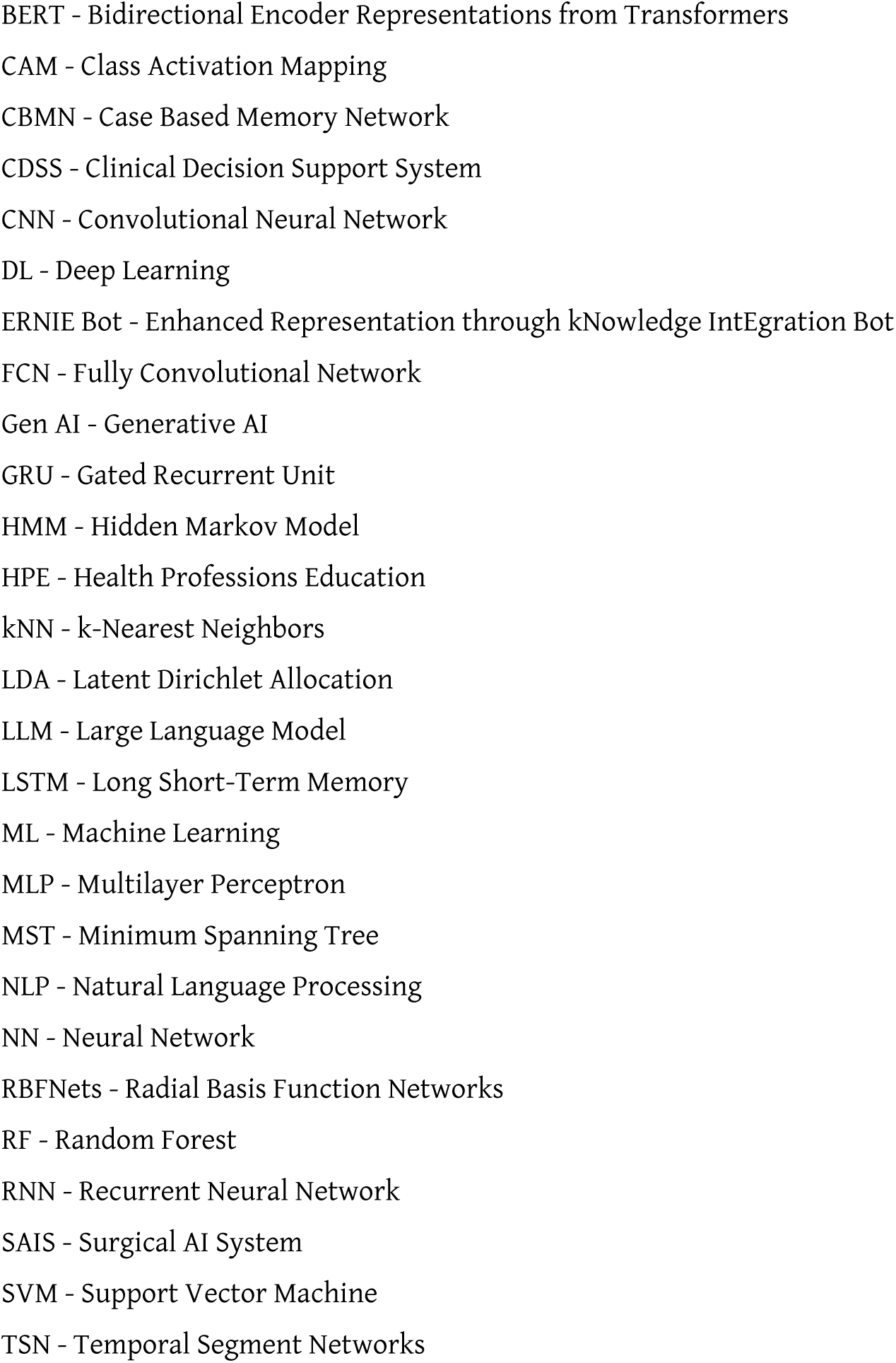
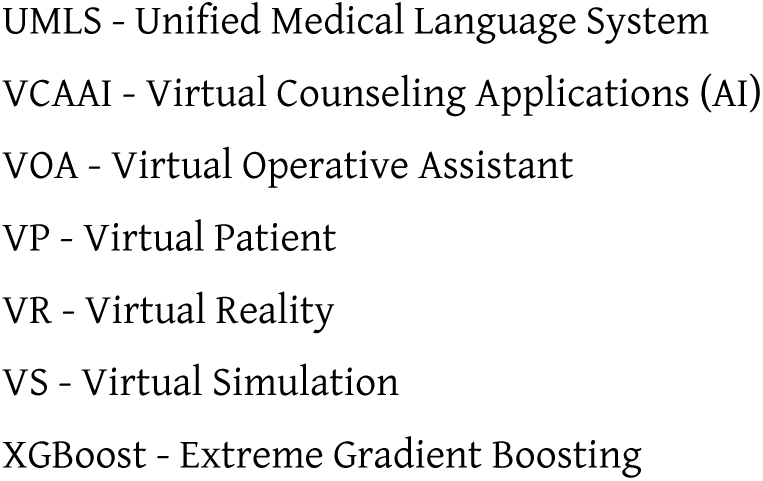
AI applications reported in included reviews.

## Notes

### Competing Interest Statement

The authors have declared no competing interest.

### Clinical Protocols

https://osf.io/dkbmn

## References

1. Abbasgholizadeh Rahimi S, Cwintal M, Huang Y, Ghadiri P, Grad R, Poenaru D, et al. Application of Artificial Intelligence in Shared Decision Making: Scoping Review. JMIR Med Inform [Internet]. 2022 Aug 9 [cited 2025 Sep 22];10(8):e36199. Available from: http://www.ncbi.nlm.nih.gov/pubmed/35943793

2. Khosravi M, Zare Z, Mojtabaeian SM, Izadi R. Artificial Intelligence and Decision-Making in Healthcare: A Thematic Analysis of a Systematic Review of Reviews. Health Serv Res Manag Epidemiol [Internet]. 2024 Jan 1 [cited 2025 Sep 22];11:23333928241234864. Available from: https://pmc.ncbi.nlm.nih.gov/articles/PMC10916499/

3. Dasta JF. Application of artificial intelligence to pharmacy and medicine. Hosp Pharm [Internet]. 1992 Apr 1 [cited 2025 Sep 22];27(4):312–5, 319. Available from: https://europepmc.org/article/med/10183640

4. Scott J, Scott R. Artificial intelligence: its use in medical diagnosis. J Nucl Med [Internet]. 1993 Mar 1 [cited 2025 Sep 22];34(3):510–4. Available from: https://europepmc.org/article/med/8441047

5. Harber P, McCoy JM, Howard K, Greer D, Luo J. Artificial intelligence-assisted occupational lung disease diagnosis. Chest [Internet]. 1991 Aug 1 [cited 2025 Sep 22];100(2):340–6. Available from: https://europepmc.org/article/med/1864103

6. Karthikeyan E, Sivaneswari S. Artificial Intelligence in Personalized Medicine for Head and Neck Cancer: Optimizing Prescriptions and Treatment Planning. Biomedical Materials and Devices [Internet]. 2025 Mar 27 [cited 2025 Sep 22];1–18. Available from: https://link.springer.com/article/10.1007/s44174-025-00320-6

7. Khalifa M, Albadawy M. Artificial Intelligence for Clinical Prediction: Exploring Key Domains and Essential Functions. Computer Methods and Programs in Biomedicine Update [Internet]. 2024 Jan 1 [cited 2025 Sep 22];5:100148. Available from: https://www.sciencedirect.com/science/article/pii/S2666990024000156

8. Kolla L, Parikh RB. Uses and limitations of artificial intelligence for oncology. Cancer [Internet]. 2024 Mar 30 [cited 2025 Sep 22];130(12):2101–7. Available from: https://www.ncbi.nlm.nih.gov/pmc/articles/PMC11170282

9. Charow R, Jeyakumar T, Younus S, Dolatabadi E, Salhia M, Al-Mouaswas D, et al. Artificial Intelligence Education Programs for Health Care Professionals: Scoping Review. JMIR Med Educ [Internet]. 2021 Dec 13 [cited 2025 Sep 22];7(4):e31043. Available from: http://www.ncbi.nlm.nih.gov/pubmed/34898458

10. Kim S, Kim SH, Kim H, Lee YM. Integrating artificial intelligence into medical curricula: perspectives of faculty and students in South Korea. Korean J Med Educ [Internet]. 2025 Mar 1 [cited 2025 Sep 22];37(1):65. Available from: https://pmc.ncbi.nlm.nih.gov/articles/PMC11900830/

11. Singla R, Pupic N, Ghaffarizadeh SA, Kim C, Hu R, Forster BB, et al. Developing a Canadian artificial intelligence medical curriculum using a Delphi study. NPJ Digit Med [Internet]. 2024 Dec 1 [cited 2025 Sep 22];7(1):1–10. Available from: https://www.nature.com/articles/s41746-024-01307-1

12. Feigerlova E, Hani H, Hothersall-Davies E. A systematic review of the impact of artificial intelligence on educational outcomes in health professions education. BMC Med Educ [Internet]. 2025 Dec 1 [cited 2025 Sep 22];25(1). Available from: https://pubmed.ncbi.nlm.nih.gov/39871336/

13. Lifshits I, Rosenberg D. Artificial intelligence in nursing education: A scoping review. Nurse Educ Pract [Internet]. 2024 Oct 1 [cited 2025 Sep 22];80:104148. Available from: https://www.sciencedirect.com/science/article/abs/pii/S1471595324002774

14. Ma J, Wen J, Qiu Y, Wang Y, Xiao Q, Liu T, et al. The role of artificial intelligence in shaping nursing education: A comprehensive systematic review. Nurse Educ Pract [Internet]. 2025 Mar 1 [cited 2025 Sep 22];84:104345. Available from: https://www.sciencedirect.com/science/article/abs/pii/S1471595325001015

15. Tozsin A, Ucmak H, Soyturk S, Aydin A, Gozen AS, Fahim M Al, et al. The Role of Artificial Intelligence in Medical Education: A Systematic Review. Surg Innov [Internet]. 2024 Aug 1 [cited 2025 Sep 22];31(4):415–23. Available from: /doi/pdf/10.1177/15533506241248239?download=true

16. Shaw K, Henning MA, Webster CS. Artificial Intelligence in Medical Education: a Scoping Review of the Evidence for Efficacy and Future Directions. Med Sci Educ [Internet]. 2025 Jun 1 [cited 2025 Sep 22];35(3):1803–16. Available from: https://link.springer.com/article/10.1007/s40670-025-02373-0

17. Rincón EHH, Jimenez D, Aguilar LAC, Flórez JMP, Tapia ÁER, Peñuela CLJ. Mapping the use of artificial intelligence in medical education: a scoping review. BMC Med Educ [Internet]. 2025 Dec 1 [cited 2025 Sep 22];25(1):526. Available from: https://pmc.ncbi.nlm.nih.gov/articles/PMC11993958/

18. Page MJ, McKenzie JE, Bossuyt PM, Boutron I, Hoffmann TC, Mulrow CD, et al. Updating guidance for reporting systematic reviews: development of the PRISMA 2020 statement. J Clin Epidemiol [Internet]. 2021 Jun 1 [cited 2025 Sep 22];134:103–12. Available from: https://www.sciencedirect.com/science/article/abs/pii/S0895435621000408

19. Ogbonnaya CN, Li S, Tang C, Zhang B, Sullivan P, Erden MS, et al. Exploring the Role of Artificial Intelligence (AI)-Driven Training in Laparoscopic Suturing: A Systematic Review of Skills Mastery, Retention, and Clinical Performance in Surgical Education. Vol. 13, Healthcare (Switzerland). Multidisciplinary Digital Publishing Institute (MDPI); 2025.

20. Costa-Dookhan KA, Adirim Z, Maslej M, Donner K, Rodak T, Soklaridis S, et al. Applications of Artificial Intelligence for Nonpsychomotor Skills Training in Health Professions Education: A Scoping Review. Vol. 100, Academic Medicine. Wolters Kluwer Health; 2025. p. 635–44.

21. Bozkurt SA, Aydoğan S, Dursun Ergezen F, Türkoğlu A. A systematic review and sequential explanatory synthesis: Artificial intelligence in healthcare education, a case of nursing. Vol. 72, International Nursing Review. John Wiley and Sons Inc; 2025.

22. Adarkwah MA, Badu SA, Osei EA, Adu-Gyamfi E, Odame J, Schneider K. ChatGPT in healthcare education: a double-edged sword of trends, challenges, and opportunities. Vol. 4, Discover Education. Discover; 2025.

23. Furey P, Town A, Sumera K, Webster CA. Approaches for integrating generative artificial intelligence in emergency healthcare education within higher education: a scoping review. Critical Care Innovations [Internet]. 2024 [cited 2025 Sep 22];7(2):34–54. Available from: www.criticalcareinnovations.eu

24. Zhang W, Cai M, Lee HJ, Evans R, Zhu C, Ming C. AI in Medical Education: Global situation, effects and challenges. Educ Inf Technol (Dordr). 2024 Mar 1;29(4):4611–33.

25. Hasan Sapci A, Aylin Sapci H. Artificial intelligence education and tools for medical and health informatics students: Systematic review. Vol. 6, JMIR Medical Education. JMIR Publications Inc.; 2020.

26. Chary M, Parikh S, Manini AF, Boyer EW, Radeos M. A review of natural language processing in medical education. Western Journal of Emergency Medicine. 2019;20(1):78–86.

27. Kranz A, Abele H. The Impact of Artificial Intelligence (AI) on Midwifery Education: A Scoping Review. Vol. 12, Healthcare (Switzerland). Multidisciplinary Digital Publishing Institute (MDPI); 2024.

28. Li W, Shi HY, Chen XL, Lan JZ, Rehman AU, Ge MW, et al. Application of artificial intelligence in medical education: A meta-ethnographic synthesis. Med Teach. 2025;47(7):1168–81.

29. Lee J, Wu AS, Li D, Kulasegaram K (mahan). Artificial Intelligence in Undergraduate Medical Education: A Scoping Review. Vol. 96, Academic Medicine. Wolters Kluwer Health; 2021. p. S62–70.

30. Isaac S, Phillips MR, Chen KA, Carlson R, Greenberg CC, Khairat S. Usability, Acceptability, and Implementation of Artificial Intelligence (AI) and Machine Learning (ML) Techniques in Surgical Coaching and Training: A Scoping Review. J Surg Educ. 2024 Jul 1;81(7):994–1003.

31. Silva C, Nascimento D, Dantas GG, Fonseca K, Hespanhol L, Rego A, et al. Impact of artificial intelligence on the training of general surgeons of the future: a scoping review of the advances and challenges. Vol. 39, Acta Cirurgica Brasileira. Sociedade Brasileira para o Desenvolvimento de Pesquisa em Cirurgia; 2024.

32. Preiksaitis C, Rose C. Opportunities, Challenges, and Future Directions of Generative Artificial Intelligence in Medical Education: Scoping Review. JMIR Med Educ. 2023 Oct 20;9:e48785.

33. Varma JR, Fernando S, Ting BY, Aamir S, Sivaprakasam R. The Global Use of Artificial Intelligence in the Undergraduate Medical Curriculum: A Systematic Review. Cureus. 2023 May 30;

34. Chan KS, Zary N. Applications and Challenges of Implementing Artificial Intelligence in Medical Education: Integrative Review. JMIR Med Educ. 2019 Jun 15;5(1):e13930.

35. Xu X, Chen Y, Miao J. Opportunities, challenges, and future directions of large language models, including ChatGPT in medical education: a systematic scoping review. Vol. 21, Journal of Educational Evaluation for Health Professions. Korea Health Personnel Licensing Examination Institute; 2024.

36. Hui M (Lucy), Sacoransky E, Chung A, Kwan BY. Exploring the integration of artificial intelligence in radiology education: A scoping review. Vol. 54, Current Problems in Diagnostic Radiology. Elsevier Inc.; 2025. p. 332–8.

37. Labrague LJ, Sabei S Al. Integration of AI-Powered Chatbots in Nursing Education: A Scoping Review of Their Utilization, Outcomes, and Challenges. Vol. 20, Teaching and Learning in Nursing. Elsevier Inc.; 2025. p. e285–93.

38. Weidener L, Fischer M. Teaching AI Ethics in Medical Education: A Scoping Review of Current Literature and Practices. Vol. 12, Perspectives on Medical Education. Ubiquity Press; 2023. p. 399–410.

39. Silmi H, Susanti AI. Revolutionizing Midwifery Education: A Scoping Review of Artificial Intelligence Methods. Jurnal Paedagogy. 2025 Apr 25;12(2):501.

40. Yeung JWY, Ho KHM, Cheung J, Tsang JTY, Chong DKY. Artificial intelligence-based technology in communication training in nursing education: A scoping review. Vol. 59, Journal of Professional Nursing. W.B. Saunders; 2025. p. 40–50.

41. Buchanan C, Howitt ML, Wilson R, Booth RG, Risling T, Bamford M. Predicted Influences of Artificial Intelligence on Nursing Education: Scoping Review. JMIR Nurs [Internet]. 2021 Jan 1 [cited 2025 Sep 22];4(1):e23933. Available from: https://nursing.jmir.org/2021/1/e23933

42. Kirubarajan A, Young D, Khan S, Crasto N, Sobel M, Sussman D. Artificial Intelligence and Surgical Education: A Systematic Scoping Review of Interventions. J Surg Educ. 2022 Mar 1;79(2):500–15.

43. Yanik E, Intes X, Kruger U, Yan P, Diller D, Van Voorst B, et al. Deep neural networks for the assessment of surgical skills: A systematic review. Journal of Defense Modeling and Simulation. 2022 Apr 1;19(2):159–71.

44. Pupic N, Ghaffari-Zadeh A, Hu R, Singla R, Darras K, Karwowska A, et al. An evidence-based approach to artificial intelligence education for medical students: A systematic review. PLOS Digital Health. 2023 Nov 1;2(11).

45. Gerdes M, Bayne A, Henry K, Ludwig B, Stephenson L, Vance A, et al. Emerging Artificial Intelligence-Based Pedagogies in Didactic Nursing Education: A Scoping Review. Vol. 50, Nurse Educator. Lippincott Williams and Wilkins; 2025. p. E7–12.

46. Al-Saadawi A, Tehranchi S, Ahmed S, Nzeako OJ. Exploring the Current Applications of Artificial Intelligence in Orthopaedic Surgical Training: A Systematic Scoping Review. Cureus. 2025 Apr 4;

47. Janumpally RK. The Role of Natural Language Processing in Graduate Medical Education: A Scoping Review. Cureus. 2025 Mar 24;

48. Lucas HC, Upperman JS, Robinson JR. A systematic review of large language models and their implications in medical education. Vol. 58, Medical Education. John Wiley and Sons Inc; 2024. p. 1276–85.

49. Gordon M, Daniel M, Ajiboye A, Uraiby H, Xu NY, Bartlett R, et al. A scoping review of artificial intelligence in medical education: BEME Guide No. 84. Vol. 46, Medical Teacher. Taylor and Francis Ltd.; 2024. p. 446–70.

50. Artsi Y, Sorin V, Konen E, Glicksberg BS, Nadkarni G, Klang E. Large language models for generating medical examinations: systematic review. BMC Med Educ. 2024 Dec 1;24(1).

51. Lewis LS, Hartman AM, Brennan-Cook J, Felsman IC, Colbert B, Ledbetter L, et al. Artificial Intelligence and Admissions to Health Professions Educational Programs: A Scoping Review. Nurse Educ. 2025 Jan 1;50(1):E13–8.

52. Aster A, Laupichler MC, Rockwell-Kollmann T, Masala G, Bala E, Raupach T. ChatGPT and Other Large Language Models in Medical Education — Scoping Literature Review. Vol. 35, Medical Science Educator. Springer; 2025. p. 555–67.

53. Chytas D, Noussios G, Paraskevas G, Vasiliadis A V., Giovanidis G, Troupis T. Can ChatGPT play a significant role in anatomy education? A scoping review. Vol. 109, Morphologie. Elsevier Masson s.r.l.; 2025.

54. Qiu Y, Liu C. Capable exam-taker and question-generator: the dual role of generative AI in medical education assessment. Global Medical Education. 2025 Jan 14;

55. Aziz MHA, Rowe C, Southwood R, Nogid A, Berman S, Gustafson K. A scoping review of artificial intelligence within pharmacy education. Vol. 88, American Journal of Pharmaceutical Education. Elsevier B.V.; 2024.

56. Grunhut J, Wyatt AT, Marques O. Educating Future Physicians in Artificial Intelligence (AI): An Integrative Review and Proposed Changes. J Med Educ Curric Dev. 2021 Jan;8.

57. Hallquist E, Gupta I, Montalbano M, Loukas M. Applications of Artificial Intelligence in Medical Education: A Systematic Review. Cureus. 2025 Mar 1;

58. Ethical Considerations and Implications for AI in Nursing | Oncology Nursing Society [Internet]. [cited 2025 Sep 22]. Available from: https://www.ons.org/publications-research/voice/news-views/10-2024/ethical-considerations-and-implications-ai-nursing

59. Principles for the Responsible Use of Artificial Intelligence in and for Medical Education | AAMC [Internet]. [cited 2025 Sep 22]. Available from: https://www.aamc.org/about-us/mission-areas/medical-education/principles-ai-use

60. Chan MMK, Wan AWH, Cheung DSK, Choi EPH, Chan EA, Yorke J, et al. Integration of Artificial Intelligence in Nursing Simulation Education A Scoping Review. Nurse Educ. 2025 Jul 1;50(4):195–200.

61. Labrague LJ, AL Sabei S, AL Yahyaei A. Artificial intelligence in nursing education: A review of AI-based teaching pedagogies. Vol. 20, Teaching and Learning in Nursing. Elsevier Inc.; 2025. p. 210–21.

62. Lee QY, Chen M, Ong CW, Ho CSH. The role of generative artificial intelligence in psychiatric education– a scoping review. BMC Med Educ. 2025 Dec 1;25(1).

